# Screening for breast cancer with mammography

**DOI:** 10.1101/2024.06.06.24308542

**Authors:** Peter C Gøtzsche, Karsten Juhl Jørgensen

## Abstract

**Background:** A variety of estimates of the benefits and harms of mammographic screening for breast cancer have been published and national policies vary. This is an update of a review previously updated 2013 and originally published 2001.

**Objectives:** To assess the effect of screening for breast cancer with mammography on mortality and morbidity.

**Search methods:** For this 2023 update, we searched PubMed, CENTRAL, the Cochrane Breast Cancer Group Specialised Register, the World Health Organization International Clinical Trials Registry Platform (WHO ICTRP) and ClinicalTrials.gov up to 28 February 2023.

**Selection criteria:** Randomised clinical trials (RCTs) comparing mammographic screening with no mammographic screening.

**Data collection and analysis:** Two authors independently extracted data. Study authors were contacted for additional information. Our main outcomes of interest were deaths due to breast cancer, any cancer, and due to any cause, and harms measured as overdiagnosis, number of mastectomies, lumpectomies, use of radiotherapy and of chemotherapy. Certainty of evidence was assessed with GRADE.

**Main results:** Eight eligible trials from Europe and North America that compared women offered screening mammography with women not offered screening were included. We excluded a trial because the randomisation failed to produce comparable groups. The eligible trials included 600,000 women in the age range 39 to 74 years.

The trials with adequate randomisation did not show a benefit in terms of a reduction in breast cancer mortality at 13 years (risk ratio (RR) 0.90, 95% confidence interval (CI) 0.79 to 1.02; 33 vs 30 deaths from breast cancer per 10,000 women; 3 RCTs; 292,153 participants). The findings at 24 years were similar to those at 13 years. Our certainty in both estimates was downgraded 1 level to ‘low’ due to changes in technology and treatment (indirectness) and due to imprecision. The trials with suboptimal randomisation showed a reduction in breast cancer mortality at 13 years with an RR of 0.75 (95% CI 0.67 to 0.83; 4 RCTs; 306,937 participants; very low certainty evidence).

In women below age 50 years, the results from adequately randomised trials did not show a reduction in breast cancer mortality at 13 years of follow-up (RR 0.87, CI 0.73 to 1.03; 28 vs 24 deaths from breast cancer per 10,000 women; 3 RCTs; 218,697 participants, low certainty evidence), nor for women at least 50 years (RR 0.94, CI 0.77 to 1.15; 53 vs 50 deaths from breast cancer per 10,000 women; 2 RCTs; 74,261 participants, low certainty evidence). Only one trial included women aged 70 years and above and could not provide a reliable effect estimate.

We found that breast cancer mortality was an unreliable outcome that was biased in favour of screening, mainly because of the risk of differential misclassification of cause of death. The trials with adequate randomisation did not find an effect of screening on total cancer mortality, including breast cancer, (RR 1.00, 95% CI 0.96 to 1.04; 288 vs 288 cancer deaths per 10,000 women; 3 RCTs; 292,954 participants; moderate certainty evidence; the follow-up was 10.5 years for Canada, 9 years for Malmö and 23 years for the UK age trial). All-cause mortality was not reduced (RR 0.98, 95% CI 0.94 to 1.03 after 7 years; RR 0.99, 95% CI 0.95 to 1.03 after 13 years; 324 vs 328 deaths per 10,000 women; and RR 1.01, 95% CI 0.99 to 1.04 after 24 years; 773 vs 765 deaths per 10,000 women; 2 RCTs; 250,671 participants; moderate certainty evidence) in the adequately randomised trials.

There were more lumpectomies and mastectomies combined in the screened groups, likely reflecting overtreatment (RR 1.31, 95% CI 1.22 to 1.42; 164 vs 214 operations per 10,000 women; 2 RCTs; 132,321 participants; moderate certainty evidence), as were the number of mastectomies alone (RR 1.20, 95% CI 1.08 to 1.32; 122 vs 102 per 10,000 women; 2 RCTs; 132,321; moderate certainty evidence). The use of radiotherapy was similarly increased whereas there was no difference in the use of chemotherapy (data for each outcome available from only one adequately randomised trial; low certainty evidence). Breast screening increased the number of breast cancer diagnoses (overdiagnosis)(RR 1.25, CI 1.18 to 1.34, 142 vs 113 diagnoses at 7 to 9 years of follow-up; 3 RCTs, 292,979 participants; moderate certainty evidence) in trials that did not screen the control group after the intervention phase.

**Authors’ conclusions:** Because of substantial changes in screening technology, treatment, and breast cancer awareness since the trials were done, the estimates from the trials are uncertain in today’s setting. As breast cancer mortality is an unreliable outcome that is biased in favour of screening, it is noteworthy that screening did not reduce total cancer mortality or total mortality. Breast screening does not meet the criteria that population screening should be based on rigorously performed randomised trials that show that the benefits outweigh the harms. No studies have been completed in low income countries and one small study from Colombia has yet to provide data on long term outcomes. Women, clinicians and policy makers should consider the trade-offs and the uncertainties carefully when they decide whether or not to attend or support breast screening programmes.

## Plain language summary

### What are the benefits and harms of screening for breast cancer with mammography?

#### Key messages

1. The most reliable studies did not show that breast screening with mammography reduces your risk of dying from breast cancer. While other studies did show this, they are less reliable.
2. Breast screening detects cancers that would never have caused death or disease in the absence of screening (overdiagnosis). This increases your risk of having a breast or a lump in your breast removed needlessly (overtreatment). Breast screening also causes false positive results, which is when the mammogram raises a suspicion of breast cancer that is later put to rest. False positive tests are common and can negatively affect qualiy of life also after a serious diagnosis is ruled out.
3. Substantial changes in technology, treatment, and greater public awareness of breast cancer since the studies were done means that the effects of breast screening are uncertain and that any benefit is likely smaller today.

#### What is breast cancer?

Breast cancer is a common cancer in women. The risk increases with age. It is a highly variable disease, with some cases developing rapidly and aggressively while others grow slowly or not at all. This complicates screening, which is more likely to detect slow-growing than fast growing cancers.

#### How does breast screening work?

Screening with mammography uses X-ray imaging to find breast cancer before a lump is felt. Screening is therefore intended for women in whom breast cancer is not suspected. The idea is to detect cancer earlier, when a cure is more likely. However, as some cancers have spread before screen detection is possible, one cannot assume that earlier detection is beneficial.

#### What did we want to find out?

We wanted to find out if mammography screening reduces the risk of dying from breast cancer; if it reduces the need for treatment; if it reduces the risk of dying overall; and to which extent it causes harms in terms of overdiagnosis and overtreatment of breast cancers not destined to cause death or symptoms in the lifetime of the women.

#### What did we do?

We searched for studies that compared screening with no screening.

We compared and summarized the results of the studies and rated our confidence in the evidence, based on factors such as study methods, study context, and size of studies.

#### What did we find?

We found eight trials that involved 600,000 women in the age range 39 to 74 years. The design of some studies was more reliable than others, mainly due to the way women were distributed between the screening group and the control group, but also for other reasons. The most reliable studies showed that screening likely did not reduce the risk of dying from breast cancer, regardless of age group. Due to the age of the trials, our certainty of this result was assessed as ‘low’. While other trials found a benefit from screening, their less reliable designs mean that our certainty in these resuts is even lower (’very low’).

Neither our analysis of the most or of the least reliable trials showed that screening reduced the risk of dying when all causes were considered.

The trials indicated with moderate certainty that the risk of being overdiagnosed happened to about 1 in 5 of those diagnosed with breast cancer during the period they were offered screening.

#### What are the limitations of the evidence?

There were important differences in the estimated benefit (reduced risk of death from breast cancer) between well and less well designed trials. The main design limitation was that some trial randomised groups of women rather than individuals, which meant that the two groups did not have a comparable risk of getting breast cancer.

There has been substantial changes in technology and treatment since the trials were done, and increased awareness of breast cancer and the importance of seeking care as soon as possible. This means that the possible benefit of breast screening today is likely quite different from that in the trials. It is uncertain if mammography screening delivers an important benefit today but we are certain it causes serious harms, overdiagnosis and overtreatment and false positive results. The included trials were performed in Europe and North America and did not consider effects in minorities.

#### How up to date is the evidence?

This review updates our previous review but did not identify new trials. The evidence is up to date to February 2023.

## Summary of findings

### Summary of findings 1 Summary of findings table - Screening with mammography compared to no screening with mammography for women not suspected of breast cancer (all age groups)

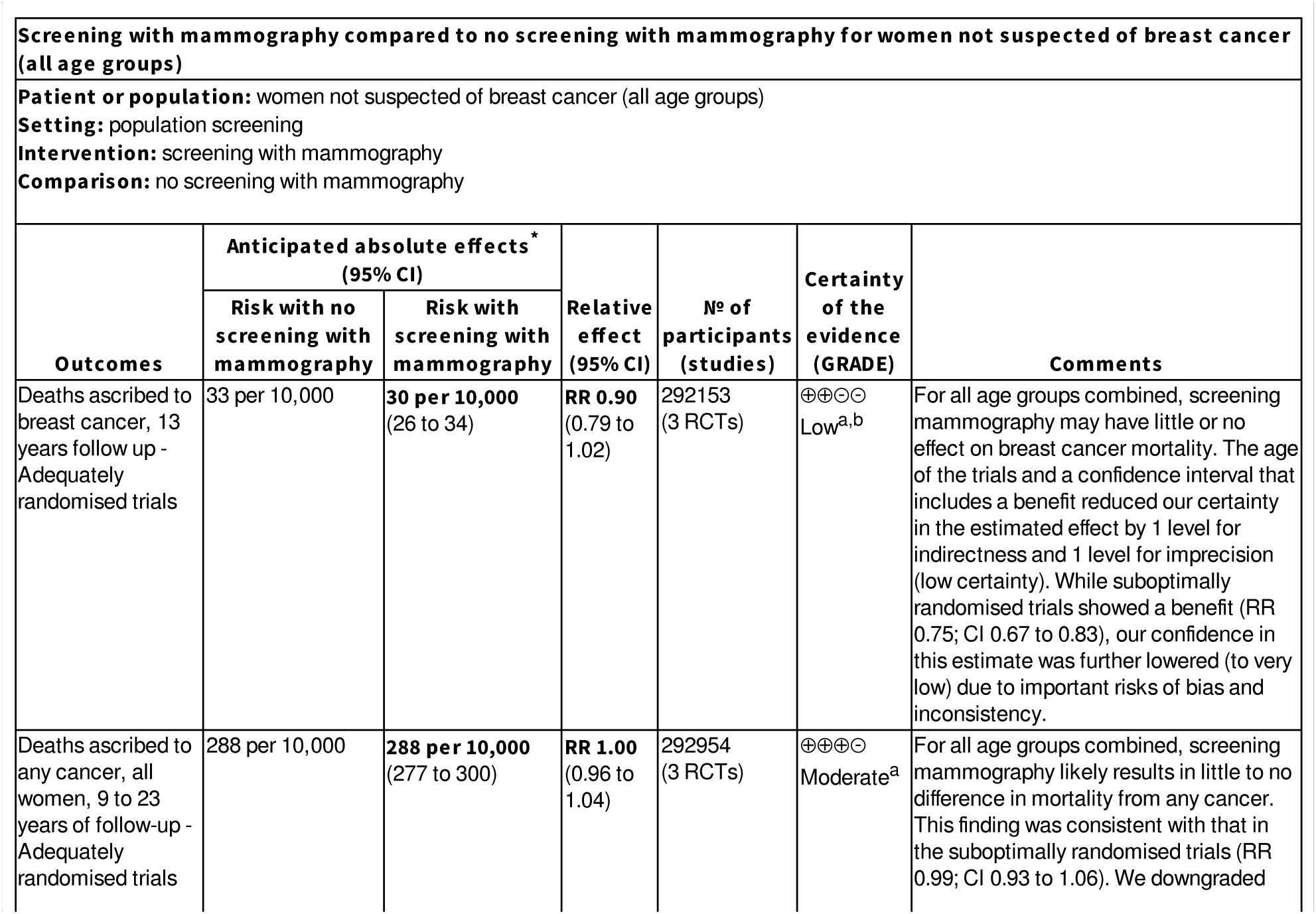
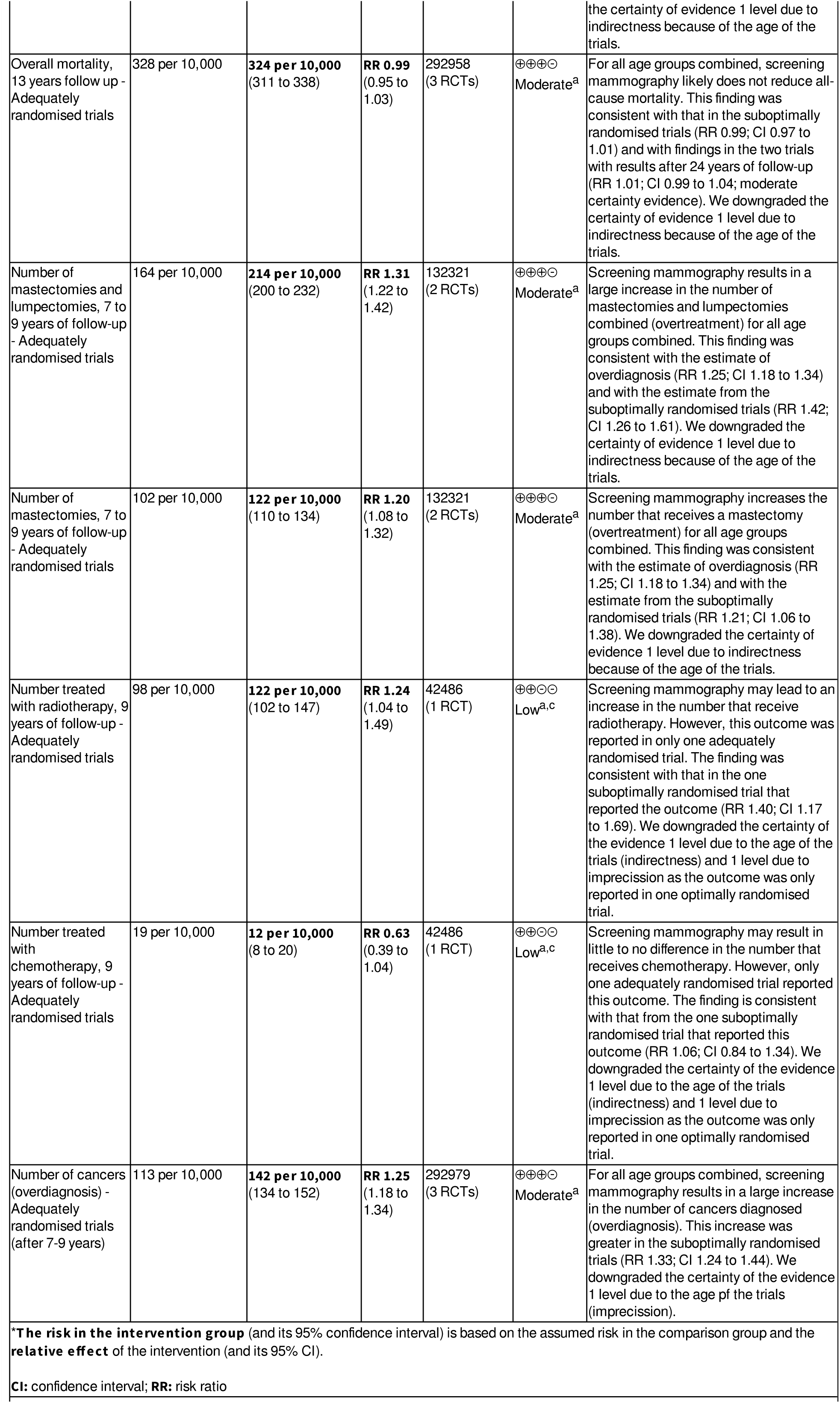
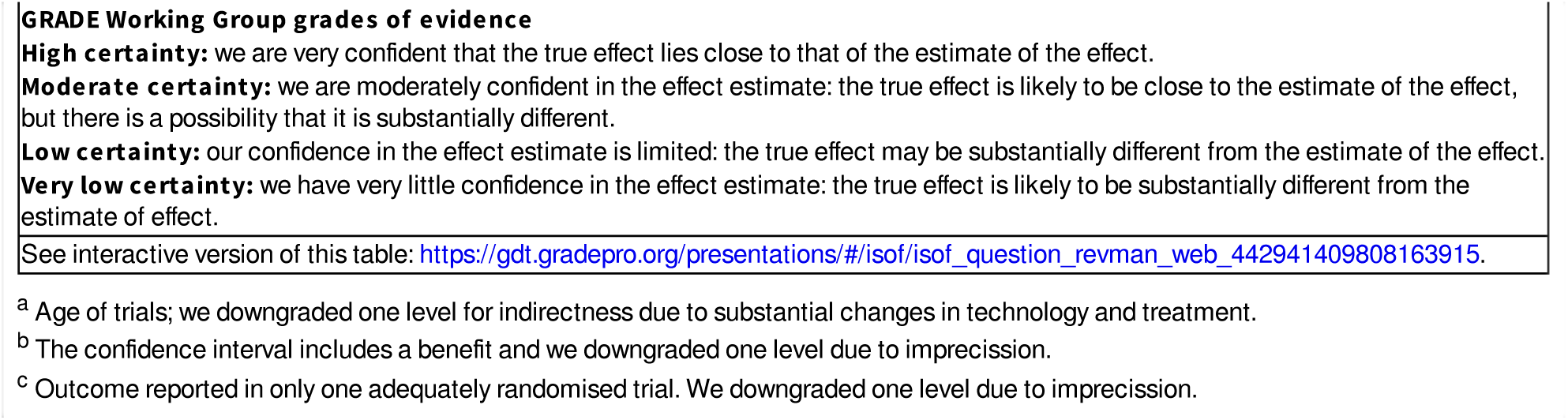

### Summary of findings 2 Summary of findings table - Screening with mammography compared to no screening with mammography for women not suspected of breast cancer (ages below 50 years)

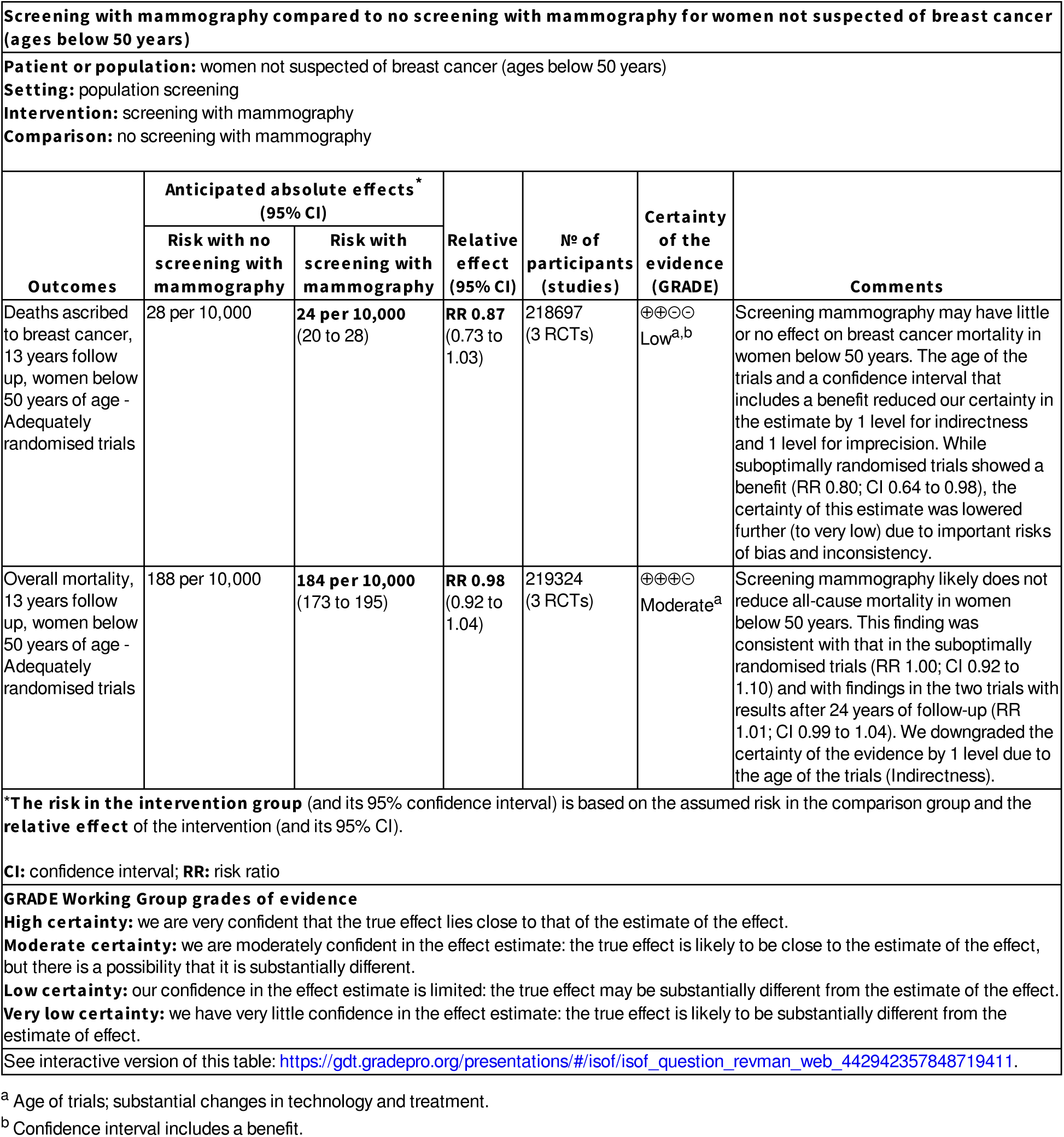

### Summary of findings 3 Summary of findings table - Screening with mammography compared to no screening with mammography for women not suspected of breast cancer (at least 50 years)

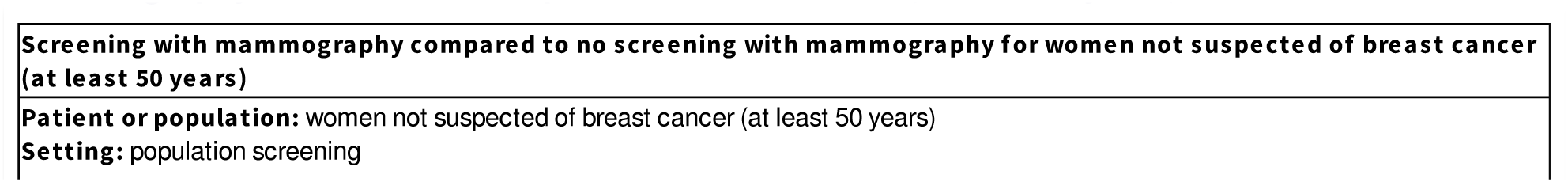
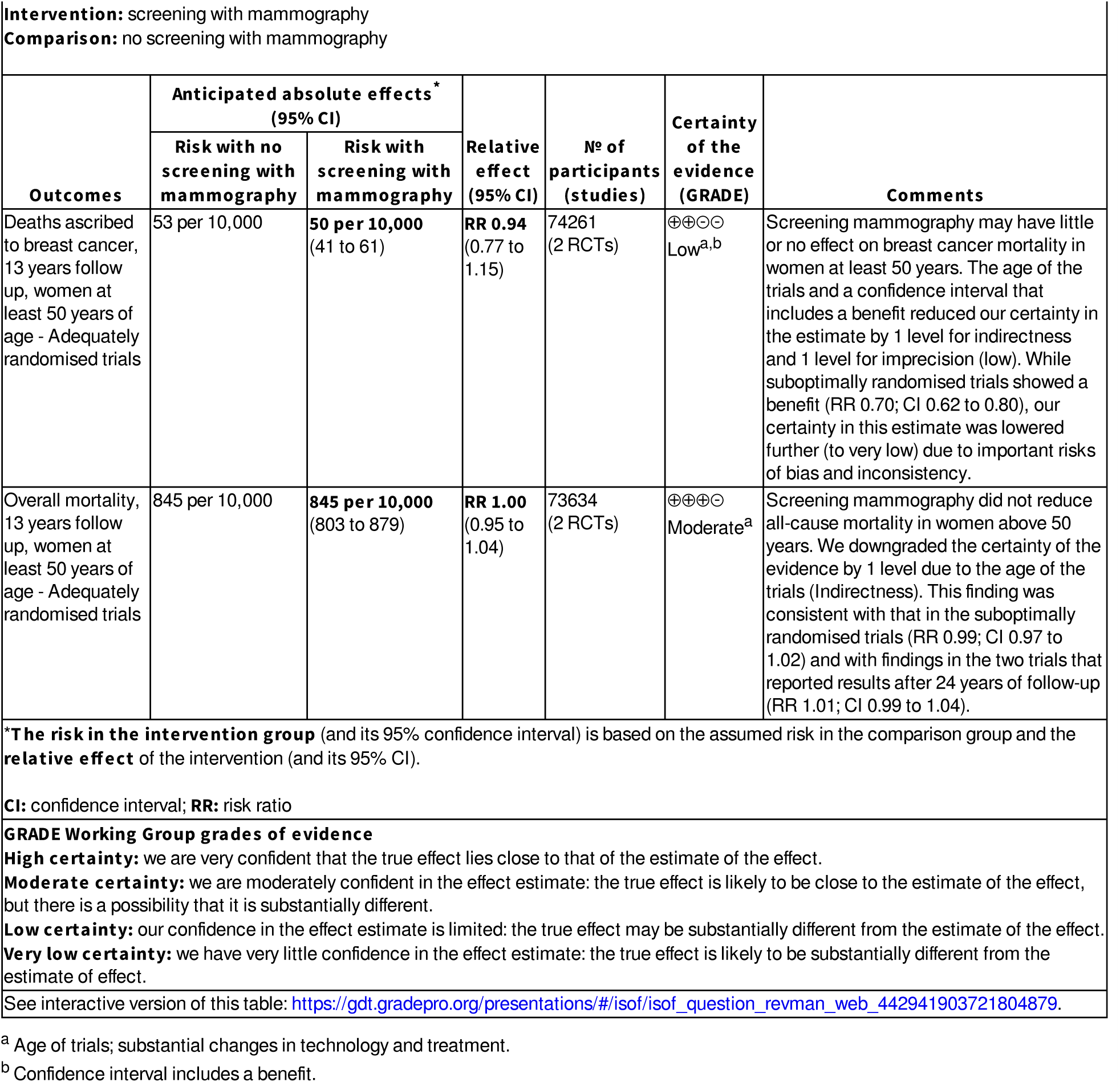

### Summary of findings 4 Summary of findings table - Screening with mammography compared to no screening with mammography for women not suspected of breast cancer (at least 70 years)

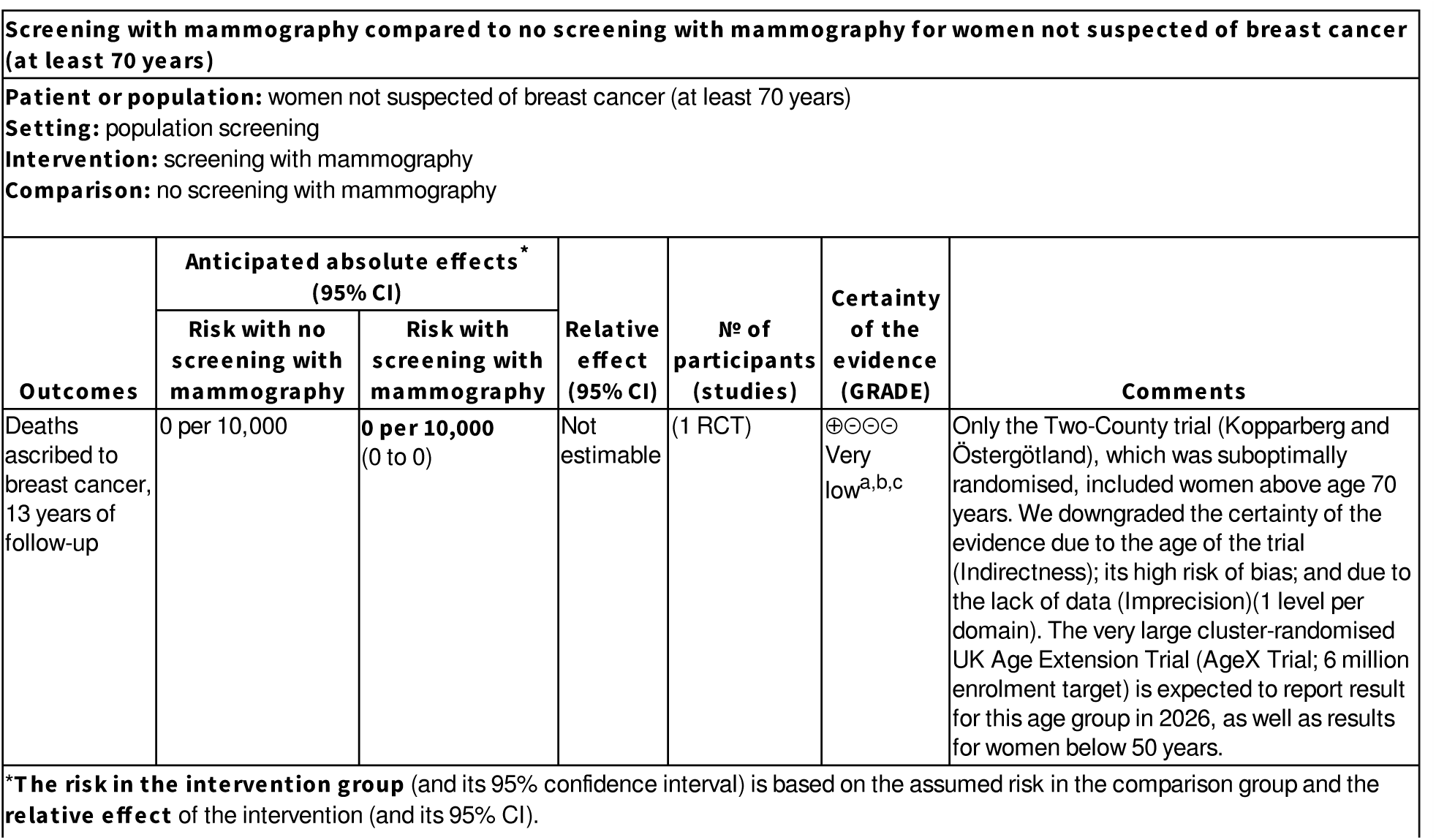
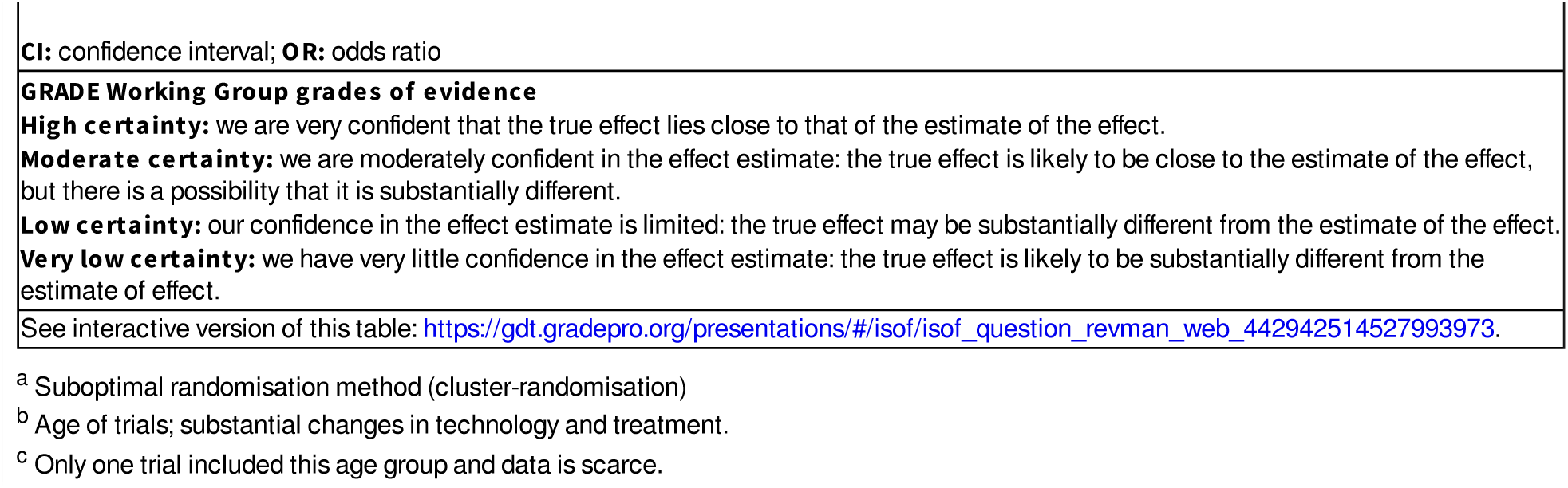

## Background

Breast cancer is an important cause of death among women worldwide with marked regional differences in disease burden (Arnold, 2022). Early detection through screening with mammography has the potential to reduce mortality, but it also leads to overdiagnosis and overtreatment (IARC, 2002). Since screening preferentially identifies slow-growing tumours (length bias) (Final reports, 1977; Fox, 1979), the harms of unnecessary treatment of overdiagnosed tumours could reduce or outweigh potential benefits.

The best way to reliably estimate the effectiveness of screening is with randomised trials at low risk of bias, as biases in the trials can easily erase or create the comparatively small effects of population based screening interventions (IARC, 2002). Large trials, involving 650,000 women, have been carried out in North America and Europe (Canada, 1980; Edinburgh, 1978; Göteborg, 1982; Malmö, 1976; New York, 1963; Stockholm, 1981; Two-County, 1977; UK age trial, 1991). It should be noted that, as for most interventions, the trials do not assess effects in minority groups or subgroups with poor access to care or particular risk profiles.

Several systematic reviews and meta-analyses have been published (Berry, 1998; Blamey, 2000; Cox, 1997; Demissie, 1998; Elwood, 1993; European Commission Initiative, 2020; Glasziou, 1992; Glasziou, 1995; Glasziou, 1997; Gøtzsche, 2000; Gøtzsche, 2011; Hendrick, 1997; Humphrey, 2002; IARC, 2002; Kerlikowske, 1995; Kerlikowske, 1997; Larsson, 1996; Larsson, 1997; Nelson, 2016; Nyström, 1993; Nyström, 1996; Nyström, 1997; Nyström, 2000; Nyström, 2002; Olsen, 2001a; Olsen, 2001b; Smart, 1995; Swed Cancer Soc, 1996; UK review, 2012; Wald, 1993).

The large number of reviews reflects the controversies surrounding mammography screening and the uncertainties of its effects in women of various ages. There is wide variation in screening policies between different countries, with some countries abstaining from introducing screening partly because of the lack of a documented reduction in all-cause mortality (Isacsson, 1985; Skrabanek, 1993; Swift, 1993). One area of concern is the potential for radiotherapy treatment of low-risk women, such as those who have their cancers identified at screening, to increase all-cause mortality because of adverse cardiovascular effects (EBCTCG, 1995; EBCTCG, 2000). Harms from radiotherapy has likely diminished in today’s setting, but increased overdiagnosis with greater sensitivity of the screening test may mean more women are exposed. Overdiagnosis of breast cancer is acknowledged as the most important harm of breast screening (Barratt, 2015), although uncertainty around its magnitude remains, as it does for its main benefit (UK review, 2012). Overdiagnosis is the detection of cancers that would never have appeared in the lifetime of the individual in the absence of screening and is perhaps best known from prostate cancer screening.

In addition, there is concern that cause of death has not been ascribed in an unbiased fashion in some of the trials due to lack of blinded outcome assessment. Finally, carcinoma in situ is much more likely to be detected with screening mammography and although less than half of the cases will progress to become invasive (Nielsen, 1987; Welch, 1997), only 18% after 20 years of follow-up in the Canadian trials (To, 2014), these women are often treated with surgery, drugs, and radiotherapy.

Since the trials were performed, major advances in earlier diagnosis of clinical cancers with increased breast cancer awareness (Rostgaard, 2010) and in breast cancer treatment (Riemsma, 2010) have happened. This has resulted in reductions in breast cancer mortality of 30% or more, most pronounced in younger women, below the age range invited for breast screening (Autier, 2011a). Collectively, this means that the relevance of the results of the original trials have diminished in today’s setting. This has led some prominent guideline groups to use modelling studies (Draft USPSTF recommendation, 2023) as the basis for screening recommendations, which has raised concerns (Harris, 2024; Woloshin, 2023). As modelling studies come with substantial uncertainty, we do not include them in this update. We do not include observational studies either for the same reason, which is in agreement with other reviews (UK review, 2012; European Commission Initiative, 2020).

Meta-analyses of screening are often deficient (Walter, 1999) and few of the meta-analyses listed above have taken account of the risk of bias in the individual trials or considered harms as well as benefits. We have identified important weaknesses in the trials (Gøtzsche, 2000; Gøtzsche, 2000a; Gøtzsche, 2004; Gøtzsche, 2011) and have now updated our Cochrane Review with additional data from two of the least biased trials, while no new trials contributed data. This update also include addition of Summary of Findings tables and a GRADE assessment of the certainty of the evidence for individual outcomes.

### Description of the condition

Breast cancer is the most common cancer among women worldwide (World Cancer Data, 2023). It is a highly variable disease with some cases growing rapidly and aggressively and some slowly or not at all. Like for most cancers, breast cancer incidence increase with age and the rapidity of growth slows. This has implications for the usefulness and the benefit to harm balance of screening. Breast cancer is also known to be able to return many years after treatment, sometimes after 20 years or more. This means it is difficult to say a breast cancer is ‘cured’, likely due to micro-metastases being present in the body after treatment. Breast cancer is thus regarded as a potentially systemic disease which affect treatment choices, although the exact time of metastasis is difficult to determine. Ideally, breast screening should detect cancers prior to metastasis to affect mortality and maximise chances of curative treatment.

### Description of the intervention

Screening with mammography uses X-ray imaging to find breast cancer before symptoms are noticed. The X-ray screening test generally does not provide the diagnosis but may raise a suspicion. Those who are found to have suspicious lesions are subjected to further tests such as ultrasound, MRI scans, and biopsies. Those in which these follow-up tests have excluded a diagnosis of breast cancer are said to have experienced a false-positive screening mammogram. Screening mammography differs from diagnostic mammography in that those who recieve screening mammograms do not have symptoms or a suspicion of breast cancer. Mammography screening programmes thus invite women without symtoms of breast cancer. While the screening test uses X-rays, the dose is quite low and presents a very small risk in itself (IARC, 2002).

### How the intervention might work

The idea with breast screening is to detect and treat breast cancer earlier, when a cure may be more likely and treatment possibly less aggressive. Breast screening is commonly offered annually, biennially, or triennially, varying between countries (IARC, 2002). The most commonly screened age range is 50 to 69 years, but women in their 40s and 70s are sometimes targeted as well (IARC, 2002). The interval between screening rounds means that rapidly growing cancers are less likely to be detected through screening and more likely to appear between screening rounds. This phenomenon is called ‘length bias’, as slower growing cancers has a greater length of time to allow screen detection (Welch, 2004). Cancers detected between screening rounds are known as ‘interval cancers’. Interval cancers are thus, on average, the more aggressive and fast growing ones (Welch, 2004), and they cannot benefit from breast screening. The fact that breast screening is best at detecting cancers that grow slowly or not at all is the cause of its major harm: overdiagnosis. These women would have lived without symptoms from their cancer before they died from another cause. It means they unnecessarily experience the stress of a breast cancer diagnosis and the harms of breast cancer treatment (Barratt, 2015). This is different from false positive results, where the suspicion of cancer is later dismissed through follow-up tests. False positive results affect far more women and result in important psychological harms (Brodersen, 2013).

Earlier detection of breast cancer makes women live longer with the diagnosis, increasing the apparent survival time with the disease. However, survival time is a deceptive outcome, as earlier diagnosis invariably increases survival time, even if screening does not reduce mortality. This phenomenon is called ‘lead time bias’ (Welch, 2004). Breast cancer mortality can also be a deceptive outcome, as cause of death is difficult to ascertain; as overdiagnosis leads to overtreatment, which increases deaths; and as a cancer diagnosis in itself increases cardiovascular mortality and risk of suicide (Fang, 2012).

### Why it is important to do this review

While earlier detection of cancer is well-documented to reduce incidence and mortality for some cancers, i.e. cervical (Raffle, 2003) and colon cancer (Jodal, 2019), this is not common. For example, large randomised trials of screening for ovarian cancer have shown that, although screening effectively brought the time of diagnosis forward and detected cancer at an earlier stage, this did not lead to reduced ovarian cancer mortality (Menon, 2021; Prorok, 2018). For other cancers, it is the balance between a possible reduction in disease-specific mortality and important harms that questions the rationale for screening, e.g. prostate cancer screening with the prostate specifc antigen (PSA) test. The fact that we cannot be certain that screening reduces disease-specific mortality or that a benefit outweighs the harms, is the reason that guideline groups such as the UK National Screening Committe require evidence from high-quality randomised trials to recommend it (UKNSC Criteria).

Mammography screening is being offered to billions of women world wide when they reach a certain age. It is resource intensive and the balance between its benefits and harms is contested. It is therefore important to know what the benefits and harms are. This is an update of a Cochrane review first published in, 2001, and updated in, 2009,, 2011, and, 2013. The previous versions questioned whether the data from the randomised trials supported the value of breast screening and identified important biases in key trials. Updated mortality results from two low risk of bias trials and new requirements for reporting in Cochrane reviews (GRADE assessments and Summary of Findings tables) necessitated this update.

## Objectives

To study the effect of screening for breast cancer with mammography on mortality and morbidity.

## Methods

### Criteria for considering studies for this review

#### Types of studies

Randomised clinical trials. We did not limit our inclusion of trials based on location, setting, definition of condition, demographic factors, the setting of the screening intervention (hospitals, private clinics, mobile units, etc.), or method of diagnosis (i.e. digital or print mammograms).

Trials using suboptimal randomisation methods such as cluster randomisation, were included but evaluated in separate subgroup analyses. In cases where substantial heterogeneity between trial results could be explained by use of optimal versus suboptimal randomisation methods, we based our conclusions on trials with optimal randomisation methods and chose not to present summary estimates including all trials.

We have discussed recent observational studies in this review as they have provided important contextual knowledge, e.g. in relation to evidence of overdiagnosis and other harms of screening in today’s setting.

However, such studies were not formally included or analysed in this review.

#### Types of participants

Women without clinically suspected or previously diagnosed breast cancer.

#### Types of interventions

The intervention was screening mammography with X-ray imaging. We included trials whether they used film or digital mammograms and did not exclude trials if they used technology such as computer-assisted detection. We did not include trials of tomosynthesis, magnetic resonance imagig (MRI), or ultrasound. Breast screening using X-ray in combination with ultrasound is evaluated in another review (Glechner, 2023). We did not exclude trials based on the number or frequency of screening tests or based on age groups included. The control was no offer of screening mammograms, but we accepted clinical and self-breast examination in the control group.

#### Types of outcome measures

We included trials whether they reported our pre-specified outcomes or not. All our outcome measures of effects were dichotomous (binary data) and differences are presented as risk ratios (RR) with 95% confidence intervals. Absolute differences are also presented using a denominator that allows direct comparisons between benefits and harms, i.e. in our Summary of Findings tables. We report outcomes at 7, 13 and, when possible, 25 years of follow-up. Overdiagnosis was meassured as the difference in incidence between the screening and control arm at the latest time of follow-up in those trials that did not offer mammography screening to the control group at the end of the intervention phase. This is the same definition as used in another review (UK review, 2012).

##### Primary outcomes

Mortality from breast cancer

Mortality from any cancer

All-cause mortality

Use of surgical interventions

Use of adjuvant therapy

Harms of mammography

##### Secondary outcomes

None

### Search methods for identification of studies

#### Electronic searches

For the, 2023 update of our review, we searched the following databases up to 28 February, 2023:

- PubMed (Appendix 1).
- The Cochrane Central Register of Controlled Trials (CENTRAL) (The Cochrane Library, 2023, Issue 2) (Appendix 2).
- The Cochrane Breast Cancer Group’s Specialised Register. Details of the search strategies used by the CBCG for the identification of studies and the procedure used to code references are outlined in their module (http://onlinelibrary.wiley.com/o/cochrane/clabout/articles/BREASTCA/frame.html). Trials with the keywords ‘mammography’ and ‘screening’ were extracted for consideration.
- The World Health Organization (WHO) International Clinical Trials Registry Platform (ICTRP) search portal (http://apps.who.int/trialsearch/Default.aspx) (Appendix 3).
- Clinicaltrials.gov (http://clinicaltrials.gov/ct2/search) (Appendix 4).

In the original version of the review, we used a very broad search strategy. We searched PubMed with (breast neoplasms[MeSH] OR “breast cancer” OR mammography[MeSH] OR mammograph*) AND (mass screening[MeSH] OR screen*). This search was supplemented with a search on author names in the author field (Alexander F*, Andersson I*, Baines C*, Bjurstam N*, Duffy S*, Fagerberg G*, Frisell J*, Miller AB, Moss S*, Nystrom L*, Shapiro S, Tabar L*). The latest search was done on 22 November, 2012 and 29,222 records were imported into ProCite. Until the, 2009 review, these records were searched for author names, cities and eponyms for the trials; thereafter, all new records were browsed. This very broad search strategy, combined with browsing the titles and reading the abstracts when a paper might be relevant for mammography screening, enabled us to assemble also observational studies of the benefits and harms of screening.

We searched the World Health Organization’s International Clinical Trials Registry Platform (22 November, 2012) with this strategy, for Recruitment Status ALL: (Condition: breast AND (cancer% OR carcinoma% OR neoplas% OR tumour% OR tumor%) AND Intervention: screen OR mass screen%) OR (Condition: breast AND (cancer% OR carcinoma% OR neoplas% OR tumour% OR tumor%) AND Intervention: mammograph%) OR (Condition: breast neoplasm AND Intervention: mammography).

#### Searching other resources

We scanned reference lists and included letters, abstracts, grey literature and unpublished data to retrieve as much relevant information as possible. There were no language restrictions.

### Data collection and analysis

#### Selection of studies

In this updated review, we used Cochrane’s Screen4Me workflow to help assess the search results. Screen4Me comprises three components: known assessments – a service that matches records in the search results to records that have already been screened in Cochrane Crowd and been labelled as an RCT or as Not an RCT; the RCT classifier – a machine learning model that distinguishes RCTs from non-RCTs, and if appropriate, Cochrane Crowd – Cochrane’s citizen science platform where the Crowd help to identify and describe health evidence. For more information about Screen4Me and the evaluations that have been done, please go to the Screen4Me webpage on the Cochrane Information Specialist’s portal: https://community.cochrane.org/organizational-info/resources/resourcesgroups/information-specialists-portal. In addition, more detailed information regarding evaluations of the Screen4Me components can be found in the following publications: Noel-Storr, 2020; Noel-Storr, 2021; Marshall, 2018; Thomas, 2020.

Two authors independently decided which trials to include based on the prestated criteria using Covidence software. Disagreements were resolved by discussion (Figure 1).

**Figure 1.**
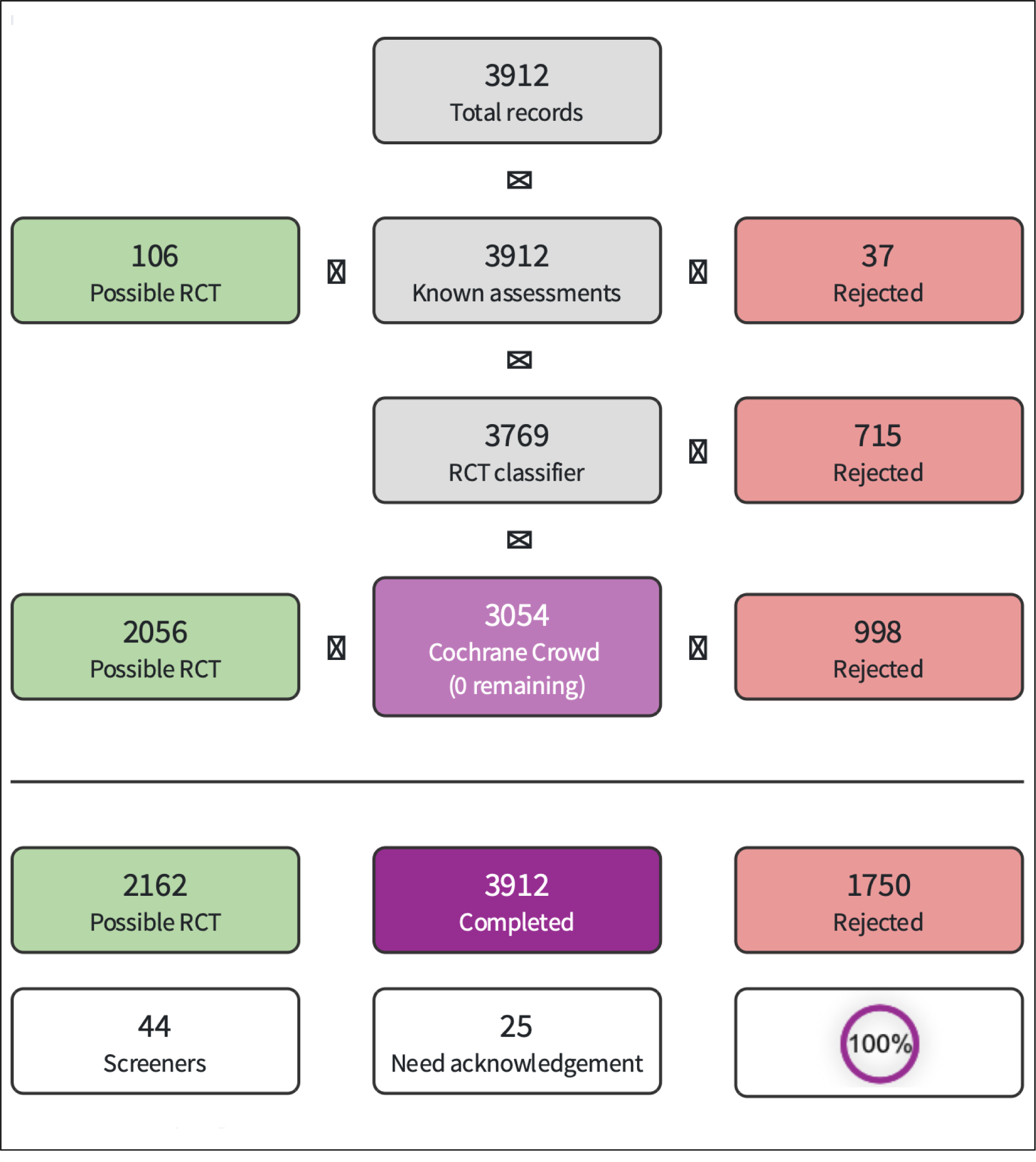
Screen4Me Summary Diagram

#### Data extraction and management

Two authors independently extracted methodological and outcome data; disagreements were resolved by discussion. Extracted data included: number of women randomised; randomisation and blinding procedures; exclusions after randomisation; type of mammography; number of screenings and interval between screenings; attendance rate; introduction of screening in the control group; co-interventions; number of cancers identified; breast cancer mortality; cancer mortality; all-cause mortality; harms of mammography; and use of surgical interventions, chemotherapy, radiotherapy, tamoxifen and other adjuvant therapy.

#### Assessment of risk of bias in included studies

We assessed whether the randomisation was adequate and led to comparable groups, following standard criteria as closely as possible (Higgins, 2008). These included sequence generation and allocation concealment. We also assessed blinding of outcome assessors; incomplete outcome data; selective reporting; and other biases. The risk of bias for each domain was assessed as ‘high’, ‘moderate ‘ or ‘low’. As trials of population screening look for small differences in absolute terms, they are sensitive to bias and we payed particular attention to the randomisation method and possible baseline imbalances. We divided the trials into those with adequate randomisation (individual, centralised randomisation) and those with suboptimal randomisation (cluster randomisation, randomisation by date of birth, or randomisation in other ways that would raise concern about baseline imbalances). According to the Cochrane Handbook (Higgins, 2008), the primary analysis in a systematic review should be based on studies at low risk of bias. We therefore did not combine results from adequately randomised studies with other studies.

#### Measures of treatment effect

Risk ratios and 95% confidence intervals. All included outcomes were binary.

#### Unit of analysis issues

We based our conclusions on the individually randomised trials with adequate randomisation methods. Results from suboptimally randomised trials that used cluster randomisation are presented in separate subgroup analyses and did not form the basis for our conclusions. As the included trials were old, information required to adjust for clustering effects were not available. Given that we present the results from the cluster randomised trials for transparency and completeness, and as they did not influence our conclusions, an adjustment for clustering effects would not have changed them.

#### Dealing with missing data

This was assessed as part of our risk of bias assessment. We contacted the primary investigators to clarify uncertainties and to obtain additional data. We analysed available data only and did not impute missing data.

#### Assessment of heterogeneity

In case of indications of substantial statistical heterogeneity (non-overlapping confidence intervals or I^2^ >70%) we explored possible causes in subgroup analyses. Possible methodological reasons for heterogeneity in terms of adequate versus suboptimal randomisation was also explored in subgroup analyses and using the Cochrane risk of bias tool. Any possible clinical heterogeneity was evaluated through the indirectness domain in our GRADE assessment.

#### Assessment of reporting biases

As trials of population screening tend to be very large, we consider it unlikely that trials were performed but not reported and we have not found indications that such trials exist. The size of the trials also means that small study effects are unlikely and we therefore did not use funnel plots to explore this. Incomplete data assessment and incomplete outcome reporting was considered as part of our risk of bias assessment.

#### Data synthesis

We performed intention-to-treat analyses, when possible, by including all randomised women. A fixed-effect model with the Mantel-Haenszel method was used, and 95% confidence intervals (CI) are presented. Absolute risk are calculated and presented in our Summary of Findings tables.

In the trials with suboptimal randomisation, we could not carry out a proper analysis for all-cause mortality as we did not have access to the necessary data to correct for baseline differences (see ‘Risk of bias in included studies’) but present the available data in the graphs for the sake of completeness. For breast cancer mortality, our estimates are not formally correct because we were unable to adjust for baseline differences since baseline characteristics were not reported for several of the suboptimally randomised trials. However, they turned out to be in close agreement with the estimates and CIs published by the trialists.

We report outcome data at approximately 7, 13, and 24 years, which were the most common follow-up periods in the trial reports and report effect estimates at multiple time points as not all trials presented data after very long follow-up and as some effects could be diluted over time. We present age groups under 50 years of age, 50 years and above; and 70 years and above, which is the age limits that has most often been used by the trialists and in screening programmes.

#### Subgroup analysis and investigation of heterogeneity

Apart from analyses by age groups and the division of the trials according to whether they were adequately or suboptimally randomised, we did not perform subgroup analyses.

#### Sensitivity analysis

We did not do any sensitivity analyses as we had already explored the possible impact of age and randomisation method on the robustness of results in subgroup analyses.

#### Summary of findings and assessment of the certainty of the evidence

We exported results of our meta-analyses to GRADEpro GDT, which presented these including absolute numbers for our calculated risk ratios. We selected effect meassures from the least biased trials to be presented in the Summary of Findings table and presented a table for all age groups combined; for women aged <50 years; aged >50 years; and 70 years and above. We present Summary of Findings tables for various age groups separately as an expansion of breast screening is currently considered in various countries. However, not all outcomes could be assessed for all age groups and the most complete outcome set is reported for the combined asssessment of the intervention for all age groups. We prioritised to present results at longest time of follow up, but as not all trials reported results for total mortality at 25 years, we present this oucome at 13 years of follow-up in the table. In GRADEpro GDT, we performed the GRADE assessment of the certainty of the evidence for each outcome. We assed the domains: risk of bias; inconsistency; indirectness; imprecision; and publication bias.

Data for all our outcomes were available for only one comparison (breast screening in all age groups combined versus no screening). For other comparisons, outcomes for which no data were available are not shown to avoid large empty tables for some age groups.

## Results

### Description of studies

Our process of trial identification is depicted in Figure 1 and Figure 2. A description of the 8 included trials can be found here: Canada, 1980; Canada, 1980a; Canada, 1980b; Edinburgh, 1978; Göteborg, 1982; Göteborg, 1982a; Göteborg, 1982b; Kopparberg, 1977; Malmö, 1976; Malmö II, 1978; New York, 1963; Östergötland, 1978; Stockholm, 1981; Two-County, 1977; UK age trial, 1991. A description of the 3 excluded trial can be found here: Berglund, 2000; Dales, 1979; Singapore, 1994; and a description of the 2 trials awaiting classification can be found here: AgeX Trial; Murillo, 2016.

**Figure 2.**
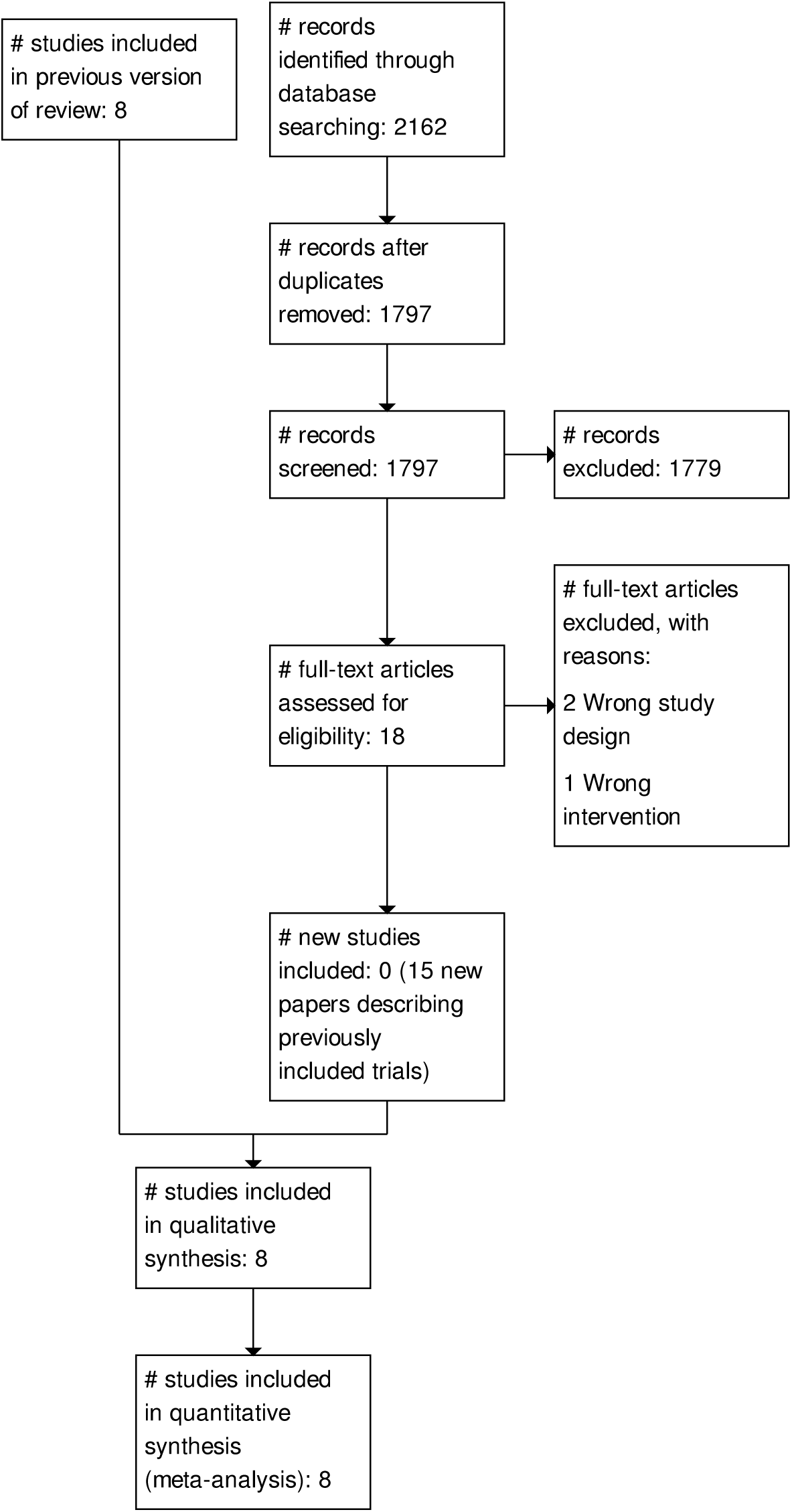
PRISMA flow diagram of review update.

#### Results of the search

In this updated review, after de-duplication, the search identified a total of 3,912 records. In assessing these, we used Cochrane’s Screen4Me workflow to help identify potential reports of randomised trials. The results of the Screen4Me assessment process can be seen in Figure 1. We then assessed the remaining 2,162 records left after Screen4Me. When 365 duplicates were removed,, 1797 records were assessed for eligibility, of which 18 were retrieved in full text. We excluded 1 record because it did not address the relevant intervention and excluded 2 records because the study design did not fulfil our inclusion criteria. We did not identify any new, completed trials compared to our previous update, nor did we find any additional trials through other sources. The results of the searches are presented in the PRISMA flow diagram (Figure 2).

In total, we included 8 studies (8 from the previous review, with 15 new papers describing already included trials identified in the updated search).

The original PubMed search we performed in, 2012 identified 29,222 records.

#### Included studies

We included eight trials (Canada, 1980; Edinburgh, 1978; Göteborg, 1982; Malmö, 1976; New York, 1963; Stockholm, 1981; Two-County, 1977; UK age trial, 1991), which comprised slightly different subtrials. The Canadian trial was actually two trials, one covering the age group 40 to 49 years (Canada, 1980a) and the other 50 to 59 years (Canada, 1980b). The Edinburgh and Malmö trials continued to include women as they passed the lower age limit for entry to the trial, and the Two-County trial had different randomisation ratios in the two counties (Kopparberg, 1977; Östergötland, 1978). Most trials covered the age range 45 to 64 years, but the UK Age trial invited women aged 39 to 41 years to participate. The Canadian trial was the only one in which the women were individually randomised after invitation and informed consent to participate; the others used a variety of procedures based on a prespecified segment of the female population that was randomised to invitation for screening or to a control group.

The number of screening invitations was in the range of four to nine for all trials except the Stockholm and Two-County trials, in which a large fraction were invited for only two or three screenings. In the Two-County trial, the mammographically screened women were encouraged to perform breast self-examinations once a month on a fixed date (Rapport, 1982). This was Swedish policy generally but we do not know for certain whether this was also true for the Göteborg, Malmö and Stockholm trials. Clinical examinations of screened women were performed in New York and Edinburgh. In Canada, in the 40 to 49 year age group, screened women had an annual clinical breast examination whereas control women were examined at the first visit and were taught self-examination for use thereafter. In the 50 to 59 year age group, all women had their breasts clinically examined annually.

The women in the control group were not invited to screening at any point in time in the New York trial, whereas they were invited for screening after 10 to 13 years of follow up in the Edinburgh, Malmö and UK age trials. In the Canadian trial, most of the women in the control group were invited when the trial ended (Baines, 2005). Some women were invited for screening while the trial was still ongoing in the Göteborg, Stockholm and Two-County trials (see ‘Risk of bias in included studies’).

In all trials, women in the control groups were offered usual care. This included mammography on indication, that is for suspected malignancy, with the probable exceptions of the New York trial and the first five years of the Two-County trial.

According to the information we identified, the technical quality of the mammograms and the observer variation were assessed only in the Canadian trial. There are data on diagnostic rates, however, that show that the sensitivity in the trials that followed the New York trial has not consistently improved (Fletcher, 1993; IARC, 2002). Various combinations of one- and two-view mammography were used, i.e. one view mammography was used in the Two-County trial whereas two-view mammography was used in the Canadian trial (see ‘Characteristics of included studies’).

An additional trial in the UK is ongoing (http://www.controlled-trials.com/ISRCTN33292440). This is an age extension, cluster randomised trial, recruiting women aged 47-49 or 71-73 years old, and aiming for a sample size of 3 million women. It started in, 2010 and has not yet reported any results.

A small trial from Colombia where only two women died from breast cancer at the time of reporting is awaiting classification (Murillo, 2016).

#### Excluded studies

We excluded two small trials of several interventions including mammography (Berglund, 2000; Dales, 1979) and a trial involving 166,600 women where the only intervention was a prevalence screen and where exclusions after randomisation occurred only in the screened group; previous cancer at any site was an exclusion criterion and more than, 1500 women were excluded from the screened group, 468 because they had already died (Singapore, 1994).

### Risk of bias in included studies

The trials have been conducted and reported over a long period of time, during which standards for reporting trials have improved. The New York trial, for example, was first reported in, 1966 but crucial details on the randomisation method, exclusions and blinding were not published until 20 years later (Aron, 1986; Shapiro, 1985; Shapiro, 1988). Our risk of bias assessment is depicted in our risk of bias graph (Figure 3) and our risk of bias summary (Figure 4), as well as in our forest plots. Data on use of radiotherapy and chemotherapy in the Kopparberg trial were published 14 years after the main results (Tabar, 1999). Below, we discuss the trial methodology in detail, which is essential reading to understand the controversies surrounding the effects of screening and the often conflicting information presented. The trials are described consecutively by start date.

**Figure 3.**
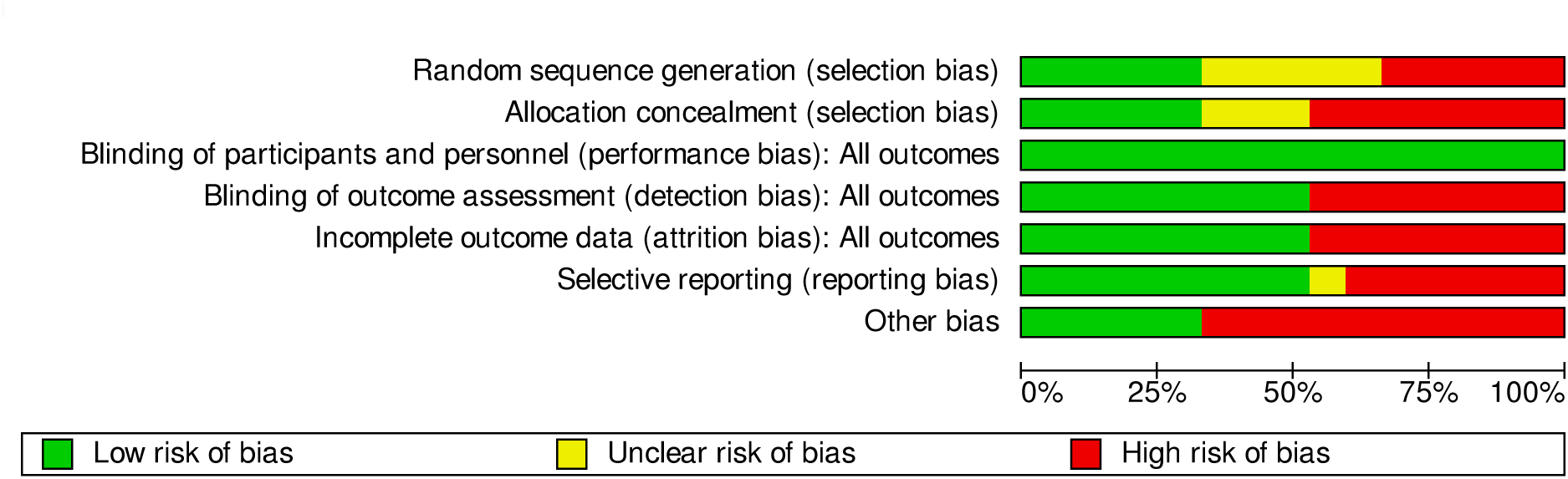

**Figure 4.**
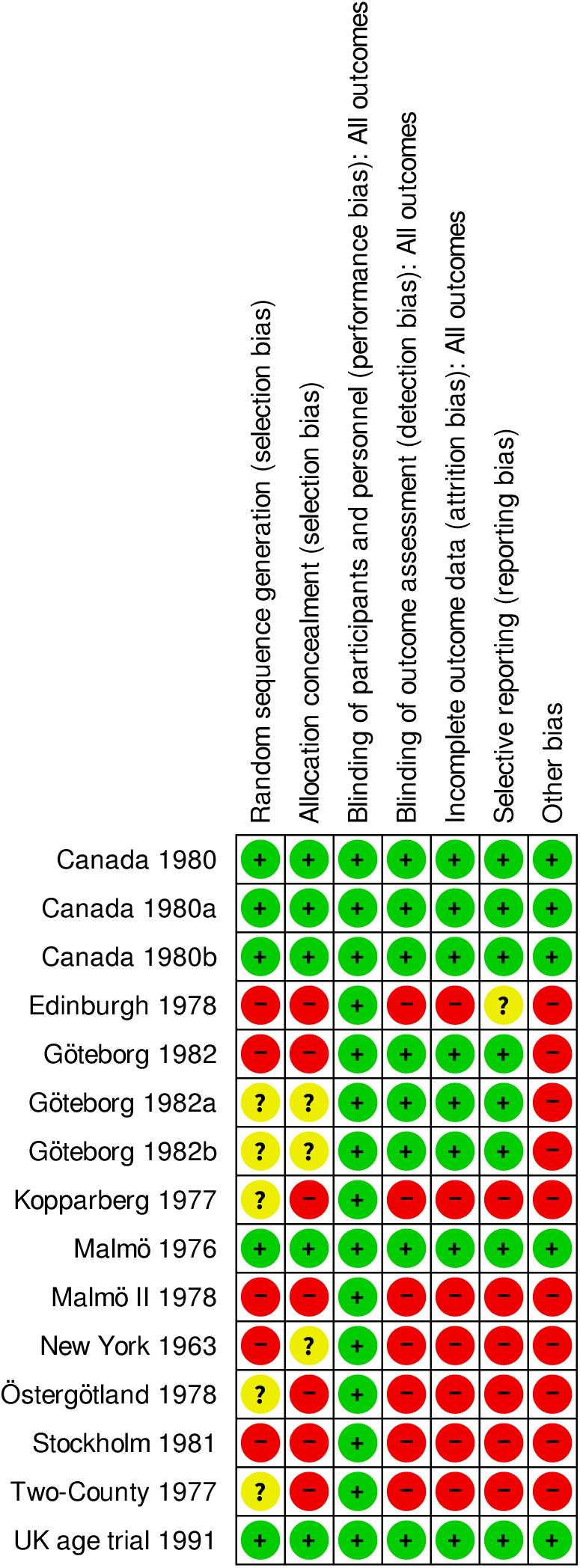

#### The New York trial (New York, 1963)

##### Population studied

The New York trial (also called the Health Insurance Plan (HIP) trial) invited women who were members of an insurance plan and aged 40 to 64 years from December, 1963 to June, 1966. It reported an individual randomisation within pairs matched by age, family size and employment group (Shapiro, 1985). It is not clear whether the randomisation method was adequate; it was described as “alternation” by researchers who contacted one of the trial investigators (Freedman, 2004). The entry date for a woman was the date she was scheduled for the examination (Shapiro, 1966); the matched control was assigned the same date (Shapiro, 1985). The matched pairs method should lead to intervention and control groups of exactly the same size. This is supported by the approximate numbers given in several publications, for example “The women were carefully chosen as 31,000 matched pairs” (Strax, 1973). The largest published exact number of women invited is 31,092 (Fink, 1972).

##### Comparability of groups

Postrandomisation exclusions of women with previous breast cancer occurred but this status “was most completely ascertained for screened women,” whereas women in the control group “were identified through other sources as having had breast cancer diagnosed before their entry dates” (Shapiro, 1988). Using information in the trial reports (Fink, 1972; Shapiro, 1985; Shapiro, 1994), we calculated that 853 (31,092 minus 30,239) women were excluded from the screened group because of previous breast cancer compared with only 336 (31,092 minus 30,756) in the control group. Although it was reported that great care was taken to identify these women, the lead investigator noted that more than 20 years after the trial started some prior breast cancer cases among the controls were unknown to the investigators and those women should have been excluded (Shapiro, 1985a). This creates a bias in favour of screening for all-cause mortality and likely also for breast cancer mortality though the authors have written, without providing data, that ascertainment of cases of previous breast cancer was “nearly perfect” in those women who died from breast cancer (Shapiro, 1988).

It is difficult to evaluate whether there were other baseline differences between the groups. In one paper (Shapiro, 1972) the text described all randomised women and referred to a table that showed baseline differences as percentages but did not provide the numbers upon which the percentages were based. Footnotes explained that some of the data were based on 10% and 20% samples. The table title referred to women entering the trial in, 1964, and not all women as claimed in the text. Assuming that the table title is correct, the data presented in some cases were a, 1964 subgroup of 10% and 20% samples. These resulting samples are therefore too small to study other possible baseline differences than those related to differential exclusion of women with previous breast cancer.

##### Assignment of cause of death

We found no data on the autopsy rate. Assignment of cause of death was unblinded for 72% of the women with breast cancer (Shapiro, 1988). The differential exclusions and unblinded assessments make us question the reliability of the reported breast cancer mortality rates.

##### Likelihood of selection bias

We classified the trial as suboptimally randomised.

#### The Malmö trial (Malmö, 1976)

##### Population studied

This trial recruited women aged 45 to 69 years. Randomisation was carried out by computer within each birth year cohort (Andersson, 1981), dividing a randomly arranged list in the middle (Andersson, 1999a). The first publications noted that 21,242 women were randomised to the screening group and 21,240 to the control group (Andersson, 1980; Andersson, 1981a).

##### Comparability of groups

A later publication reported four more women in the control group (Andersson, 1983) but the main publication (Andersson, 1988) reported only 21,088 women in the study group and 21,195 in the control group. It did not account for the 199 or 203 missing women. The number of missing women was largest in the 45 to 50 years age group (137 from the intervention group and 26 or 27 from the control group), mainly because the, 1929 birth year cohort was recruited by an independent research project that included mammography (Andersson, 2001). The trialists recruited less than the planned 50% of this birth year cohort, but this does not explain why 26 or 27 women were missing from the control group. Exclusion of the, 1929 birth year cohort from analysis changes the risk ratio for death from breast cancer by only 0.01 (Andersson, 2001). For 17 of the 25 birth year cohorts, the size of the study and control groups were identical or differed by only one, as expected. The largest difference in the other eight cohorts, apart from the, 1929 one, was 25 fewer women than expected in the study group for the, 1921 cohort (Nyström, 2002). Thus, the authors of a meta-analysis of the Swedish trials did not report on all randomised women in Malmö (Nyström, 2002).

The date of entry into the trial was defined differently for the two groups. For the mammography group it was the date of invitation (Andersson, 1988), and the midpoint of these dates for each birth year cohort defined the date of entry for women in the control group (Andersson, 2000). Enrolment began in October, 1976 (Andersson, 2000) and ended in September, 1978 (Andersson, 1988). It is not clear whether screening of the control group began in December, 1990 (Nyström, 2000) or in October, 1992 (Nyström, 2002). Most women in the control group were never screened (Nyström, 2002). We calculated the interval between when screening started in the study group and in the control group (the intervention contrast) to be 19 years (Nyström, 2002). In the meta-analyses of the Swedish trials, breast cancer cases diagnosed before randomisation were explicitly excluded, further reducing the screened group by 393 and the control group by 412 (Nyström, 1993); in total 86 more women were excluded from the screened group than the control group. Baseline data on age were not significantly different in the screened group and the control group (Gøtzsche, 2000a).

##### Assignment of cause of death

The autopsy rate for breast cancer cases as presented in the main publication for this trial (Andersson, 1988) was high at 76%, but it was halved from, 1985 to, 1997 (Andersson, 2000). Cause-of-death assessments were blinded up to, 1988 (Andersson, 2000).

##### Likelihood of selection bias

We classified the trial as adequately randomised.

#### The Malmö II trial (Malmö II, 1978)

##### Population studied

This was an extension of the Malmö trial, called MMST II. Women who reached the age of 45 years were enrolled between September, 1978 and November, 1990; screening of the control group began in September, 1991 (Nyström, 2000). The long enrolment period gives an average estimated intervention contrast of eight years. Although the entry criterion for age was stated to be 45 years, the trialists included, 6780 women aged 40 to 44 (Nyström, 2002).

##### Comparability of groups

The MMST II trial has been published only in brief (Andersson, 1997). We therefore cannot check whether there were differential postrandomisation exclusions. If the same procedure as in the Malmö trial had been followed, the sizes of the study and control group cohorts should not differ by more than one. However, the group size differed more for seven of the 13 birth year cohorts (Nyström, 2002). The reported numbers in the individual cohorts do not add up to the reported totals, but to 28 fewer in the study group and 28 more in the control group. Because of an administrative error, the entire, 1934 birth year cohort was invited for screening (Andersson, 1999b). If this cohort is excluded, there is still a gross imbalance with, 5724 women in the study group and only, 5289 in the control group, for those aged 45 to 49 years (P = 0.00004, Poisson analysis). In total, there were, 9581 and, 8212 women in the analyses, respectively (Nyström, 2002).

This trial was neither included nor mentioned in the, 1993 meta-analysis of the Swedish trials (Nyström, 1993). The lead investigator informed us that it was not conducted according to a formal protocol (Andersson, 1999b), whereas the most recent meta-analysis reported that the trial was conducted with the same protocol as the older part of the trial (Nyström, 2002). When the breast cancer mortality rate in the screening group is plotted against the control group rate for eight trials, with data from younger women, the Malmö II trial is a clear outlier (Berry, 1998).

##### Assignment of cause of death

An official registry was used for cause-of-death assessments.

##### Likelihood of selection bias

We classified the trial as suboptimally randomised.

#### The Two-County trial (Kopparberg, 1977; Two-County, 1977; Östergötland, 1978)

##### Population studied

This trial recruited women 40 years of age and over in Kopparberg and Östergötland; the two subtrials were age-matched and cluster randomised (21 and 24 clusters, respectively). The selection of clusters was stratified to ensure an even distribution between the two groups with respect to residency (urban or rural), socioeconomic factors and size (Kopparberg, 1977; Tabar, 1979; Östergötland, 1978). The randomisation process and the definition of the date of entry have been inconsistently described; and some women were only 38 years of age, below the inclusion criterion (Nyström, 2002). According to the first publications, random allocation of the women in each community block took place three to four weeks before screening started (Fagerberg, 1985); all women from a given block entered the trial at the same time and this date was the date of randomisation (Tabar, 1985).

However, it has also been described that a public notary allocated the clusters in Östergötland by tossing a coin (Nyström, 2000) while witnesses were present (Fagerberg, personal communication,, 1999). We have been unable to find any detailed description of the randomisation in Kopparberg but found a recent description for the whole trial: “Randomisation was by traditional mechanical methods and took place under the supervision of the trial statistician” (Duffy, 2003). Thus it is not clear whether the randomisation was carried out on one occasion or whether it took place over several years.

Women were invited to their first screening from October, 1977 to January, 1980 in Kopparberg (Tabar, 1981). The cohorts in Östergötland were defined between May, 1978 and March, 1981. It is not clear how many women were randomised and reported numbers vary considerably, both for numbers randomised (Table 1) and for numbers of breast cancer deaths, despite similar follow up (Gøtzsche, 2004). Documentation of baseline comparability was called for in, 1988 (Andersson, 1988a) but it appears not to have been published. Since the randomisation was stratified after socioeconomic factors (Tabar, 1991), baseline data potentially affecting mortality should exist.

**Table 1.** Examples of varying numbers of women in the Swedish trials.

| Study | Age range | Study group | Control group | Reference |
| --- | --- | --- | --- | --- |
| Malmö | 40-74 | 21242 | 21240 | Andersson 1980 |
|  | 40-74 | 21242 | 21244 | Andersson 1983 |
|  | 40-74 | 21088 | 21195 | Andersson 1988 |
| Kopparberg | total | 47389 | 22658 | Socialstyrelsen 1985 |
|  | 40-74 | 39051 | 18846 | Tabar 1985 |
|  | 40-74 | 38589 | 18582 | Tabar 1989 |
|  | 40-74 | 38562 | 18478 | Nyström 1993 |
|  | 40-74 | 38589 | 18582 | Tabar 1995 |
|  | 40-74 | 38568 | 18479 | Nyström 2000 |
|  | 40-74 | 38588 | 18582 | Nixon 2000 |
|  | 40-74 | data not available | data not available | Nyström 2002 |
|  | 40-49 | 9625 | 5053 | Tabar 1988 |
|  | 40-49 | data not available | data not available | Nyström 1993a |
|  | 40-49 | 9582 | 5031 | Tabar 1995 |
|  | 40-49 | 9650 | 5009 | Nyström 1997 |
| Östergötland | total | 47001 | 45933 | Socialstyrelsen 1985 |
|  | 40-74 | 39034 | 37936 | Tabar 1985 |
|  | 40-74 | 38491 | 37403 | Tabar 1989 |
|  | 40-74 | 38405 | 37145 | Nyström 1993 |
|  | 40-74 | 38491 | 37403 | Tabar 1995 |
|  | 40-74 | 38942 | 37675 | Nyström 2000 |
|  | 40-74 | 39105 | 37858 | Nixon 2000 |
|  | 40-74 | 38942 | 37675 | Nyström 2002 |
|  | 40-49 | 10312 | 10625 | Tabar 1988 |
|  | 40-49 | data not available | data not available | Nyström 1993a |
|  | 40-49 | 10262 | 10573 | Tabar 1995 |
|  | 40-49 | 10240 | 10411 | Nyström 1997 |
| Stockholm | 40-64 | 40318 | 19943 | Frisell 1989a |
|  | 40-65 (sic) | 38525 | 20651 | Nyström 1993 |
|  | 40-64 | 40318 | 19943 | Frisell 1997 |
|  | 40-69 | 39139 | 20978 | Nyström 2000 |
|  | 40-49 | data not available | data not available | Nyström 1993a |
|  | 40-49 | 14842 | 7103 | Frisell 1997 |
|  | 40-49 | 14185 | 7985 | Nyström 1997 |
|  | 40-49 | 14303 | 8021 | Nyström 2002 |
| Göteborg | 40-59 | 20724 | 28809 | Nyström 1993 |
|  | 39-59 | 21650 | 29961 | Bjurstam 1997a |
|  | 40-59 | 21000 | 29200 | Nyström 2000 |
|  | 40-49 | 10821 | 13101 | Nyström 1993a |
|  | 39-49 | 11724 | 14217 | Bjurstam 1997 |
|  | 40-49 | 10888 | 13203 | Nyström 2002 |

##### Comparability of groups

The randomisation procedure seems to have led to non-comparable groups. First, breast cancer mortality in the control group was almost twice as high in Kopparberg compared to Östergötland (0.0021 versus 0.0012, P = 0.02). This was not apparent from the tabulated data (Tabar, 1985). The published graphs are also potentially misleading; although adjacent mortality curves look much the same the two y-axes are differently scaled (Tabar, 1995). Second, in Kopparberg more women in the control group were diagnosed with breast cancer before entry to the trial than in the study group. How the diagnostic information was obtained was not described (Tabar, 1989) and the number of women excluded for this reason was not stated, but can be calculated by comparing two tables (Tabar, 1985; Tabar, 1989). More women were excluded from the control group than from the study group (P = 0.03); most of the imbalance occurred in the age group 60 to 69 years (P = 0.007). In Östergötland, numbers of exclusions were very similar, 1.40% versus 1.39%. Third, age-matching was reported (Tabar, 1979; Tabar, 1981; Tabar, 1985a) but study group women were on average five months older (Nixon, 2000), which is a small bias against screening.

We were unable to ascertain when systematic screening of the control group started. The available information is conflicting and the range of the discrepancies amounts to three years for both counties (Arnesson, 1995; Duffy, 2003; Nyström, 1993, ; Nyström, 2000; Nyström, 2002; Rapport, 1982; Tabar, 1979; Tabar, 1985; Tabar, 1992). It seems most likely that screening of the control group in Kopparberg started in, 1982, in accordance with the trial protocol (Rapport, 1982) and a doctoral thesis (Nyström, 2000). In this case, the impression conveyed in the main publication for the trial that screening was offered to the control group after publication of the results in April, 1985 is incorrect (Tabar, 1985; Tabar, 1992). In the protocol, a five-year intervention period was planned but with a stopping rule based on statistical significance testing every six months (Rapport, 1982). The trial publications did not mention the repeated looks at the data (Tabar, 1985). We estimated an intervention contrast of five years for Kopparberg and eight years for Östergötland. A valid comparison of benefits and harms of screening should be confined to the period prior to screening of the control group.

No information is available from the primary author of this trial (Atterstam, 1999; Prorok, 2000; Tabar, 2000a). We have not received information from Nyström either on the missing account of the randomisation process in Kopparberg, or from the Swedish National Board of Health (Socialstyrelsen), which funded the trial.

##### Assignment of cause of death

The autopsy rate was 36% (Projektgruppen, 1985). According to an investigator involved with the trial (Crewdson, 2002), other Swedish trialists (Nyström, 2002), and an IARC report (IARC, 2002), cause-of-death assessments were not blind. This has been disputed by the lead investigator of the trial (Tabar, 2002). In a meta-analysis of the Swedish trials, a blinded independent endpoint committee reassessed the death classifications (Nyström, 1993).

##### Likelihood of selection bias

We classified the trial as suboptimally randomised and likely to be biased.

#### The Edinburgh trial (Edinburgh, 1978)

##### Population studied

This trial used cluster randomisation with about 87 clusters (the number varies in different reports); the age group was 45 to 64 years. Coded general practices were stratified by size and allocated by manual application of random numbers. In one district, at least three of the 15 practices initially randomised to the screening group later changed allocation status, and at least four others were added (Alexander, 1989). Two of these practices were unintentionally told the wrong group, and three changed allocation group because of “statistical considerations” (Roberts, 1984). One practice was included in the follow up even though it was a pilot screening practice that did not participate in the randomisation (Roberts, 1990). The trialists have conducted replicate analyses with these women removed (Alexander, 2000) but as far as we know the data have not been published.

##### Comparability of groups

Doubts about the randomisation process were raised by the trialists (Alexander, 1989), supported by baseline differences: 26% of the women in the control group and 53% in the study group belonged to the highest socioeconomic level (Alexander, 1994), and mammographic screening was associated with an unlikely 26% reduction in cardiovascular mortality (Alexander, 1989). Entry dates were defined differently. In most practices the entry date was the date the invitation letter was issued; for women in hospital it was the date their names appeared on a list sent to their general practitioner. The entry date for five practices was not defined. In the control group, the entry date was the date the physician’s practice was indexed. Before entry, the general practitioners in the screening practices had to decide whether each woman would be suitable for invitation to screening. Physicians in the control practices decided whether each woman would be eligible to receive a leaflet about breast self-examination (Roberts, 1984). The eligibility criteria were thus broader for the control group and the entry dates seem to be earlier. Practices were enrolled one at a time over a period of 2.5 years, from, 1979 to, 1981 (Alexander, 1989). Women turning 45 years of age and women moving into the city were enrolled on an ongoing basis (Roberts, 1984). Recruitment of the control group began in the 10th year of follow up (Alexander, 1994). The exclusion procedures were different in the study and control groups (Chamberlain, 1981; Roberts, 1984) and 338 versus 177 women were excluded because of prior breast cancer (Alexander, 1994).

##### Likelihood of selection bias

This trial was not adequately randomised and was so biased that it cannot provide reliable data. We have therefore shown its results in a separate graph, for completeness only.

#### The Canadian trial (Canada, 1980; Canada, 1980a; Canada, 1980b)

##### Population studied

Women aged 40 to 59 years were individually randomised after invitation and giving informed consent. Their names were entered successively on allocation lists, where the intervention was prespecified on each line. An independent review of ways in which the randomisation could have been subverted uncovered no evidence of this (Bailar, 1997). Enrolment took place from January, 1980 to March, 1985 (Canada, 1980a).

##### Comparability of groups

Fifty-nine women in the age group 40 to 49 years and 54 in the age group 50 to 59 years were excluded after randomisation (Miller, 2000; Miller, 2002); none were excluded because of previous breast cancer. The comparison groups were nearly identical in size (25,214 versus 25,216 aged 40 to 49 years; and 19,711 versus 19,694 aged 50 to 59 years), and were similar at baseline for age and nine other factors of potential prognostic importance (Baines, 1994; Canada, 1980; Canada, 1980a; Canada, 1980b; Miller, 2000; Miller, 2002). There were more small node-positive cancers at baseline in the screened group than in the control group among women aged 40 to 49 years, but this is a post-hoc subgroup finding which is probably a result of the intervention (Baines, 1995; Baines, 1997; Canada, 1980). Several women with positive nodes were probably unrecognised in the control group (Miller, 1997a). This is supported by the fact that 47% of women with node-negative cancer in the usual care group died of breast cancer compared with 28% in the mammography group (Miller, 1997). Exclusion of the deaths caused by these cancers did not change the result (Baines, 1995; Baines, 1997; Canada, 1980).

##### Assignment of cause of death

The autopsy rate was low, 6% (Baines, 2001). Cause-of-death assessments were blinded for women with diagnosed breast cancer and for other possible breast cancer deaths, for follow up after seven years. For follow up after 13 years, death certificates were used in a minority of cases as some hospitals refused to release clinical records (Miller, 2000; Miller, 2002).

##### Likelihood of selection bias

We classified the trial as adequately randomised.

#### The Stockholm trial (Stockholm, 1981)

##### Population studied

In this trial, women were invited for screening if they were aged 40 to 64 years in, 1981 (born, 1917 to, 1941) and were born on days 1 to 10 in a month, or if they were aged 40 to 64 years in, 1982 (born, 1918 to, 1942) and were born on days 21 to 30 in a month (Frisell, 1986). Similarly, there were two groups of controls but since they were all born on days 11 to 20 in a month, most women served as controls twice (those born in, 1918 to, 1941).

Invitations were sent successively by ascending order of birth date (Frisell, 1989). The date of entry was the date of invitation (Frisell, 1991). Enrolment of the first cohort began in March, 1981 and ended in April, 1982; enrolment of the second cohort began in April, 1982 and ended in May, 1983 (Frisell, 2000a).

##### Comparability of groups

Since the control women born in, 1918 to, 1941 served as controls for both subtrials (Frisell, 1989a; Frisell, 2000b) they should have two entry dates, approximately one year apart, but this was not described. According to the matching there should have been a similar number of women in the screened and control groups in each subtrial, but we found an imbalance in the second subtrial (P = 0.01, Poisson analysis) with 508 more women belonging to the screened group than to the control group (Frisell, 1991). Furthermore, in the time period where 19,507 women born from, 1918 to, 1942 were invited to screening, only 929 women, all born in, 1942, were included in the control group (Nyström, 2002).

The reported numbers of women in the various subgroups are inconsistent, as are the numbers reported to us in personal communications (Frisell, 2000a; Frisell, 2000b). Because of the problems related to timing and the overlap of the two control groups, results from the two subtrials were not independent, and the estimates cannot be pooled without correction for dependence. It is not clear how these difficulties were handled in the trialists’ analysis (Frisell, 1991) or in the Swedish meta-analyses (Nyström, 1993; Nyström, 2000; Nyström, 2002).

The first trial report did not describe any women excluded after randomisation; only breast cancer cases identified during the intervention period were followed up to ascertain breast cancer deaths (Frisell, 1991). Exclusions occurred in later publications but no numbers were given (Frisell, 1997; Nyström, 1993; Nyström, 2000) and the numbers we have received in personal communications have been inconsistent (Frisell, 2000a; Frisell, 2000b).

Of those attending the first screening, 25% had had a mammogram in the two previous years (Frisell, 1989a). Information on screening of the control group varied. A meta-analysis noted that a few women were screened after three years and most after four years (Nyström, 1993), a doctoral thesis stated that the controls were invited for screening from October, 1985 (Nyström, 2000), and the trialists noted that they were invited during, 1986 (Frisell, 1989a; Frisell, 1991). We estimated an intervention contrast of four years. A valid comparison of benefits and harms of screening should be restricted to this period (Frisell, 1991).

##### Assignment of cause of death

It is not stated whether cause-of-death assessments were blinded for this initial period. The autopsy rate was 22% (Nyström, 2000).

##### Likelihood of selection bias

We classified the trial as suboptimally randomised.

#### The Göteborg trial (Göteborg, 1982)

##### Population studied

This trial included women aged 39 to 59 years. Birth year cohorts were randomised by the city municipality’s computer department with the ratio between study group and control group adjusted according to the capacity of the screening unit (Bjurstam, 2000; Nyström, 2002). The randomisation was by cluster based on date of birth in the, 1923 to, 1935 cohorts, and by individual birth date for the, 1936 to, 1944 cohorts (Bjurstam, 1997).

##### Comparability of groups

We found baseline data only on age, and only for those aged 39 to 49 years. Since the allocation ratios were irregular due to limited screening capacity (Bjurstam, 2016), we could not assess the comparability of groups and adequacy of randomisation, but the randomisation process is described as by day-of-birth-cluster up to November, 1983, after which individual randomisation was used (Bjurstam, 2016). The randomisation ratios were most extreme for the oldest and the youngest birth-year cohorts randomised in clusters; for, 1923, there were 2.0 times as many women in the control group as in the study group, whereas for, 1935 there were only 1.1 times as many. Since breast cancer mortality increases with age, this bias favoured screening and can be adjusted for only by comparing the results within each birth-year cohort before they are pooled (Bjurstam, 2003).

This was the only trial to show a difference in total mortality at 13 years of follow-up (RR 0.89, CI 0.83 to 0.95) (Analysis 1.10). As the trial was much underpowered to show such a difference, this result lends support to our assessment that the randomisation was suboptimal and led to baseline differences for prognostic factors important for survival. Furthermore, fewer breast cancers were identified in the screening arm than in the control arm when the control arm had been screened once at the end of the trial period (incidence rate ratio for women 39 to 59: 0.90). This can be calculated from data presented in a table in (Bjurstam, 2016). For breast screening to reduce disease specific mortality, the requisite advancement of time of diagnosis means there must be more cancers detected in the screening arm than the control arm. The difference was driven by women aged 39 to 49 (incidence rate ratio: 0.82) whereas the incidence was similar between groups in women aged 50 to 59 years (incidence rate ratio: 0.99). As the disease specific mortality difference in the trial was driven by the younger age group as well (RR 0.60 vs 0.82 (ns))(Bjurstam, 2016), the apparent benefit of breast screening in this trial could be explained by baseline differences.

Entry dates were not defined but the birth year cohorts were randomised one at a time, beginning with the, 1923 cohort in December, 1982 and ending in April, 1984 with the, 1944 cohort. A similar proportion of women were excluded from the study and control groups, 254 (1.2%) and 357 (1.2%), because of previous breast cancer (Bjurstam, 2003). Information on screening of the control group varied, ranging from three to seven years after randomisation (Bjurstam, 1997; Bjurstam, 2003; Nyström, 1993, figure; Nyström, 2000). We estimated an intervention contrast of five years. A valid comparison of benefits and harms of screening should be confined to this period.

##### Assignment of cause of death

The autopsy rate was 31% (Nyström, 2000). Cause-of-death assessments were blinded.

##### Likelihood of selection bias

We classified the trial as suboptimally randomised.

#### The UK age trial (UK age trial, 1991)

##### Population studied

This trial included women aged 39 to 41 years who were randomised individually between, 1991 and, 1997 to an intervention group or a control group, in a ratio of 1:2. Women in the control group received no information about the trial. The trial was undertaken in 23 breast-screening units in England, Wales, and Scotland. Women were identified from lists of patients from general practitioners held on local Health Authority databases and randomisation was carried out stratified by practice. Prior to this, the general practitioners could remove women with previous breast cancer and others deemed inappropriate to invite for screening. From, 1992 onwards the allocations were carried out on the Health Authority computer system with specifically written software. Before this, for women in three early centres, random numbers generated from the coordinating centre computer were applied to the lists.

##### Comparability of groups

We found baseline data only on age; the mean age was 40.38 and 40.39 years, respectively.

Thirty and 51 women (0.05%) were excluded from analysis for similar reasons in the two groups. The intervention contrast was 10 years. A valid comparison of benefits and harms of screening should be confined to this period.

##### Assignment of cause of death

There was no information on autopsy rate; information on cause of death was obtained from the central register of the National Health Service.

##### Likelihood of selection bias

We classified the trial as adequately randomised.

#### Sources of data used for the meta-analyses

Deaths ascribed to breast cancer: Alexander, 1999; Andersson, 1988; Bjurstam, 1997; Bjurstam, 2003; Duffy, 2020a; Frisell, 1997; Habbema, 1986; Miller, 1992a; Miller, 1992b; Miller, 2000; Miller, 2002; Miller, 2014; Moss, 2006; Moss, 2015; Nyström, 1993; Nyström, 1993a; Nyström, 2002; Roberts, 1990; Shapiro, 1977; Shapiro, 1982; Tabar, 1988; Tabar, 1995.

Mortality among breast cancer patients: Tabar, 1988.

Deaths ascribed to cancer, all patients: Andersson, 1988; Aron, 1986; Duffy, 2020a; Miller, 2000; Miller, 2002; Shapiro, 1988; Tabar, 1988.

All-cause mortality: Andersson, 1988; Aron, 1986; Bjurstam, 1997; Duffy, 2020a; Miller, 1992a; Miller, 1992b; Miller, 2000; Miller, 2002; Moss, 2006; Nyström, 2000; Nyström, 2002; Projektgruppen, 1985; Roberts, 1990; Shapiro, 1977; Tabar, 1989.

Mastectomies and lumpectomies: Andersson, 1988; Frisell, 1986; Frisell, 1989a; Miller, 1993; Shapiro, 1972; Tabar, 1999.

Radiotherapy: Andersson, 1988; Benjamin, 1996; Shapiro, 1972; Tabar, 1999. Chemotherapy and hormone therapy: Andersson, 1988; Tabar, 1999.

Number of cancers: Andersson, 1988; Baines, 2016; Bjurstam, 1997; Frisell, 1989a; Miller, 1993; Moss, 2005; Tabar, 1991.

### Allocation

We classified three trials as adequately randomised (Canada, Malmö and UK age trial) and four as suboptimally randomised (Göteborg, New York, Stockholm, Two-County), as was also the extension of the Malmö trial, MMST II. One trial (Edinburgh) was not adequately randomised and cannot provide reliable data.

### Blinding

We classified three trials as having low risk of bias for cause of death assessment (Canada, Malmö and UK age trial) and four trials as having high risk of bias due to lack of blinded cause of death assessment (Göteborg, New York, Stockholm, and Two-County).

### Incomplete outcome data

We classified four trials as having low risk of bias due to incomplete reporting (Canada, Malmö, Göteborg and UK age trial) and three as having high risk (New York, Stockholm, Two-County).

### Selective reporting

We classified four trials as having low risk of bias due to selective reporting (Canada, Malmö, Göteborg and UK age trial) and three as having high risk (New York, Stockholm, Two-County).

### Other potential sources of bias

We classified three trials as having low risk of bias due to other reasons (Canada, Malmö, and UK age trial) and four as having high risk (New York, Stockholm, Göteborg, and Two-County).

## Effects of interventions

Eight trials provided data. We classified three trials as adequately randomised (Canada, Malmö and UK age trial) and four as suboptimally randomised (Göteborg, New York, Stockholm, Two-County), as was also the extension of the Malmö trial, MMST II. One trial (Edinburgh) was assessed as being too unreliable to provide reliable data due to substantial baseline imbalances and it is accordingly excluded from other key reviews (UK review, 2012); we have therefore only shown its results for completeness, in a separate graph (Analysis 1.22). As the short-term results from the UK Age trial were obtained after a mean follow up of 10.7 years, we included them in the results both after 7 and after 13 years(Analysis 1.1; Analysis 1.2). The adequately randomised trials provided 40% of the breast cancer deaths after 13 years (Analysis 1.2). The effects of the intervention for various age groups and times of follow-up, as well as our GRADE assessments for primary and secondary outcomes, are summarized in our Summary of Findings tables here: Summary of findings table 1; Summary of findings table 2; Summary of findings table 3; Summary of findings table 4.

### Deaths ascribed to breast cancer

We judged assignment of breast cancer mortality to be unreliable and biased in favour of screening (see above and ‘Discussion’), but included this outcome because it was the main focus in all trials.

The three adequately randomised trials did not find an effect of screening on deaths ascribed to breast cancer, risk ratio (RR) 0.93 (95% CI 0.79 to 1.09) after 7 years; RR 0.90 (95% CI 0.79 to 1.02) after 13 years; and RR 0.95 (95% CI 0.86 to 1.04) after 22 years (data available only for the Canadian and UK age trials)(Analysis 1.1; Analysis 1.2; Analysis 1.7). The four suboptimally randomised trials found a beneficial effect: RR 0.71 (95% CI 0.61 to 0.83) after 7 years and RR 0.75 (95% CI 0.67 to 0.83) after 13 years (Analysis 1.1; Analysis 1.2).

The adequately randomised trials did not find an effect of screening on deaths ascribed to breast cancer in the youngest age group (under 50 years of age at randomisation except for Malmö for which the limit was 55 years): RR 0.94 (95% CI 0.78 to 1.14) after 7 years and RR 0.87 (95% CI 0.73 to 1.03) after 13 years (Analysis 1.3; Analysis 1.5). The suboptimally randomised trials found an RR of 0.81 (95% CI 0.63 to 1.05) after 7 years and RR of 0.80 (95% CI 0.64 to 0.98) after 13 years (Analysis 1.3; Analysis 1.5). For women aged >50 years, the estimates for the adequately randomised trials were RR 0.88 (95% CI 0.64 to 1.20) and RR 0.94 (95% CI 0.77 to 1.15), respectively; for suboptimally randomised trials they were RR 0.67 (95% CI 0.56 to 0.81) and RR 0.70 (95% CI 0.62 to 0.80), respectively (Analysis 1.4; Analysis 1.6).

Only the Two-County trial included women aged 70 years and above and reported results. It was not possible to provide reliable estimates of effect for this age group.

### Deaths ascribed to any cancer

The adequately randomised trials did not find an effect of screening on deaths ascribed to any cancer, including breast cancer; RR 1.00, 95% CI 0.96 to 1.04; the follow up was 10.5 years for Canada, 9 years for Malmö and 23 years for the UK age trial (Analysis 1.8). The suboptimally randomised trials did not provide reliable estimates of total cancer mortality (see above); the estimate for the two suboptimally randomised trials that provided data (New York and Two-County trials) was RR 0.99 (95% CI 0.93 to 1.06)(Analysis 1.8).

### All-cause mortality

All-cause mortality was not reduced; RR 0.98, 95% CI 0.94 to 1.03 after 7 years; RR 0.99, 95% CI 0.95 to 1.03 after 13 years; and RR 1.01, 95% CI 0.99 to 1.04 after 22 years (data only available for the Canadian and UK Age trials)(Analysis 1.9; Analysis 1.10; Analysis 1.15). The suboptimally randomised trials did not provide reliable estimates of the effects on all-cause mortality (see ‘Risk of bias in included studies’ and ‘Discussion’) and the reported effects were heterogeneous (P = 0.03 after 7 years; P = 0.001 after 13 years). For completeness, their mortality estimates are shown in the graphs but collectively, they did not show a difference either (Analysis 1.10). For women under age 50 years, see (Analysis 1.11; Analysis 1.13); for women over age 50 years, see (Analysis 1.12; Analysis 1.14).

### Number of cancers (overdiagnosis)

More women were diagnosed with breast cancer in the screened group in the adequately randomised trials that did not systematically screen women in the control group after the intervention phase (RR 1.25, CI 1.18 to 1.34) (Analysis 1.23). Systematic screening offered to the control group at the end of the intervention phase means overdiagnosis cannot be reliably assessed in the remaining trials.

### Surgery

More breast operations (mastectomies plus lumpectomies) were performed in the study groups than in the control groups: RR 1.31 (95% CI 1.22 to 1.42) for the adequately randomised trials; RR 1.42 (95% CI 1.26 to 1.61) for the suboptimally randomised trials before systematic screening in the control group started (data were available only for Kopparberg and Stockholm)(Analysis 1.16). The increased surgery rate could not be explained by the excess of detected tumours at the first screen but seemed to persist, as the mean follow up was seven years for Canada and nine years for Malmö. For Stockholm, the reported data after five years had been transformed according to the smaller size of the control group (Frisell, 1989a). We recorrected and found that also for this trial the excess of surgery persisted (RR 1.37 after first round; RR 1.48 after five years).

The number of mastectomies (excluding partial mastectomies, quadrantectomies and lumpectomies) was also increased: RR 1.20 (95% CI 1.08 to 1.32) for the adequately randomised trials; RR 1.21 (95% CI 1.06 to 1.38) for the suboptimally randomised trials (Analysis 1.17).

### Other adjuvant therapy

We found little information on other adjuvant therapy. It differed substantially for two of the Swedish trials even though they were carried out at the same time. Chemotherapy was given to only 7% of the breast cancer patients in Malmö but to 31% in Kopparberg before the control group was screened (Analysis 1.19). Conversely, hormone therapy was given to 17% in Malmö, and to 2% in Kopparberg (Analysis 1.20). Information exists from Kopparberg on therapeutic adjuvant therapy given over the years but has not been published (Tabar, 1999).

### Radiotherapy

More women received radiotherapy in the study groups: RR 1.24 (95% CI 1.04 to 1.49) for Malmö after nine years; and RR 1.40 (95% CI 1.17 to 1.69) for Kopparberg before the control group screen (Analysis 1.18).

### Harms

We found no comparative data on psychological morbidity. Duration of sick leave and mobility of the shoulder were recorded in the Two-County trial (Rapport, 1982) but have not been reported.

## Discussion

### Summary of main results

The decision to embark on the screening programmes was made mainly because of the positive results in the New York and Two-County trials (Forrest report, 1986). Policy makers and many scientists believed that the benefit of screening was well documented. However, information essential to judging the reliability of the trials was often unpublished or published only in Swedish, in theses, letters, conference reports, reviews, or in journals that are not widely read and with titles and abstracts that did not indicate that important data were described.

Furthermore, the harms of screening received very little attention.

#### Breast cancer mortality

The main focus in the screening trials was breast cancer mortality, as very large trials are needed to assess the effect on all-cause mortality. We cannot assume, however, that a beneficial effect on breast cancer mortality can be translated into improved overall survival. First, screening may increase mortality because of overdiagnosis and the increased use of radiotherapy. A meta-analysis predicted that overall, radiotherapy is beneficial for women at high risk of local recurrence. However, it is harmful for women at particularly low risk such as those who have their cancers found by screening and those who are overdiagnosed. This is primarily because of damage to the coronary arteries and development of heart failure resulting from at least some types of radiotherapy (EBCTCG, 2000) and because radiotherapy causes lung cancer. A meta-analysis of radiotherapy showed that there was a 27% excess mortality from heart disease and a 78% excess mortality from lung cancer (EBCTCG, 2005a). This excess mortality becomes important when many healthy women are overdiagnosed, even if radiotherapy has been improved and harms reduced since the trials.

Second, assessment of cause of death is susceptible to bias. The authors of the Two-County trial assessed cause of death openly and reported a 24% reduction in breast cancer mortality for Östergötland (Tabar, 2000), whereas a meta-analysis of the Swedish trials based on an official cause of death register reported only a 10% reduction for Östergötland (Nyström, 2002). The trial authors reported 10 fewer deaths from breast cancer in the study group despite slightly longer follow up, and 23 more deaths in the control group. They have not provided a plausible explanation of this large discrepancy (Duffy, 2002; Tabar, 2002). In, 2009, “a complete audit of breast cancer cases and deaths” in the Two-County trial was published, but it is not convincing (Holmberg, 2009). There was no blinding; it was not an independent audit; there was no attempt at producing a new data set based on the clinical records (which were only retrieved “where necessary”); and the Two-County trialists were directly involved with interpretations and resolving disagreements.

The bias seems to favour screening even when cause of death is determined blindly. In the New York trial, differential misclassification might be responsible for about half of the reported breast cancer mortality benefit. A similar number of dubious cases were selected for blinded review from each group, but a much smaller proportion of the screened group were finally classified as having died from breast cancer (Gøtzsche, 2004).

Furthermore, although the mammographic equipment was standard at the time, its performance was poor. Only 15% of 299 cancers in the study group were detected solely by mammography, and mammography did not identify a single case of minimal breast cancer (< 1 cm) (Thomas, 1977). The New York trial reported a 35% reduction in breast cancer mortality after seven years, but we consider it unlikely that it was a true effect.

In conjunction with the first meta-analysis of the Swedish trials, causes of death were reclassified blindly in some patients (Nyström, 1993). Breast cancer was considered the underlying cause of death in 419 of the screened group and 409 of the control group according to Statistics Sweden, and in 418 and 425 cases according to the committee (Nyström, 1993). The fact that all 17 reclassifications favoured the screened group suggests differential misclassification. This bias is difficult to avoid (Gøtzsche, 2001). Early cancers are treated by lumpectomy and radiotherapy, and radiotherapy reduces the rates of local recurrence by about two-thirds (EBCTCG, 2000). This might increase the likelihood that deaths among screen-detected breast cancer cases will be misclassified as deaths from other causes (EBCTCG, 1995) and that too many deaths in the control group will be misclassified as breast cancer deaths. In fact, for the Swedish trials it was stated that “most patients with locally advanced disease will die due to cancer” and that breast cancer as the underlying cause of death includes women with locally advanced breast cancer, whereas women who have been treated successfully should not be classified as having breast cancer deaths if another specified disease could be the cause of death (Nyström, 2000). The use of an official cause of death register as in more recent meta-analyses (Nyström, 2002) cannot solve these problems.

Postrandomisation exclusion of women who already had breast cancer at the time of entry to the trial is another possible source of bias. The exclusions were sometimes made many years after the trial started, or even after it had ended. In the Two-County trial, only women who were considered to have died from breast cancer were excluded (Nixon, 2000), a highly bias-prone process because those assessing cause of death were not blinded for screening status. Furthermore, the process seemed not to have been adequately monitored as it was not possible to identify prior breast cancers in Östergötland, by cluster (Nixon, 2000). It should therefore not be possible to do analyses that respect the clustering with those women excluded, although such analyses have been reported (Tabar, 1989; Tabar, 1990; Tabar, 1991; Tabar, 1995). A study that used the same registers as those used by the trialists found that a large number of breast cancer cases and deaths seemed to be missing in reports on the Two-County trial (Zahl, 2006). Another study found that the large reduction in breast cancer mortality agreed poorly with the cancer stages that were reported (Zahl, 2001).

The largest effects on breast cancer mortality were reported in trials that had long intervals between screenings (Two-County trial), invited a large fraction of the women to only two or three screenings (Two-County and Stockholm trials), started systematic screening of the control group after three to five years (Two-County, Göteborg and Stockholm trials), had only one-view mammography rather than two views (Two-County trial), and that had poor equipment for mammography (New York trial); and the cancers found with mammography were considerably smaller in the Canadian trial than in the Two-County trial (Narod, 1997). This suggests that differences in reported effects are related to the risk of bias in the trials rather than to the quality of the mammograms or the screening programmes. The sensitivity of mammographic readings in the trials that followed the New York trial has not consistently improved (Fletcher, 1993; IARC, 2002) and meta-analyses have failed to find an association between mammographic quality and breast cancer mortality (Glasziou, 1995; Kerlikowske, 1995). A meta-analysis found that the effect of screening was largest in those trials that found fewest node-positive cancers in the screened group relative to the control group (Gøtzsche, 2011). However, the regression line was in the wrong place. A screening effectiveness of zero (same proportion of node-positive cancers in the screened group as in the control group) predicted a 16% reduction in breast cancer mortality after 13 years (95% CI 9% to 23% reduction). This can only occur if there is bias, and there was bias for both variables, assessment of cause of death and of the number of node-positive cancers.

Several of the trials had clinical examination or regular self-examination of the breasts as part of their design (see ‘Description of studies’) but this is not likely to have had a major influence on the effect estimates. The effect of clinical examination is uncertain, and large randomised trials did not find an effect of self-examination (Kösters, 2003).

#### Cancer mortality

The major difficulty in assessing cause of death might have occurred when the patients were diagnosed with more than one malignant disease (Miller, 2001). The importance of autopsy is illustrated by the fact that 21% of the women with breast cancer who died in the Malmö trial had two or three types of different cancers (Andersson, 1988a; Janzon, 1991). Patients with cachexia and no signs of recurrence of breast cancer would likely be assigned to another type of cancer.

Since cancer mortality is likely to be less subject to bias than breast cancer mortality, we calculated what the expected cancer mortality (including breast cancer mortality) would be if the reported reduction in breast cancer mortality of 29% after seven years for the suboptimally randomised trials (Analysis 1.1) were true. Weighting the four trials that provided data on number of cancer deaths (Analysis 1.8), the expected risk ratio was 0.95.

However, all-cancer mortality in these trials was not reduced (RR 1.00, 95% CI 0.96 to 1.05), and this estimate was higher than what was expected (P = 0.02). This provides further evidence that assessment of cause of death was biased in favour of screening. Data from the Two-County trial (Tabar, 1988) illustrates the misclassification directly (Analysis 1.21)(Gøtzsche, 2004). Among women with a diagnosis of breast cancer, mortality for other cancers was higher in the screened group and mortality from all other causes also tended to be higher. The increase in mortality for causes other than breast cancer amounts to 38% of the reported decrease in breast cancer mortality in the Kopparberg part of the trial and 56% in the Östergötland part.

It has been shown that belief in the effectiveness of an intervention may influence the decision on which type of cancer caused the patient’s death (Newschaffer, 2000). Also, lethal complications of cancer treatments are often ascribed to other causes. The size of this misclassification is 37% for cancer generally and 9% for breast cancer (Brown, 1993).

For our current update, we could include cancer deaths also from the UK Age trial. Screening still had no effect on cancer mortality, RR 1.00 (95% CI 0.96 to 1.04).

#### All-cause mortality

The trials were not powered to detect an effect on all-cause mortality, but it is an important outcome since the findings related to breast cancer mortality seem to be biased. The fact that trials including 600,000 women did not have power to show an effect on all-cause mortality clearly indicates the comparatively small effect of the intervention, if any. The complex designs and insufficient reporting precluded us from providing reliable estimates for all-cause mortality in the trials with suboptimal randomisation. Furthermore, these trials had introduced early screening of the control group or had differentially excluded women after randomisation. Incidentally, however, all-cause mortality after 13 years was the same in adequately randomised trials and in suboptimally randomised trials (RR 0.99, 95% CI 0.95 to 1.03; and RR 0.99, 95% CI 0.97 to 1.01, respectively). There were many more deaths after 22 years, and RR was now 1.01 (95% CI 0.99 to 1.94) for the adequately randomised trials, which speaks against any mortality benefit of mammography screening.

While the Göteborg trial found a reduction in all-cause mortality (Bjurstam, 2016), being the only trial to do so, it was substantially underpowered to show this, even if its estimated reduction in cause-specific mortality was correct. The reduction in all-cause mortality therefore supports that the trial was biased due to suboptimal randomisation and that its estimated effect on breast cancer mortality is unreliable.

In, 2000, the estimate reported for the four Swedish trials was RR 1.00 (95% CI 0.98 to 1.02) after adjustment for imbalances in age (Nyström, 2000). In, 2002, the authors reported a 2% non-significant reduction in all-cause mortality (RR 0.98, 95% CI 0.96 to 1.00) and stated that they would have expected a 2.3% reduction (Nyström, 2002). However, the calculation was incorrect and the expected reduction, given their results, was only 0.9% (Gøtzsche, 2002a). The error has been acknowledged (Nyström, 2002a; The Lancet Erratum, 2002) but the published response to our criticism was also incorrect (Nyström, 2002b). The reported decrease of 2% in total mortality corresponds to a 10% decrease in all-cancer mortality, which is not plausible (see ‘Cancer mortality’ above).

The Östergötland part of the Two-County trial contributed about half of the deaths in the, 2002 report and had a risk ratio for all-cause mortality of 0.98 (Nyström, 2002). The women were randomised to only 24 clusters. In the Edinburgh trial there were 87 clusters, but double as many women in the invited group belonged to the highest socioeconomic level compared to the control group (Alexander, 1994). Socioeconomic factors are strong mortality predictors and could easily explain a 2% reduction in all-cause mortality, but such data remain unpublished and are also unavailable for the other Swedish trials. It has been reported that pretrial breast cancer incidence and breast cancer mortality were similar in the study group and in the control group in Östergötland (Nyström, 2002), but the power of the test was very low (Gøtzsche, 2002a). In contrast, another report found that breast cancer mortality was 15% lower in the invited groups in the Two-Country trial and that correction for this difference changed the estimate of the effect from a 31% reduction to a 27% reduction in breast cancer mortality (Duffy, 2003).

It is not clear why the unadjusted and age-adjusted estimates for all-cause mortality were the same with an RR of 0.98. The, 2002 Swedish meta-analysis comprised 43,343 deaths whereas in the, 2000 meta-analysis of 27,582 deaths the estimates were RR 1.06 (95% CI 1.04 to 1.08) (Gøtzsche, 2000) and RR 1.00 (95% CI 0.98 to 1.02) (Nyström, 2000), with non-overlapping confidence intervals. The Kopparberg part of the Two-County trial was not available for the, 2002 meta-analysis, but this should not have made any difference since the RR for Kopparberg was 1.00 (95% CI 0.96 to 1.04) (Nyström, 2000). The only other difference is that the extended data for the Malmö trial (MSST II) were included, but this trial contributed only 702 deaths (1.6%).

All-cause mortality has been reported to be lower in the Two-County trial when the analysis was confined to women with breast cancer (Tabar, 2002a). Such subgroup analyses are very unreliable, as are similar analyses in historically controlled studies (Tabar, 2001; Tabar, 2003a), since many breast cancer cases in the screened groups will have an excellent prognosis because of overdiagnosis and length bias (Berry, 2002).

#### Overdiagnosis and overtreatment

Overdiagnosis is an inevitable consequence of cancer screening and a critically important source of harm (IARC, 2002). Screening primarily identifies slow-growing cancers and cell changes that are biologically benign (Doll, 1981; Ernster, 1996; Fox, 1979). This is because slow-growing tumours have existed for longer than fast-growing tumours in the detectable range of tumour sizes and are therefore more likely to be detected at a screening session (length bias). Survival of women with screen-detected cancers is therefore very high, for example 97% in Malmö after 10 years (Janzon, 1991). Even within the same stage, it is higher than for cancers detected clinically (Moody-Ayers, 2000).

The level of overdiagnosis and overtreatment was about 25% in the trials that did not introduce early screening in the control group, and somewhat larger (33%) in the suboptimally randomised trials before the control group screen (Analysis 1.23). This is apart from the New York trial, which is unreliable since far more breast cancer cases were excluded from the screened group than from the control group (Shapiro, 1977; Shapiro, 1982; Shapiro, 1989). The true increase in surgery is considerably larger, however. As the excess surgery in the trials is similar to the increase in diagnoses, reoperations have likely not been included, although many women are operated upon more than once. In New South Wales, for example, one third of women with carcinoma in situ had either mastectomy alone (19%) or after breast conserving surgery (17%) (Kricker, 2000). The method of surgery has changed substantially since the trials were done and less invasive techniques are preferred today. The certainty of the estimates from the trials was downgraded for this reason (indirectness).

Large observational studies support that breast screening causes substantial overdiagnosis and overtreatment. Incidence increases of 40% to 60% since screening was implemented have been reported for Australia, Finland, Norway, Sweden, UK and USA (Barratt, 2005; Douek, 2003; Fletcher, 2003; Gøtzsche, 2004; IARC, 2002; Jonsson, 2005; Morrell, 2010; Ries, 2002; Zahl, 2004. In two additional studies, overdiagnosis was calculated as the percentage of all diagnoses, rather than the percentage of additional diagnoses; correcting for this gives an overdiagnosis of 45% in USA (Bleyer, 2012) and 18-33% in Norway (Kalager, 2012). The Norwegian estimate did not include carcinoma in situ and was also an underestimate for other reasons (Jørgensen, 2012). A small study from Copenhagen claimed that it is possible to screen without overdiagnosis, but it showed the expected prevalence peak, had very little power and provided no statistical analyses in support of the claim (Olsen, 2003). A study that included the whole of Denmark and also non-screened age groups found 33% overdiagnosis (Jørgensen, 2009a). A systematic review that adjusted for decreases in incidence, if any, in older age groups no longer screened, and also for the trend in background incidence, found an overdiagnosis of 35% for invasive cancer and 52% when carcinoma in situ was included, in countries with organised screening programmes (Jørgensen, 2009). Recently, based on long-term follow-up of the Canadian trials, overdiagnosis was estimated at 40% for the 40-49 years age group and 30% for the 50-59 year age group (Baines, 2016).

Data from the UK show that when screening was extended to the age group 65-70 years in, 2001, a sharp rise in invasive breast cancer incidence occurred in these women although they had been offered screening many times when they were younger and had already contributed to a massive increase in the incidence of DCIS and invasive cancers (Jørgensen, 2011). This is difficult to explain unless we assume that many screen-detected cancers would have regressed spontaneously if left alone, which is supported by a study from Norway with a strong design (Zahl, 2008), and by a similarly designed study from Sweden (Zahl, 2011). A US study also suggested that some breast cancers regress, since the incidence declined much too rapidly after the use of hormone replacement therapy stopped (Chlebowski, 2009). Another US study, of the breast cancer incidence and mortality rates during the period, 1975 to, 2000 when screening was introduced found that, in order to explain the observed trends, it was necessary to postulate that approximately 40% of the observed cancers had limited malignant potential and would have regressed if undetected (Fryback, 2006).

Screening increased the number of mastectomies by 20%. Since screening advances the time of diagnosis, a policy change towards more lumpectomies could have led to an overestimate. However, the policy change has occurred slowly (Nattinger, 2000) and even in the period, 1993 to, 1995, 52% of breast surgery in California was mastectomy (Malin, 2002). In Stockholm, the increase in mastectomies was larger after five years of screening (25%) than after the first round (16%), and when screening was introduced in Southeast Netherlands, the rate of breast-conserving surgery increased by 71% while the rate of mastectomy increased by 84% (Gøtzsche, 2002) despite the fact that this study did not include carcinoma in situ. The percentage of cases of carcinoma in situ treated by mastectomy declined from 71% in, 1983 to 40% in, 1993 in USA, but the estimated total numbers of mastectomies for this condition increased almost three-fold (Ernster, 1997). In the UK, mastectomies increased by 36% for invasive cancer and by 422% for carcinoma in situ from, 1990 to, 2001 (Douek, 2003). Carcinoma in situ is more often treated by mastectomy than invasive cancer (Patnick, 2012).

Conversely, use of mammography in the control group would lead to an underestimate of overdiagnosis. In the trials from Malmö and Canada, 24% (Andersson, 1988), 17% (Miller, 1992b) and 26% (Baines, 1994) of the women in the control group reported having received a mammogram during the trial; in the Two-County trial, it was 13% (Tabar, 1985); in the Göteborg trial, 18% of women in the control group received a mammogram in a two-year period during the trial (Bjurstam, 2003). In the Stockholm trial, 25% of those attending the first screening had had a mammogram in the two previous years (Frisell, 1989a), and in the Göteborg trial, as many as 51% of the women in the age group 39-49 had ever received a mammogram (Bjurstam, 1997). It is difficult to understand that this trial, with so much contamination reducing the observed benefit, reported a 45% reduction in breast cancer mortality.

The documented increase in mastectomies contrasts with assertions by trialists (Tabar, 1989), policy makers (Statusrapport, 1997; Swed Cancer Soc, 1996; Westerholm, 1988), websites supported by governmental institutions and advocacy groups (Jørgensen, 2004), and invitational letters sent to women invited to screening (Jørgensen, 2006; Gøtzsche, 2009) that early detection spares patients more aggressive treatments, in particular mastectomy. This is likely because the focus is on an individual woman who is diagnosed earlier and not on the effects of breast screening at the population level. Publications that base their claims on numbers that include the control group screen (Tabar, 2003) are also misleading, as are presentations of relative numbers rather than absolute numbers (Statusrapport, 1997). The proportion of breast preserving operations is said to be increasing, but the trend for the number of mastectomies is not revealed. A small study from Florence, without a control group (Paci, 2002), was also unreliable (Gøtzsche, 2002b). The authors asserted that if screening increased the number of mastectomies, populations in which screening has been introduced should see a subsequent increase.

Obviously, since the mastectomy rate has gone down steadily throughout many years, also in countries without screening, it is only to be expected that the authors found a decrease in the mastectomy rate when screening was introduced.

Denmark has a unique control group, as only 20% of the population was screened throughout 17 years. The large increase in mastectomies when screening was introduced has not been compensated later or by a corresponding decline in older age groups (Jørgensen, 2011). A study from Norway has confirmed this (Suhrke, 2011).

Quality assurance programmes could possibly reduce the surgical activity to some degree, but they could also increase it. In the UK, for example, the surgeons were blamed for not having treated even more women with carcinoma in situ by mastectomy (BASO audit, 2000), and the number of women treated by mastectomy almost doubled from, 1998 to, 2008 (Dixon, 2009).

Two to three years after breast cancer treatment, 47% of the women reported pain, usually several times a week (Gärtner, 2009). Only half of those with pain reported that it was mild (corresponding to 1-3 on a 10-point scale). The pain was equally common among those who had had breast-conserving surgery as among those with a mastectomy, and pain was more common when the women had had radiotherapy. Thus, half of all the overdiagnosed women will suffer from chronic pain, presumably for the rest of their lives.

#### False-positive diagnoses, psychological distress and painful mammograms

False-positive diagnoses can cause considerable and sustained psychological distress (Bülow, 2000; Salz, 2010), not only initially (Brodersen, 2006) but for years after the women are declared free from cancer (Brodersen, 2013). Many women experience anxiety, worry, despondency, sleeping problems, negative impact on sexuality and behaviour, and changes in their relationships with family, friends, and acquaintances as well as in existential values (Brodersen, 2006; Brodersen, 2007; Brodersen, 2013; Salz, 2010). In a large study, the severity of the psychological distress for women with false-positive findings was between that for healthy women and those with breast cancer even three years after they had been declared free from cancer (Brodersen, 2013). Some women will feel more vulnerable about disease and see a doctor more often (Barton, 2001).

In the Stockholm trial, one-third of women with false-positive findings were not declared cancer-free at six months (Lidbrink, 1996). In the UK, women who had been declared cancer-free after additional testing or biopsies were twice as likely to suffer psychological consequences three years later than women who received a clear result after their last mammogram (Brett, 2001). In the USA, three months after they had false-positive results, 47% of women who had highly suspicious readings reported that they had substantial anxiety related to the mammogram, 41% had worries about breast cancer, 26% reported that the worry affected their daily mood, and 17% that it affected their daily function (compared to 3% with a normal mammogram) (Lerman, 1991). In Norway, 18 months after screening mammography 29% of women with false-positive results and 13% of women with negative results reported anxiety about breast cancer (Gram, 1990).

The cumulative risk of a false-positive result after 10 mammograms ranges from about 20% to 60% (Barratt, 2005; Castells, 2006; Christiansen, 2000; Elmore, 1998; Hofvind, 2004; Hubbard, 2011; Johns, 2010; Njor, 2007). It is considerably higher in USA than elsewhere, e.g. the recall rate in women aged 50 to 54 years was 13% to 14% after the first mammogram, compared to 8% in the UK (Smith-Bindman, 2003). The reported percentages are often too low because recalls due to poor technical quality of the mammogram are not included (Hofvind, 2004; Johns, 2010; Njor, 2007), although these women may be just as affected by such recalls as by a real suspicion of cancer (Brodersen, 2006). In USA, 19% would have had a biopsy after 10 mammograms (Elmore, 1998).

Thus, it seems that screening inflicts important psychological distress on more than a quarter of the healthy population of women who attend a screening programme. The women are often not being informed about this risk (Gøtzsche, 2009; Jørgensen, 2004; Jørgensen, 2006; Slaytor, 1998; Werkö, 1995) or the risk of receiving a diagnosis of carcinoma in situ (Gøtzsche, 2009; Jørgensen, 2004; Thornton, 1997).

About half of the women report that it is painful to have a mammogram taken (Armstrong, 2007; Miller, 2002a; McNoe, 1996), and half of the women who decline an invitation to the second round of screening note that the major reason was that their first mammogram was painful (Elwood, 1998).

### Overall completeness and applicability of evidence

There are now so many data on the outcomes of breast cancer treatment and mammography screening that the results can be directly applied to policy making. There have been substantial advances in treatment since the trials were performed. Anti-hormones and polychemotherapy are effective also when the cancer has metastasized (EBCTCG, 2005), and the declines in breast cancer mortality we have seen in both screened and non-screened, otherwise comparable populations (Autier, 2010) have occurred rather uniformly across prognostic groups (Blamey, 2007). An updated meta-analysis of polychemotherapy showed that some regimens reduce breast cancer mortality by about one third, largely independently of tumour characteristics (EBCTCG, 2012). This means that the effect of screening must be smaller today than when the trials were conducted in terms of the number of women who may avoid dying of breast cancer.

In order to be effective, screening must lead to a reduction in the number of advanced cancers. In the USA, there has been a very small decrease in advanced cancers (Esserman, 2009; Jørgensen, 2011). A detailed analysis of a time period spanning 30 years showed that the incidence of early-stage breast cancer in USA went up from 112 to 234 cases per 100,000 women (a 109% increase) while the incidence of late-stage cancer decreased by 8%, from 102 to 94 cases per 100,000 women (Bleyer, 2012). Moreover, the small decline in advanced cancers was confined to regional disease involving the lymph nodes; there was no reduction in disease with distant metastases. A systematic review of several countries (Australia, Italy, Norway, Switzerland, the Netherlands, UK and the USA) found that, on average, the rate of cancers larger than 20 mm was not affected by screening (Autier, 2011). In Norway, screening did not decrease the incidence of cancers in stages III and IV, as the reductions were exactly the same in screened and non-screened areas (Kalager, 2012).

In contrast to screening, increased breast cancer awareness seems to have been important. In Denmark, the average tumour size at diagnosis was 33 mm in, 1978-79, but only 24 mm ten years later, in, 1988-89 (Rostgaard, 2010). This change occurred before screening started, and in contrast to screening, breast cancer awareness is unlikely to cause overdiagnosis. The difference of 9 mm is much greater than the average difference between the screened and the control groups in the trials, which was only 5 mm (Gøtzsche, 2012a), despite the fact that the small overdiagnosed tumours would tend to spuriously exaggerate the difference. In Canada, the size of clinically detected tumours decreased by 4 mm from, 1987 to, 1999 (Narod, 2011).

There are many poor observational studies claiming large effects of screening, but they often use statistical models with unsupported assumptions or misleading comparisons (Gøtzsche, 2010; Gøtzsche, 2012). The better studies rely on unmodified data. As noted above, Denmark has a unique control group, as only 20% of the population was screened throughout 17 years. The annual decline in breast cancer mortality in the relevant age group and time-period was 1% in the screened areas and 2% in the non-screened areas. In women who were too young to benefit from screening the declines were larger, 5% and 6%, respectively (Jørgensen, 2010). Also in the UK, Sweden and Norway, there was no visible effect of screening when relevant age groups were compared (Jørgensen, 2010; Kalager, 2010; Jørgensen, 2011). The Norwegian study (Kalager, 2010) was criticized because of short follow-up, but the follow-up was 6.6 years, which is when an effect was seen in the trials.

A study reported a 15% effect in the USA (Berry, 2005), but the authors noted that the decline in breast cancer mortality coincided not only with widespread propagation of screening but also with increasing use of adjuvant therapy. They also noted that slight variations in modelling assumptions could result in marked changes in estimated effects, and the statistical models adjusted for an increase in breast cancer incidence, which was inappropriate, as much of this increase was overdiagnosis. Unlike the USA, women below age 50 years are rarely offered screening in Europe. The mean decline in breast cancer mortality between, 1989 and, 2005 in these women was 37%, whereas it was 21% in women aged 50-69 years (Autier, 2010). The declines began before organised screening in many countries and fitted better with the introduction of tamoxifen, which explains the larger decline in women too young to have been offered screening who often have oestrogen-sensitive tumours (Jørgensen, 2011). A comparison of three pairs of neighbouring European countries that had introduced screening 10-15 years apart showed no relation between screening start and the reductions in breast cancer mortality (Autier, 2011a); in fact, the reduction in breast cancer mortality was about the same in the six European countries as in USA (Bleyer, 2011). An Australian study found that most, if not all, of the reduction in breast cancer mortality could be attributed to adjuvant hormonal and chemotherapy (Burton, 2011).

Screening advocates have claimed that screening explains why breast cancer mortality rates are lower in Sweden than in Denmark (Dean, 2010), but this difference existed decades before screening. Further, the reductions in breast cancer mortality in the screening period were largest in Denmark, 49% versus 36% in Sweden in women under 50, although half of these women were invited in Sweden versus none in Denmark (Autier, 2010). In those aged 50-69 years, the reduction was 26% in Denmark versus 16% in Sweden, although only 20% of Danish women were invited, versus all in Sweden where more than 80% participated (Autier, 2010; IARC, 2002). Despite having the longest running programme, the widest invited age range, and the shortest screening interval in Europe (IARC, 2002), Sweden has experienced lower reductions in breast cancer mortality than the European median (Autier, 2010).

These studies taken in combination cast doubt as to the effectiveness of screening today. And even if screening reduces breast cancer mortality, the evidence does not support an effect on all-cause mortality or on total cancer mortality. However, both the randomised and non-randomised studies provide evidence that screening causes substantial overdiagnosis and false-positive results are very common.

### Quality of the evidence

For the adequately randomised trials, the certainty of the effect on breast cancer mortality was assessed as low. The certainty was downgraded 1 level due to the age of the trials and substantial changes in screening technology and improvements in treatment. The certainty was also downgraded due to imprecision, as the confidence interval included clinically relevant effects. The certainty of the effect estimates for most of the remaining outcomes was assessed as moderate, downgraded 1 level due to the age of the trials. As our conclusions are based on the subgroup of adequately randomied trials, we did not have to downgrade for risk of bias. Results of the suboptimally randomised trials were downgraded 1 level due to risk of bias and the certainty of the evidence for this subgroup is thus ‘very low’. As heterogeneity was explained by dividing trials into subgroups according to adequate versus suboptimal randomisation, and as there was little or no heterogeneity within each subgroup, we did not downgrade the certainty of evidence for each subgroup due to inconsistency. We did not detect or have reason to suspect publicatioin bias and thus did not downgrade for this domain. As effects were either not documented or comparatively small, we did not upgrade the certainty of the evidence due to large effects. As we included only randomised trials, counfounding effects did not impact our assessment of the certainty of the evidence. We did not detect any dose-effect relationship and thus did not upgrade based on this domain.

### Potential biases in the review process

We took great care to avoid introducing bias in the review process and to take account of the apparent biases in the randomised trials. Our most important judgement was that we found it necessary to divide trials into subgroups according to method of randomisation. Cluster-randomised trials and trials with unclear randomisation methods often had other potentially important risks of bias as well, compared to individually randomised trials with adequate randomisation methods. We found indications that suboptimal randomisation methods were associated with important baseline differences, such as fewer breast cancers being detected in the screening arm (Bjurstam, 2016), which is contrary to expectations. The confidence interval between the most and the least optimistic trial result for breast cancer mortality did not overlap and the estimated effect differed more than what would be expected without systematic differences between the trials (a 42% reduction versus no effect). Separating trials acccording to randomisation method explained the heterogeneity.

### Agreements and disagreements with other studies or reviews

Previous reviews have generally not heeded the methodological quality of the trials, but when the methods were assessed blindly, the researchers judged the Canadian trial to be of high quality and the Two-County trial to be of poor quality (Glasziou, 1995).

Prompted by our first Cochrane review in, 2001, the US Preventive Services Task Force performed an updated systematic review (Humphrey, 2002). It excluded the Edinburgh trial and reported a 16% reduction in breast cancer mortality for all ages. The authors noted that, “the mortality benefit of mammography screening is small enough that biases in the trials could erase or create it” and were concerned whether, across all age groups, the magnitude of benefit is sufficient to outweigh the harms. The Task Force gave mammography screening a grade B recommendation (US Task Force, 2002). The Task Force reported a 15% reduction in breast cancer mortality for those aged 39 to 49 years in, 2009 and larger effects in older age groups (Nelson, 2009). A comprehensive IARC report (IARC, 2002) was not a systematic review and paid little attention to the varying quality of the trials; it even included a non-randomised study in its meta-analysis. A, 2012 UK review was not a systematic review either (UK review, 2012). It used data from the Cochrane review for the benefit, but lumped the adequately randomised trials with the suboptimally randomised trials and did not take account of the improvements in treatment and breast cancer awareness. The report focussed on breast cancer mortality, and ignored all cause mortality, which biased its findings in favour of breast screening. It acknowledged that previous estimations of the benefits and harms of mammography screening had been overoptimistic and acknowledged uncertainties around estimations of the magnitude of benefit. It also acknowledged and estimated overdiagnosis as a major harm of breast screening, but did not use the Cochrane review estimate but a smaller one that was diluted because of screening in the control group (Welch, 2006).

The meta-analyses of the Swedish trials are not systematic reviews as they do not include all relevant trials. There is a high risk of bias in cluster randomised trials with few clusters (Puffer, 2003) and numbers of randomised women were inconsistently reported (Table 1). In Stockholm, for example, the number of randomised women decreased by 4.5% in the screening group but increased by 3.6% in the control group (Gøtzsche, 2000) in the Swedish, 1993 review (Nyström, 1993) compared to the trial report (Frisell, 1997). In the, 2000 and, 2002 reviews (Nyström, 2000; Nyström, 2002), numbers have increased by 1.6% in both groups but should have been the same as in the, 1993 report since all women were identified through their unique identification number (Nyström, 2002), which has been used in Sweden for several decades; exclusions of women with previous breast cancer was completed with the, 1993 review; and all three reviews were based on the exact age at randomisation, and the age range was the same. The varying numbers therefore indicate that the randomisation was not respected. The estimates in the Swedish reviews were adjusted for differences in age, but since the distribution of age would be expected to differ over socioeconomic strata, such adjustment would be expected to lead to other imbalances (Gøtzsche, 2000). Furthermore, simulation studies have shown that adjustments quite often increase bias rather than reduce it (Deeks, 2003). The most recent review of the Swedish trials reported a 15% reduction in breast cancer mortality with the follow-up model (Nyström, 2002); another estimate of 21% was based on an ‘evaluation model’, which is flawed, as it ignores breast cancer deaths among women in the control group whose breast cancer diagnosis was made after the first screening round of the control group (Berry, 1998).

## Data Availability

Data are publicly available

## Authors’ conclusions

### Implications for practice

The most reliable trials did not support that breast screening reduces breast cancer mortality for the included age groups while trials that provided very low certainty evidence indicated a benefit. Breast cancer mortality has declined over the past decades, with the greatest reductions in women below the age group commonly invited to screening (Autier, 2010), likely due to improved treatments and increased breast cancer awareness. As breast cancer mortality is an unreliable outcome that is biased in favour of screening, it is noteworthy that screening did not reduce total cancer mortality or total mortality, neither in the adequtely randomised trials, nor in the suboptimally randomised trials. Overdiagnosis has human costs; increases the use of mastectomies; and increases mortality. Women, clinicians and policy makers should consider the trade-offs and the uncertainties of these data carefully when they decide whether or not to attend or to offer breast screening programmes.

### Implications for research

We do not see any need for more mammography screening trials of the type we have reviewed. Research is needed to identify means of separating screen-detected cancers likely to result in death from cancers and cell changes identified by screening that do not need treatment. Several such trials are currently ongoing.

## Acknowledgements

For this, 2023 update, we would like to acknowledge and thank the following people for their help in assessing the search results for this review via Cochrane’s Screen4Me workflow:

Susanna Wisniewski, Mary MacCara, Igor Svintsitskyi, Nicole Askin, Therese Dalsbø, Ante Topić, Nikolaos Sideris, Anna Noel-Storr, Lai Ogunsola, Shammas Mohammed, Adrià Bermudo-Gallaguet, William Garrett, Ivan Perez-Neri, Simon-Peter Gom, Amin Sharifan, Hadi Keshavarz, Vibor Milunović, Emerald Sy, Cheryl Sumner, Victor Ghosh, Anna Brocke, Amina Berour, Dr Manmohan Singh Shergillz, Yineli Villalobos Parra, Issa Hanna.

The following people conducted the editorial process for this article:

- Sign-off Editors (final editorial decision): Annabel Goodwin and Nicholas Wilcken, Co-ordinating Editors, Cochrane Breast Cancer; Peter Tugwell, Cochrane Editorial Board
- Managing Editor (selected peer reviewers, provided editorial guidance to authors, edited the article): Liz Bickerdike, Cochrane Central Editorial Service
- Editorial Assistant (conducted editorial policy checks, collated peer-reviewer comments and supported editorial team): Leticia Rodrigues, Cochrane Central Editorial Service
- Copy Editor (copy editing and production): [NAME, AFFILIATION];
- Peer-reviewers (provided comments and recommended an editorial decision): Nuala Livingstone and Rachel Richardson, Cochrane Evidence Production and Methods Directorate (methods), Afroditi Kanellopoulou, Cochrane Evidence Production and Methods Directorate (statistics), Jo Platt, Central Editorial Information Specialist (search), and Cecilia Fabrizio, DrPH (consumer). Two additional peer reviewers provided clinical peer review but chose not to be publicly acknowledged.

We thank Freda Alexander, Ingvar Andersson, Cornelia Baines, Niels Bjurstam, Gunnar Fagerberg, Jan Frisell, Anthony B Miller and Sam Shapiro for comments on their trials, Friederike M Perl for pointing out an inconsistency in one of the trials, Mike Clarke for advice, Ole Olsen who was an author on the, 2001 version of this review and wrote the draft section on methodological quality of the trials for that version, Kay Dickersin for comments on the, 2006 update of the review, and Margrethe Nielsen who was an author on the, 2006 and, 2009 updates.

## Data and analyses

### Comparison 1 Screening with mammography versus no screening

Data and analyses
| Comparison 1 |  |  |  |  |
| --- | --- | --- | --- | --- |
| Screening with mammography versus no screening |  |  |  |  |
| Outcome or subgroup title | No. of studies | No. of participants | Statistical method | Effect size |
| 1.1 Deaths ascribed to breast cancer, 7 years follow up | 11 |  | Risk Ratio (M-H, Fixed, 95% CI) | Subtotals only |
| 1.1.1 Adequately randomised trials | 4 | 292958 | Risk Ratio (M-H, Fixed, 95% CI) | 0.93 [0.79, 1.09] |
| 1.1.2 Suboptimally randomised trials | 7 | 323369 | Risk Ratio (M-H, Fixed, 95% CI) | 0.71 [0.61, 0.83] |
| 1.2 Deaths ascribed to breast cancer, 13 years follow up | 9 |  | Odds Ratio (M-H, Fixed, 95% CI) | Subtotals only |
| 1.2.1 Adequately randomised trials | 4 | 292153 | Odds Ratio (M-H, Fixed, 95% CI) | 0.90 [0.79, 1.02] |
| 1.2.2 Suboptimally randomised trials | 5 | 306937 | Odds Ratio (M-H, Fixed, 95% CI) | 0.75 [0.67, 0.83] |
| 1.3 Deaths ascribed to breast cancer, 7 years follow up, women below 50 years of age (Malmö 55) | 9 |  | Risk Ratio (M-H, Fixed, 95% CI) | Subtotals only |
| 1.3.1 Adequately | 3 | 227333 | Risk Ratio (M-H, Fixed, | 0.94 [0.78, 1.14] |

| Outcome or subgroup title | No. of studies | No. of participants | Statistical method | Effect size |
| --- | --- | --- | --- | --- |
| randomised trials |  |  | 95% CI) |  |
| 1.3.2 Suboptimally randomised trials | 6 | 129035 | Risk Ratio (M-H, Fixed, 95% CI) | 0.81 [0.63, 1.05] |
| 1.4 Deaths ascribed to breast cancer, 7 years follow up, women at least 50 years of age (Malmö 55) | 7 |  | Risk Ratio (M-H, Fixed, 95% CI) | Subtotals only |
| 1.4.1 Adequately randomised trials | 2 | 65625 | Risk Ratio (M-H, Fixed, 95% CI) | 0.88 [0.64, 1.20] |
| 1.4.2 Suboptimally randomised trials | 5 | 195419 | Risk Ratio (M-H, Fixed, 95% CI) | 0.67 [0.56, 0.81] |
| 1.5 Deaths ascribed to breast cancer, 13 years follow up, women below 50 years of age | 8 |  | Risk Ratio (M-H, Fixed, 95% CI) | Subtotals only |
| 1.5.1 Adequately randomised trials | 3 | 218697 | Risk Ratio (M-H, Fixed, 95% CI) | 0.87 [0.73, 1.03] |
| 1.5.2 New Subgroup | 5 | 110814 | Risk Ratio (M-H, Fixed, 95% CI) | 0.80 [0.64, 0.98] |
| 1.6 Deaths ascribed to breast cancer, 13 years follow up, women at least 50 years of age | 7 |  | Risk Ratio (M-H, Fixed, 95% CI) | Subtotals only |
| 1.6.1 Adequately randomised trials | 2 | 74261 | Risk Ratio (M-H, Fixed, 95% CI) | 0.94 [0.77, 1.15] |
| 1.6.2 Suboptimally randomised trials | 5 | 194613 | Risk Ratio (M-H, Fixed, 95% CI) | 0.70 [0.62, 0.80] |
| 1.7 Deaths ascribed to breast cancer, 24 years of follow-up | 2 | 250671 | Risk Ratio (M-H, Fixed, 95% CI) | 0.95 [0.86, 1.04] |
| 1.8 Deaths ascribed to any cancer, all women | 7 |  | Risk Ratio (M-H, Fixed, 95% CI) | Subtotals only |
| 1.8.1 Adequately randomised trials | 4 | 292954 | Risk Ratio (M-H, Fixed, 95% CI) | 1.00 [0.96, 1.04] |
| 1.8.2 Suboptimally randomised trials (unreliable estimates) | 3 | 195871 | Risk Ratio (M-H, Fixed, 95% CI) | 0.99 [0.93, 1.06] |
| 1.9 Overall mortality, 7 years follow up | 11 |  | Risk Ratio (M-H, Fixed, 95% CI) | Subtotals only |
| 1.9.1 Adequately randomised trials | 4 | 292958 | Risk Ratio (M-H, Fixed, 95% CI) | 0.98 [0.94, 1.03] |
| 1.9.2 Suboptimally randomised trials (unreliable estimates) | 7 | 324977 | Risk Ratio (M-H, Fixed, 95% CI) | 0.99 [0.96, 1.02] |
| 1.10 Overall mortality, 13 years follow up | 8 |  | Risk Ratio (M-H, Fixed, 95% CI) | Subtotals only |
| 1.10.1 Adequately randomised trials | 4 | 292958 | Risk Ratio (M-H, Fixed, 95% CI) | 0.99 [0.95, 1.03] |
| 1.10.2 Suboptimally randomised trials (unreliable estimates) | 4 | 244868 | Risk Ratio (M-H, Fixed, 95% CI) | 0.99 [0.97, 1.01] |
| 1.11 Overall mortality, 7 years follow up, women below 50 years of age | 7 |  | Risk Ratio (M-H, Fixed, 95% CI) | Subtotals only |
| 1.11.1 Adequately randomised trials | 2 | 211270 | Risk Ratio (M-H, Fixed, 95% CI) | 0.97 [0.90, 1.04] |
| 1.11.2 Suboptimally randomised trials (unreliable estimates) | 5 | 99656 | Risk Ratio (M-H, Fixed, 95% CI) | 1.07 [0.98, 1.16] |
| 1.12 Overall mortality, 7 years follow up, women at least 50 years of age | 5 |  | Risk Ratio (M-H, Fixed, 95% CI) | Subtotals only |
| 1.12.1 Adequately randomised trials | 1 | 39405 | Risk Ratio (M-H, Fixed, 95% CI) | 1.01 [0.85, 1.20] |
| 1.12.2 Suboptimally randomised trials (unreliable estimates) | 4 | 161519 | Risk Ratio (M-H, Fixed, 95% CI) | 0.97 [0.94, 1.00] |
| 1.13 Overall mortality, 13 years follow up, women below 50 years of age | 6 |  | Risk Ratio (M-H, Fixed, 95% CI) | Subtotals only |
| 1.13.1 Adequately randomised trials | 3 | 219324 | Risk Ratio (M-H, Fixed, 95% CI) | 0.98 [0.92, 1.04] |
| 1.13.2 Suboptimally randomised trials (unreliable estimates) | 3 | 61344 | Risk Ratio (M-H, Fixed, 95% CI) | 1.00 [0.92, 1.10] |
| 1.14 Overall mortality, 13 years follow up, women at | 4 |  | Risk Ratio (M-H, Fixed, 95% CI) | Subtotals only |

| Outcome or subgroup title | No. of studies | No. of participants | Statistical method | Effect size |
| --- | --- | --- | --- | --- |
| least 50 years of age |  |  |  |  |
| 1.14.1 Adequately randomised trials | 2 | 73634 | Risk Ratio (M-H, Fixed, 95% CI) | 1.00 [0.95, 1.04] |
| 1.14.2 Suboptimally randomised trials (unreliable estimates) | 2 | 98261 | Risk Ratio (M-H, Fixed, 95% CI) | 0.99 [0.97, 1.02] |
| 1.15 Overall mortality, 24 years follow-up | 2 |  | Risk Ratio (M-H, Fixed, 95% CI) | Subtotals only |
| 1.15.1 Adequately randomised trials | 2 | 250671 | Risk Ratio (M-H, Fixed, 95% CI) | 1.01 [0.99, 1.04] |
| 1.16 Number of mastectomies and lumpectomies | 5 | 250479 | Risk Ratio (M-H, Fixed, 95% CI) | 1.35 [1.26, 1.44] |
| 1.16.1 Adequately randomised trials | 3 | 132321 | Risk Ratio (M-H, Fixed, 95% CI) | 1.31 [1.22, 1.42] |
| 1.16.2 Suboptimally randomised trials | 2 | 118158 | Risk Ratio (M-H, Fixed, 95% CI) | 1.42 [1.26, 1.61] |
| 1.17 Number of mastectomies | 5 | 250479 | Risk Ratio (M-H, Fixed, 95% CI) | 1.20 [1.11, 1.30] |
| 1.17.1 Adequately randomised trials | 3 | 132321 | Risk Ratio (M-H, Fixed, 95% CI) | 1.20 [1.08, 1.32] |
| 1.17.2 Suboptimally randomised trials | 2 | 118158 | Risk Ratio (M-H, Fixed, 95% CI) | 1.21 [1.06, 1.38] |
| 1.18 Number treated with radiotherapy | 2 | 100383 | Risk Ratio (M-H, Fixed, 95% CI) | 1.32 [1.16, 1.50] |
| 1.18.1 Adequately randomised trials | 1 | 42486 | Risk Ratio (M-H, Fixed, 95% CI) | 1.24 [1.04, 1.49] |
| 1.18.2 Suboptimally randomised trials | 1 | 57897 | Risk Ratio (M-H, Fixed, 95% CI) | 1.40 [1.17, 1.69] |
| 1.19 Number treated with chemotherapy | 2 | 100383 | Risk Ratio (M-H, Fixed, 95% CI) | 0.96 [0.78, 1.19] |
| 1.19.1 Adequately randomised trials | 1 | 42486 | Risk Ratio (M-H, Fixed, 95% CI) | 0.63 [0.39, 1.04] |
| 1.19.2 Suboptimally randomised trials | 1 | 57897 | Risk Ratio (M-H, Fixed, 95% CI) | 1.06 [0.84, 1.34] |
| 1.20 Number treated with hormone therapy | 2 | 100383 | Risk Ratio (M-H, Fixed, 95% CI) | 0.73 [0.55, 0.96] |
| 1.20.1 Adequately randomised trials | 1 | 42486 | Risk Ratio (M-H, Fixed, 95% CI) | 0.81 [0.60, 1.08] |
| 1.20.2 Suboptimally | 1 | 57897 | Risk Ratio (M-H, Fixed, | 0.30 [0.12, 0.72] |
| randomised trials |  |  | 95% CI) |  |
| 1.21 Mortality among breast cancer patients in the Two-County study, 7 years follow up | 2 |  | Risk Ratio (M-H, Fixed, 95% CI) | Subtotals only |
| 1.21.1 Mortality from cancers other than breast cancer | 2 | 2063 | Risk Ratio (M-H, Fixed, 95% CI) | 2.42 [1.00, 5.85] |
| 1.21.2 Mortality from causes other than breast cancer | 2 | 2063 | Risk Ratio (M-H, Fixed, 95% CI) | 1.37 [0.93, 2.04] |
| 1.22 Results for biased trial | 1 |  | Risk Ratio (M-H, Fixed, 95% CI) | Totals not selected |
| 1.22.1 Deaths ascribed to breast cancer, 7 years follow up | 1 |  | Risk Ratio (M-H, Fixed, 95% CI) | Totals not selected |
| 1.22.2 Deaths ascribed to breast cancer, 13 years follow up | 1 |  | Risk Ratio (M-H, Fixed, 95% CI) | Totals not selected |
| 1.22.3 Deaths ascribed to breast cancer, 7 years follow up, younger women (below 50 years of age) | 1 |  | Risk Ratio (M-H, Fixed, 95% CI) | Totals not selected |
| 1.22.4 Deaths ascribed to breast cancer, 7 years follow up, elderly women (at least 50 years of age) | 1 |  | Risk Ratio (M-H, Fixed, 95% CI) | Totals not selected |
| 1.22.5 Deaths ascribed to breast cancer, 13 years follow up, younger women (below 50 years of age) | 1 |  | Risk Ratio (M-H, Fixed, 95% CI) | Totals not selected |
| 1.22.6 Deaths ascribed to breast cancer, 13 years follow up, elderly women (at least 50 years of age) | 1 |  | Risk Ratio (M-H, Fixed, 95% CI) | Totals not selected |
| 1.22.7 Overall mortality, 7 years follow up | 1 |  | Risk Ratio (M-H, Fixed, 95% CI) | Totals not selected |
| 1.22.8 Number treated with radiotherapy | 1 |  | Risk Ratio (M-H, Fixed, 95% CI) | Totals not selected |
| 1.23 Number of cancers | 7 | 512246 | Risk Ratio (M-H, Fixed, 95% CI) | 1.29 [1.23, 1.35] |

| Outcome or subgroup title | No. of studies | No. of participants | Statistical method | Effect size |
| --- | --- | --- | --- | --- |
| 1.23.1<br>Adequately randomised trials (after 7-9 years) | 4 | 292979 | Risk Ratio (M-H, Fixed, 95% CI) | 1.25 [1.18, 1.34] |
| 1.23.2<br>Suboptimally randomised trials (before control group screen) | 3 | 219267 | Risk Ratio (M-H, Fixed, 95% CI) | 1.33 [1.24, 1.44] |

## What’s new

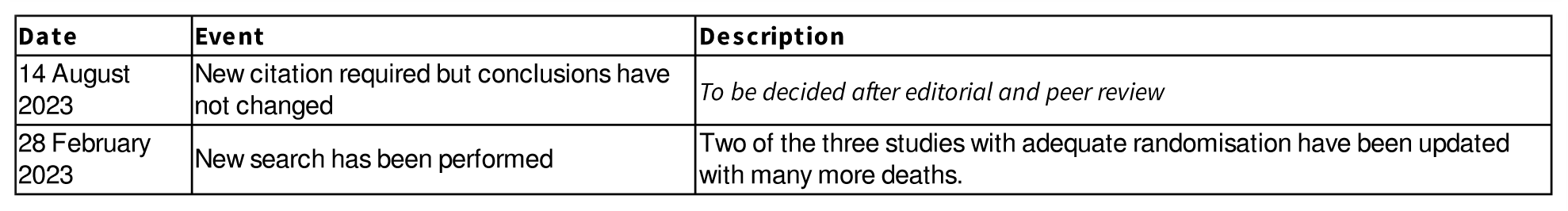

## History

Protocol first published: Issue 1,, 2000

Review first published: Issue 4,, 2001

| Date | Event | Description |
| --- | --- | --- |
| 22 November 2012 | New search has been performed | Performed search for new studies on 22 November 2012. No new studies included |
| 22 November 2012 | New citation required but conclusions have not changed | This review update includes an accumulation of changes in the discussion section |
| 17 November 2010 | Amended | Corrected labels for Figure 1.21. |
| 5 August 2009 | New citation required but conclusions have not changed | new citation = no change to conclusions |
| 3 March 2009 | New search has been performed | Data from a new trial, UK age trial, added. |
| 12 July 2006 | New citation required and conclusions have changed | Substantive amendment |

## Contributions of authors

PCG wrote the draft protocol. Two authors extracted the main data independently and contributed to the review. PCG is guarantor.

## Sources of support

### Internal sources

- Cochrane Denmark, Denmark Facilites for, 2023 update

### External sources

- Danish Institute for Health Technology Assessment, Denmark Financial support for the first version of this review

## Differences between protocol and review

A new outcome was added when we discovered that breast cancer mortality is an unreliable outcome. We have clarified that our outcome ‘number of cancers’ is an expression of the risk of overdiagnosis.

## Characteristics of studies

### Characteristics of included studies [ordered by study ID]

#### Canada, 1980

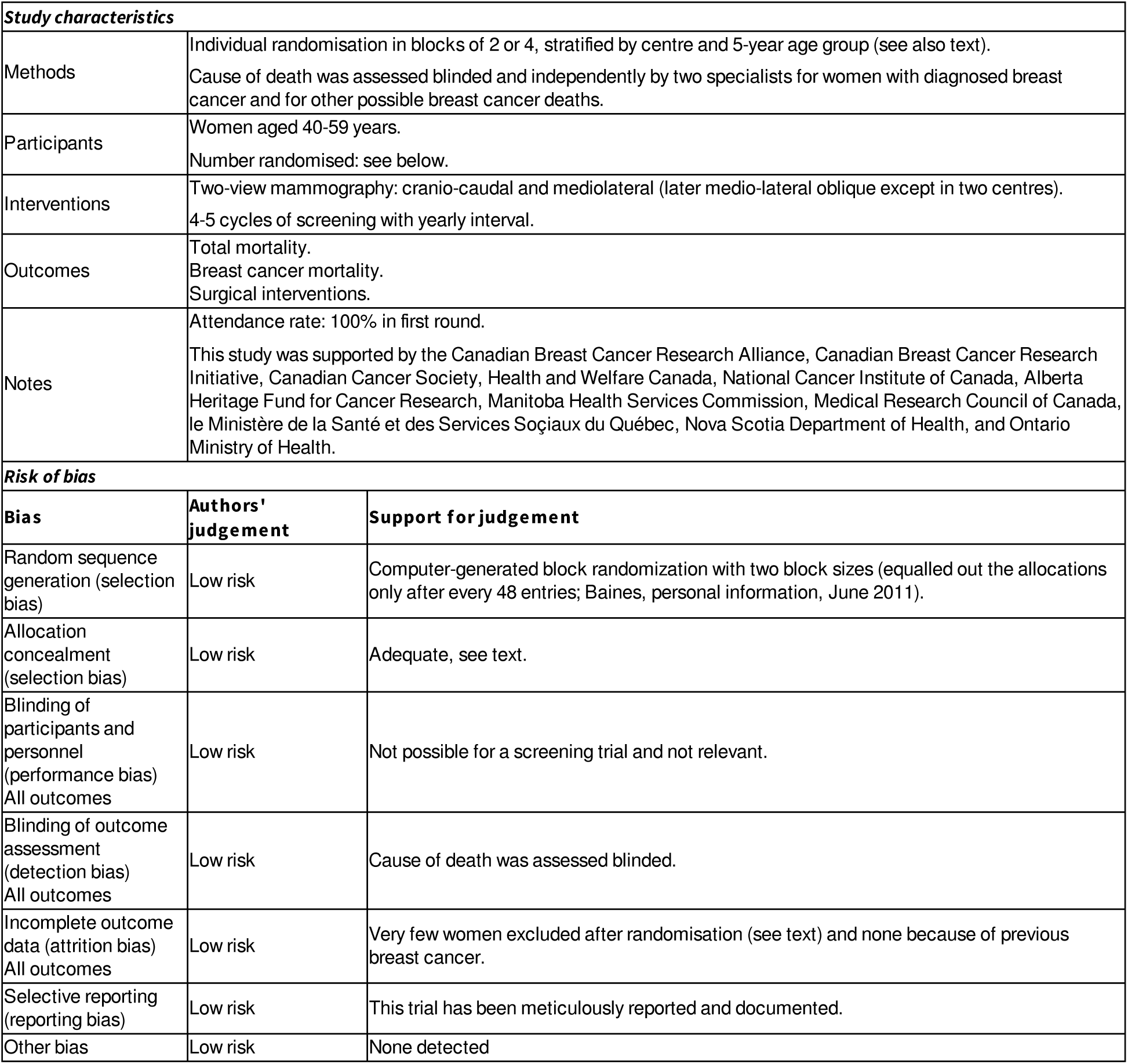

#### Canada, 1980a

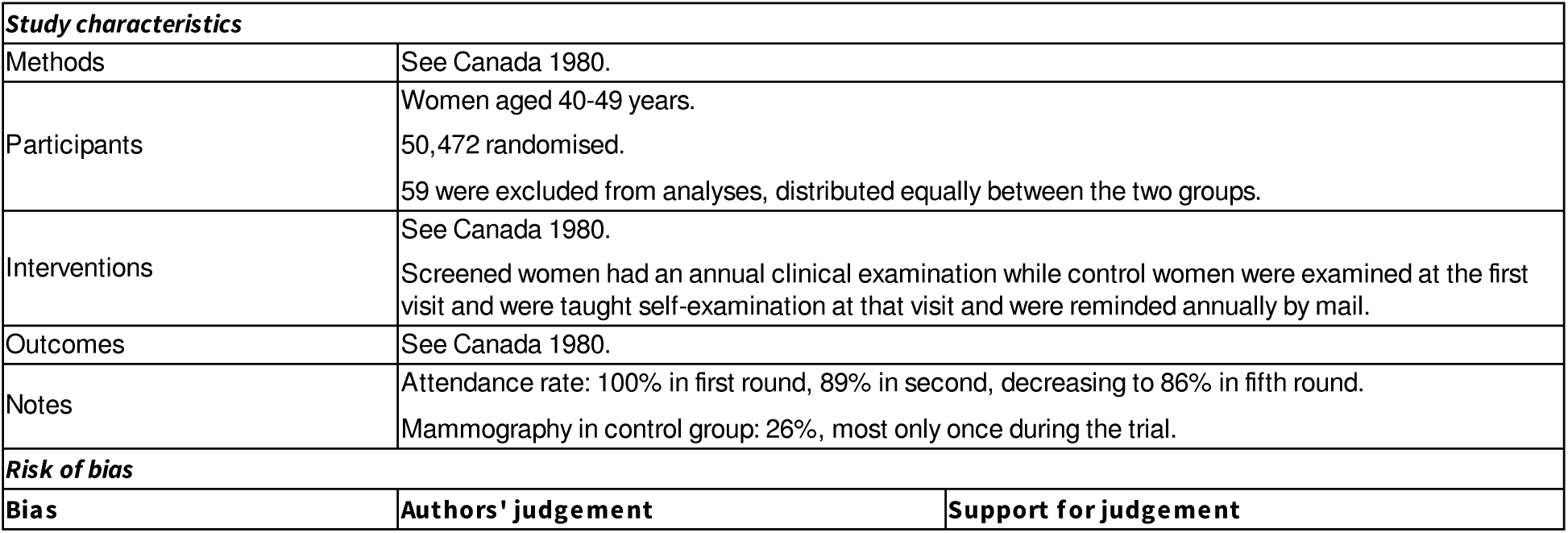
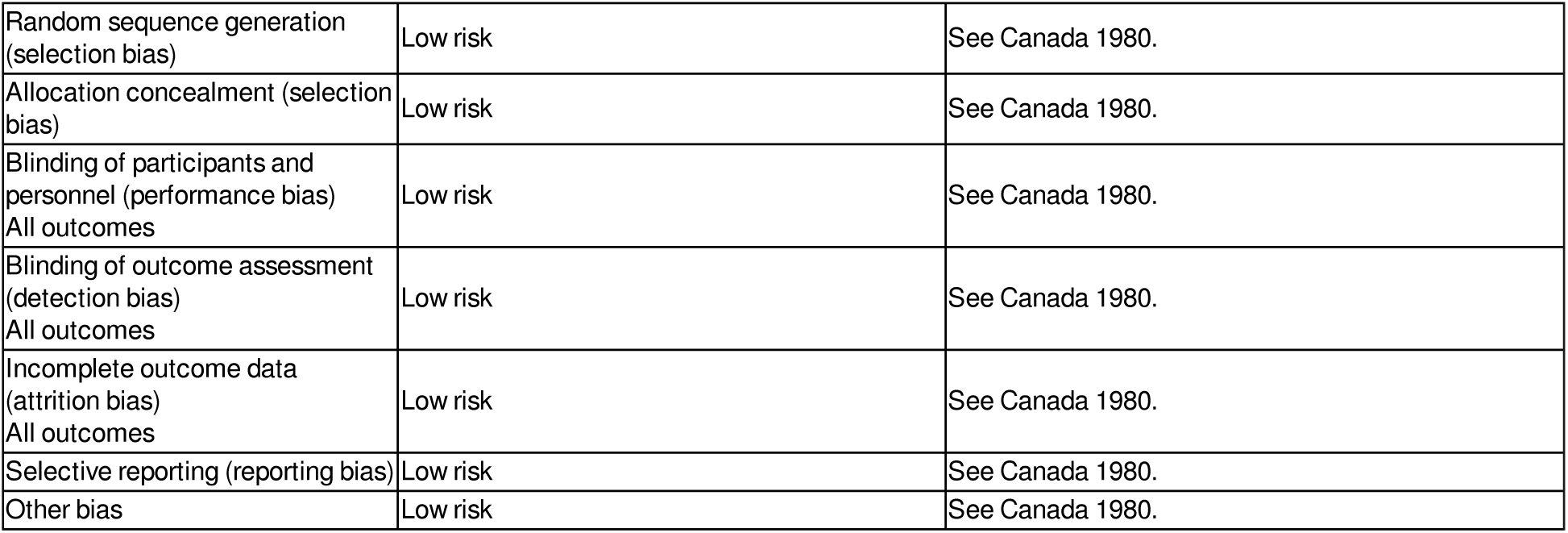

#### Canada, 1980b

Canada 1980b
| Study characteristics |  |  |
| --- | --- | --- |
| Methods | See Canada 1980. |  |
| Participants | Women aged 50-59 years.<br>39,459 randomised.<br>54 were excluded from analyses, distributed equally between the two groups. |  |
| Interventions | See Canada 1980.<br>All women had their breasts examined annually. |  |
| Outcomes | See Canada 1980. |  |
| Notes | Attendance rate: 100% in first round, 90% in second, decreasing to 87% in fifth round.<br>Mammography in control group: 17%. |  |
| Risk of bias |  |  |
| Bias | Authors' judgement | Support for judgement |
| Random sequence generation (selection bias) | Low risk | See Canada 1980. |
| Allocation concealment (selection bias) | Low risk | See Canada 1980. |
| Blinding of participants and personnel (performance bias)<br>All outcomes | Low risk | See Canada 1980. |
| Blinding of outcome assessment (detection bias)<br>All outcomes | Low risk | See Canada 1980. |
| Incomplete outcome data (attrition bias)<br>All outcomes | Low risk | See Canada 1980. |
| Selective reporting (reporting bias) | Low risk | See Canada 1980. |
| Other bias | Low risk | See Canada 1980. |

#### Edinburgh, 1978

Edinburgh 1978
| Study characteristics |  |  |
| --- | --- | --- |
| Methods | Stratified cluster randomisation; general practices were clusters; stratification was by size of practice.<br>About 87 clusters (numbers vary in different reports, see text).<br>Blinding of outcome assessment not stated. |  |
| Participants | Women aged 45-64 years.<br>Number of women and practices randomised inconsistently reported (see text).<br>Very biased exclusions occurred: exclusion procedures different in study and control group, 177 previous breast cancer cases excluded from control group and 338 from study group. |  |
| Interventions | Two-view mammography at first screen: cranio-caudal and oblique (except in one practice); only oblique in later rounds.<br>Screened group: mammography and physical examination year 1, 3, 5 and 7; physical examination year 2, 4 and 6.<br>Control group: usual care. |  |
| Outcomes | Total mortality.<br>Breast cancer mortality.<br>Radiotherapy. |  |
| Notes | Attendance rate: Circa 60% in first round; 44% in seventh round.<br>Mammography in control group: unknown.<br>This study was undertaken by the Cancer Screening Evaluation Unit, which receives support from the UK Department of Health. |  |
| Risk of bias |  |  |
| Bias | Authors' judgement | Support for judgement |
| Random sequence generation (selection bias) | High risk | No information, but some clusters later changed allocation status. |
| Allocation concealment (selection bias) | High risk | The randomisation failed to an important degree to create comparable groups. |
| Blinding of participants and personnel (performance bias)<br>All outcomes | Low risk | Not possible for a screening trial and not relevant. |
| Blinding of outcome assessment (detection bias)<br>All outcomes | High risk | Not stated. |
| Incomplete outcome data (attrition bias)<br>All outcomes | High risk | Not relevant, as randomisation failed to create comparable groups. |
| Selective reporting (reporting bias) | Unclear risk | Not relevant, as randomisation failed to create comparable groups. |
| Other bias | High risk | Not relevant, as randomisation failed to create comparable groups. |

#### Göteborg, 1982

Study characteristics
|  |  |
| --- | --- |
| Methods | See Göteborg 1982a and 1982b. |
| Participants | Women aged 39-59 years.<br>Number of women randomised: 21,904 to screening, 30,318 to control (see also text).<br>254 women (1.2%) excluded from the screening group and 357 (1.2%) from the control group due to a history of breast carcinoma prior to randomisation. |
| Interventions | See Göteborg 1982a and 1982b. |
| Outcomes | Total mortality.<br>Breast cancer mortality. |
| Notes | Mammography in control group: 18% during last two years.<br>The Swedish Cancer Society supported this study. |

Risk of bias
| Bias | Authors' judgement | Support for judgement |
| --- | --- | --- |
| Random sequence generation (selection bias) | High risk | Day of birth used. Randomisation ratios varied, not clear whether this was taken into account in the analysis. |
| Allocation concealment (selection bias) | High risk | Day of birth. |
| Blinding of participants and personnel (performance bias)<br>All outcomes | Low risk | Not possible for a screening trial and not relevant. |
| Blinding of outcome assessment (detection bias)<br>All outcomes | Low risk | Blinding of outcome assessment. |
| Incomplete outcome data (attrition bias)<br>All outcomes | Low risk | Women with previous breast cancer were excluded after randomisation. |
| Selective reporting (reporting bias) | Low risk | We found no evidence for this. |
| Other bias | High risk | The whole control group was invited to screening when the trial ended, which renders follow-up data unreliable. |

#### Göteborg, 1982a

Study characteristics
|  |  |
| --- | --- |
| Methods | Individual randomisation within year of birth cohort - by day of birth in the cohorts 1923-1935 and by computer software for the cohorts 1936-1944 - randomisation ratio varied by cohort, on average approximately 1:1.2 (see also text).<br>Blinding of outcome assessment. |
| Participants | Women aged 39-49 years.<br>Number of women randomised: 11,792 to screening, 14,321 to control (see also text).<br>68 women (0.6%) excluded from the screening group and 104 (0.7%) from the control group due to a history of breast carcinoma prior to randomisation. |
| Interventions | Two-view mammography at first screen, single at later rounds - single read at first three rounds; double read thereafter.<br>5 cycles with an interval of 18 months.<br>Control group: usual care. |
| Outcomes | Total mortality.<br>Breast cancer mortality. |
| Notes | Attendance rate: 85%, 78%, 79%, 77%, 75% in rounds 1-5.<br>66% at first screen in control group.<br>Mammography in control group: 19% during last two years; 51% ever.<br>Early systematic screening of control group. |

Risk of bias
| Bias | Authors' judgement | Support for judgement |
| --- | --- | --- |
| Random sequence generation (selection bias) | Unclear risk | See Göteborg 1982. |
| Allocation concealment (selection bias) | Unclear risk | See Göteborg 1982. |
| Blinding of participants and personnel (performance bias) All outcomes | Low risk | See Göteborg 1982. |
| Blinding of outcome assessment (detection bias) All outcomes | Low risk | See Göteborg 1982. |
| Incomplete outcome data (attrition bias) All outcomes | Low risk | See Göteborg 1982. |
| Selective reporting (reporting bias) | Low risk | See Göteborg 1982. |
| Other bias | High risk | See Göteborg 1982. |

#### Göteborg, 1982b

Göteborg 1982b
| Study characteristics |  |  |
| --- | --- | --- |
| Methods | Individual randomisation by computer software - randomisation ratio varied by cohort, on average approximately 1:1.6.<br>Blinding of outcome assessment. |  |
| Participants | Women aged 50-59 years.<br>Number of women randomised not stated explicitly, but can be calculated by comparing two trial reports (see Göteborg 1992 above for total numbers). |  |
| Interventions | Two-view mammography at first screen, single at later rounds - single read at first three rounds; double read thereafter.<br>4 cycles with an interval of 18 months.<br>Control group: usual care. |  |
| Outcomes | Total mortality.<br>Breast cancer mortality. |  |
| Notes | Attendance rate: 83% at first screen.<br>78% at first screen in control group.<br>Early systematic screening of control group. |  |
| Risk of bias |  |  |
| Bias | Authors' judgement | Support for judgement |
| Random sequence generation (selection bias) | Unclear risk | See Göteborg 1982. |
| Allocation concealment (selection bias) | Unclear risk | See Göteborg 1982. |
| Blinding of participants and personnel (performance bias)<br>All outcomes | Low risk | See Göteborg 1982. |
| Blinding of outcome assessment (detection bias)<br>All outcomes | Low risk | See Göteborg 1982. |
| Incomplete outcome data (attrition bias)<br>All outcomes | Low risk | See Göteborg 1982. |
| Selective reporting (reporting bias) | Low risk | See Göteborg 1982. |
| Other bias | High risk | See Göteborg 1982. |

#### Kopparberg, 1977

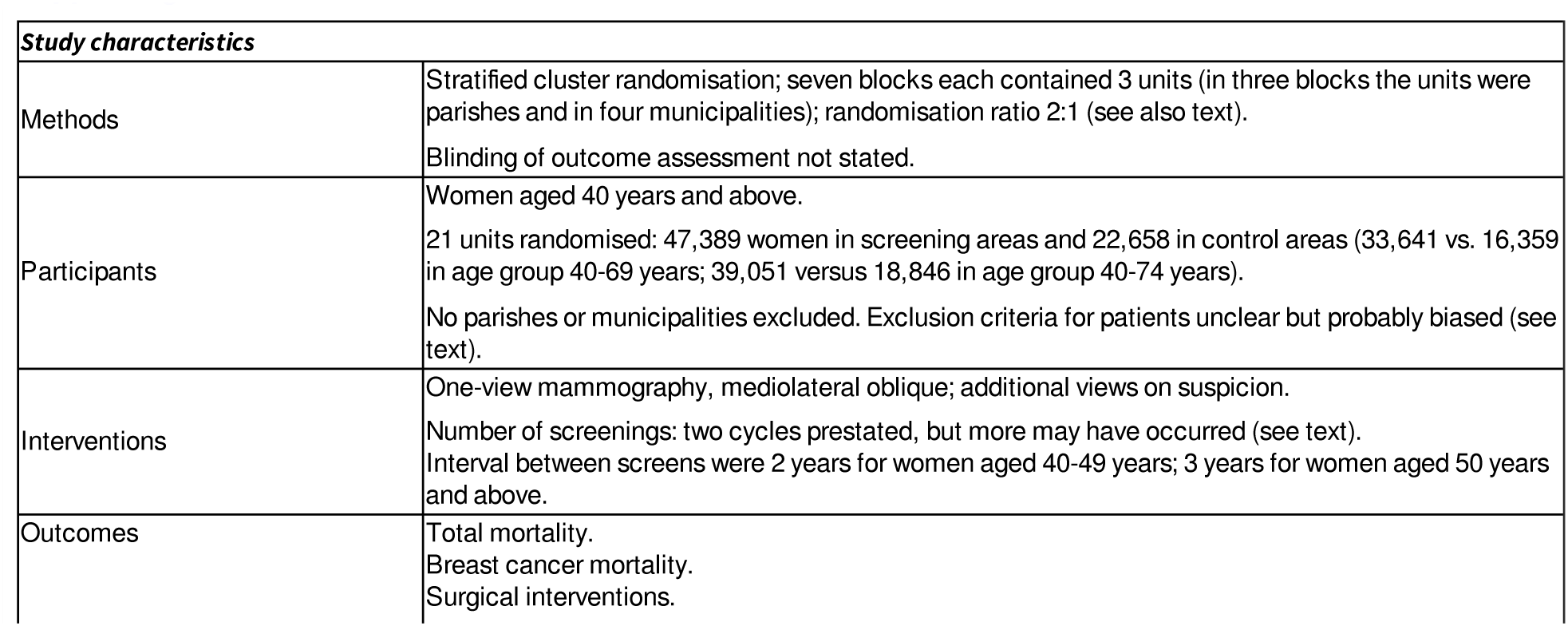
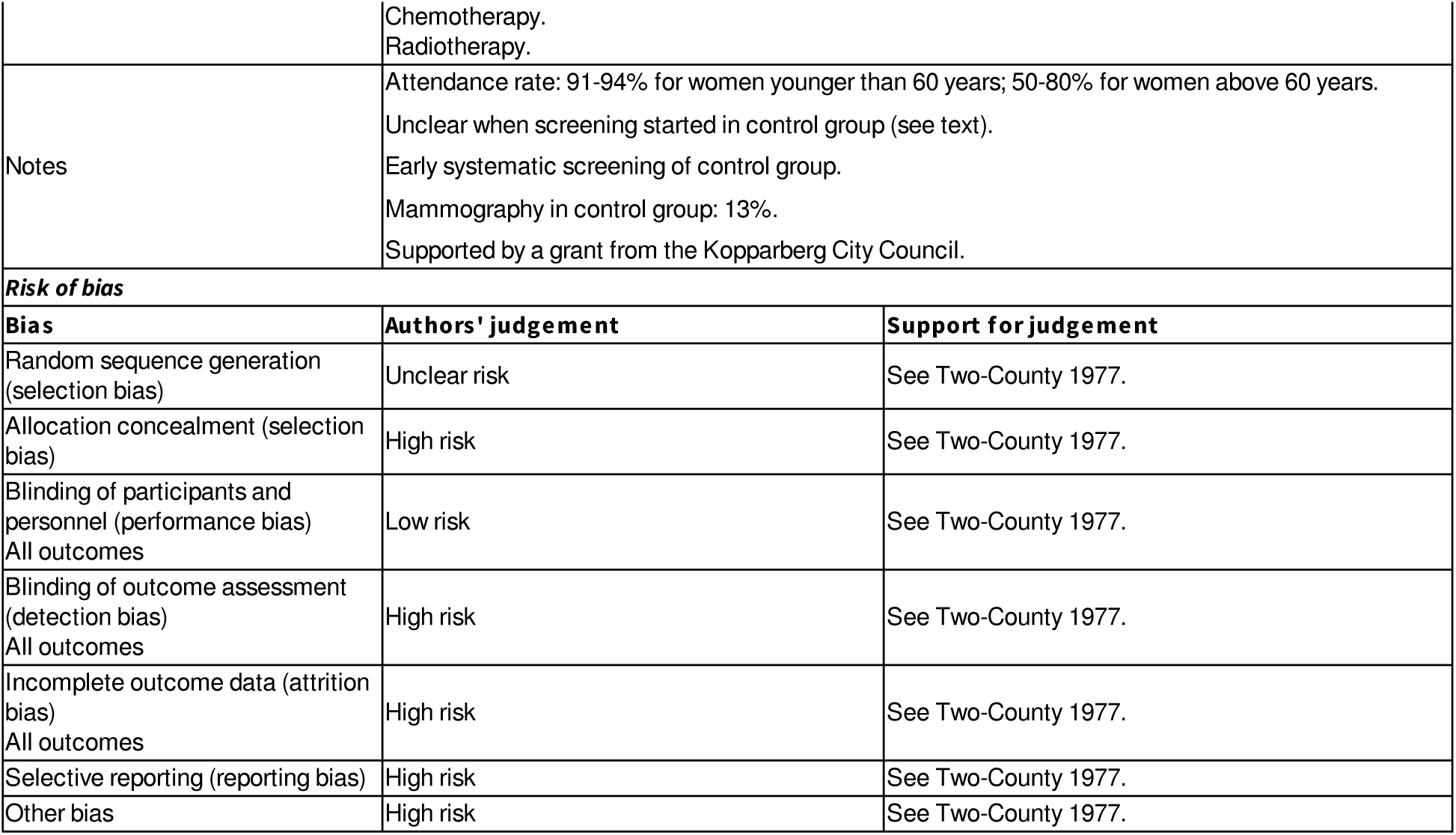

#### Malmö, 1976

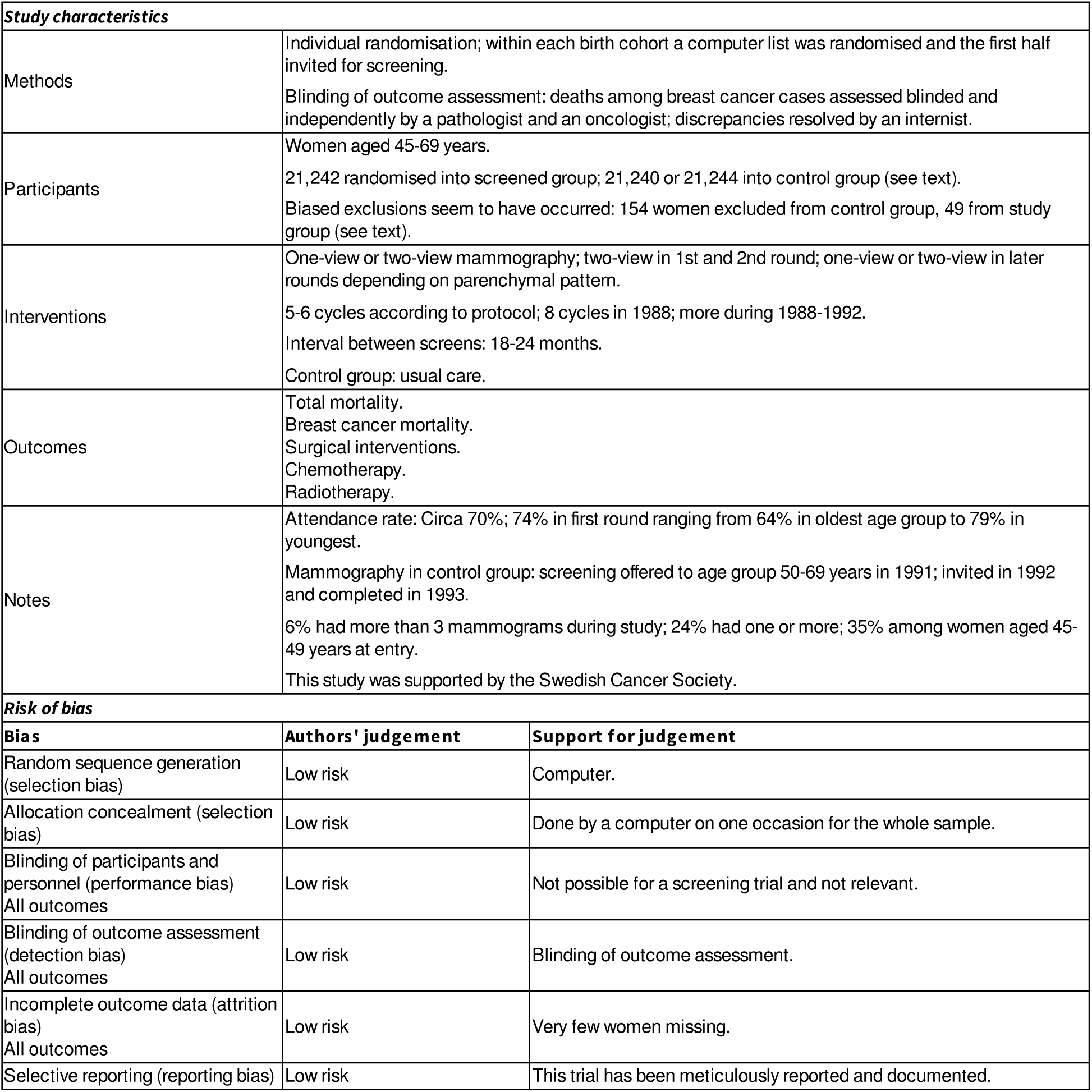
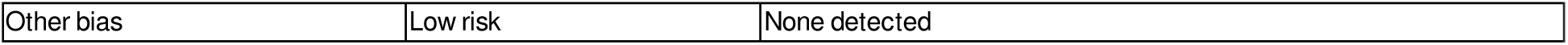

Malmö 1976
| Study characteristics |  |  |
| --- | --- | --- |
| Methods | Individual randomisation; within each birth cohort a computer list was randomised and the first half invited for screening.<br>Blinding of outcome assessment: deaths among breast cancer cases assessed blinded and independently by a pathologist and an oncologist; discrepancies resolved by an internist. |  |
| Participants | Women aged 45-69 years.<br>21,242 randomised into screened group; 21,240 or 21,244 into control group (see text).<br>Biased exclusions seem to have occurred: 154 women excluded from control group, 49 from study group (see text). |  |
| Interventions | One-view or two-view mammography; two-view in 1st and 2nd round; one-view or two-view in later rounds depending on parenchymal pattern.<br>5-6 cycles according to protocol; 8 cycles in 1988; more during 1988-1992.<br>Interval between screens: 18-24 months.<br>Control group: usual care. |  |
| Outcomes | Total mortality.<br>Breast cancer mortality.<br>Surgical interventions.<br>Chemotherapy.<br>Radiotherapy. |  |
| Notes | Attendance rate: Circa 70%; 74% in first round ranging from 64% in oldest age group to 79% in youngest.<br>Mammography in control group: screening offered to age group 50-69 years in 1991; invited in 1992 and completed in 1993.<br>6% had more than 3 mammograms during study; 24% had one or more; 35% among women aged 45-49 years at entry.<br>This study was supported by the Swedish Cancer Society. |  |
| Risk of bias |  |  |
| Bias | Authors' judgement | Support for judgement |
| Random sequence generation (selection bias) | Low risk | Computer. |
| Allocation concealment (selection bias) | Low risk | Done by a computer on one occasion for the whole sample. |
| Blinding of participants and personnel (performance bias)<br>All outcomes | Low risk | Not possible for a screening trial and not relevant. |
| Blinding of outcome assessment (detection bias)<br>All outcomes | Low risk | Blinding of outcome assessment. |
| Incomplete outcome data (attrition bias)<br>All outcomes | Low risk | Very few women missing. |
| Selective reporting (reporting bias) | Low risk | This trial has been meticulously reported and documented. |
| Other bias | Low risk | None detected |

#### Malmö II, 1978

Malmö II 1978
| Study characteristics |  |  |
| --- | --- | --- |
| Methods | See text of the review; extension of Malmö 1976. |  |
| Participants |  |  |
| Interventions |  |  |
| Outcomes |  |  |
| Notes |  |  |
| Risk of bias |  |  |
| Bias | Authors' judgement | Support for judgement |
| Random sequence generation (selection bias) | High risk | See text of the review; extension of Malmö 1976, not done according to a formal protocol, inclusion criteria violated, group sizes differed although they should have been the same, and gross and unexplained imbalance in numbers in the two groups. |
| Allocation concealment (selection bias) | High risk | See 'Random sequence generation.' |
| Blinding of participants and personnel (performance bias)<br>All outcomes | Low risk | Not possible for a screening trial and not relevant. |
| Blinding of outcome assessment (detection bias)<br>All outcomes | High risk | See 'Random sequence generation.' |
| Incomplete outcome data (attrition bias)<br>All outcomes | High risk | See 'Random sequence generation.' |
| Selective reporting (reporting bias) | High risk | See 'Random sequence generation.' |
| Other bias | High risk | See 'Random sequence generation.' |

#### New York, 1963

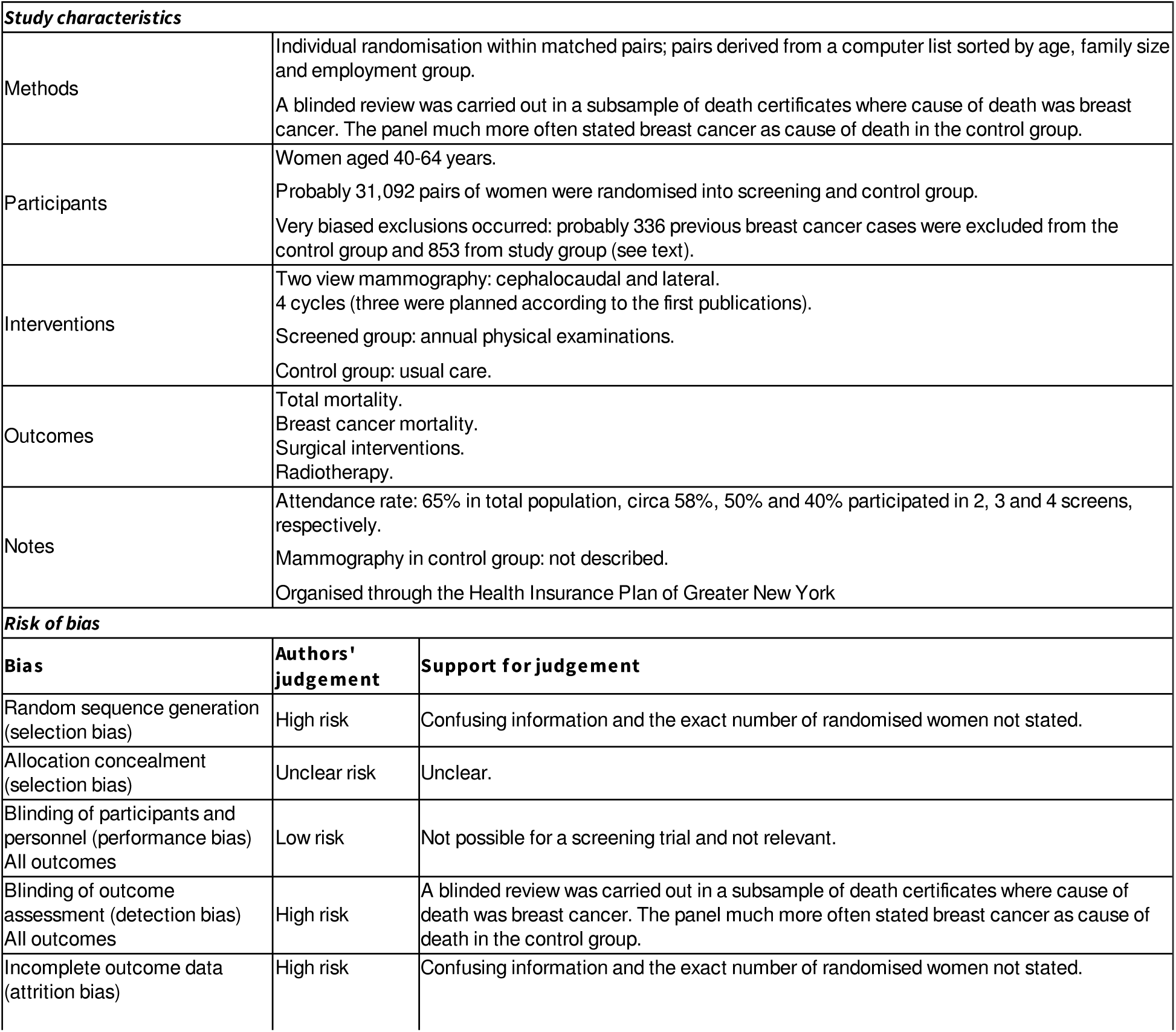
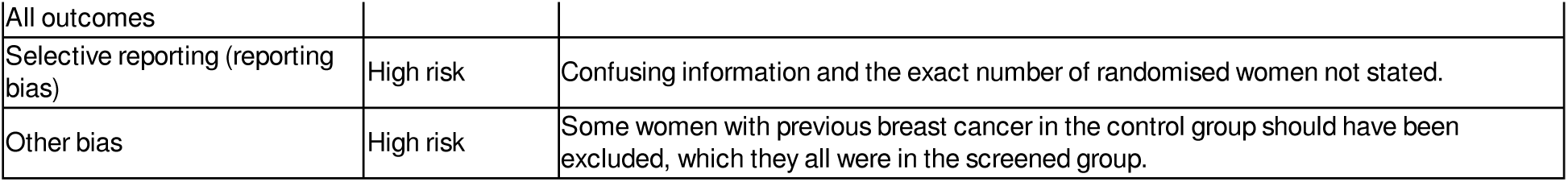

#### Stockholm, 1981

Study characteristics
|  |  |
| --- | --- |
| Methods | Individual randomisation by day of birth; 1-10 and 21-31 in study group and 11-20 in control group (see also text).<br>Blinding of outcome assessment: not stated. |
| Participants | Women aged 40-64 years.<br>Number of women randomised inconsistently reported (see text).<br>Exclusions after randomisation unclear (see text). |
| Interventions | Single oblique mammography; recalled for conventional three-view if malignancies suspected.<br>2 cycles (number not predetermined - screening introduced in control group because of results from Kopparberg).<br>Interval between screens: Circa 2 years; 2.5 years to complete first round and 2.1 to complete second round.<br>Control group: usual care. |
| Outcomes | Total mortality.<br>Breast cancer mortality.<br>Surgical interventions. |
| Notes | Attendance rate: circa 80%.<br>Mammography in control group: 8% during one year; 25% in study group during two years previous to screening.<br>Early systematic screening of control group.<br>Likely publicly funded. |

Risk of bias
| Bias | Authors' judgement | Support for judgement |
| --- | --- | --- |
| Random sequence generation (selection bias) | High risk | Day of birth. |
| Allocation concealment (selection bias) | High risk | Day of birth. |
| Blinding of participants and personnel (performance bias)<br>All outcomes | Low risk | Not possible for a screening trial and not relevant. |
| Blinding of outcome assessment (detection bias)<br>All outcomes | High risk | Blinding of outcome assessment not stated. |
| Incomplete outcome data (attrition bias)<br>All outcomes | High risk | Reported numbers are inconsistent. |
| Selective reporting (reporting bias) | High risk | Reported numbers are inconsistent. |
| Other bias | High risk | Reported numbers are inconsistent. |

#### Two-County, 1977

Study characteristics
|  |  |
| --- | --- |
| Methods | Stratified cluster randomisation (see Kopparberg 1977 and Östergötland 1978 for details).<br>Blinding of cause of death assessments in some later updates for use in Swedish meta-analyses. |
| Participants | Women aged 40-74 years.<br>(See Kopparberg 1977 and Östergötland 1978 for details). |
| Interventions | See Kopparberg 1977 and Östergötland 1978.<br>Screened women were encouraged to perform self-examination of the breasts every month.<br>Control women: usual care. |
| Outcomes | See Kopparberg 1977 and Östergötland 1978. |
| Notes | See Kopparberg 1977 and Östergötland 1978.<br>Funded by the Swedish National Board of Health and Welfare. |

Risk of bias
| Bias | Authors' judgement | Support for judgement |
| --- | --- | --- |
| Random sequence generation (selection bias) | Unclear risk | No information. |
| Allocation concealment (selection bias) | High risk | See text, information inconsistent and incomplete. |
| Blinding of participants and personnel (performance bias)<br>All outcomes | Low risk | Not possible for a screening trial and not relevant. |
| Blinding of outcome assessment (detection bias)<br>All outcomes | High risk | Blinding of outcome assessment not stated. |
| Incomplete outcome data (attrition bias)<br>All outcomes | High risk | Numbers of women, cancers and deaths vary in the reports of the trial. |
| Selective reporting (reporting bias) | High risk | Numbers of women, cancers and deaths vary in the reports of the trial. |
| Other bias | High risk | Numbers of women, cancers and deaths vary in the reports of the trial, see also main text. |

#### UK age trial, 1991

UK age trial 1991
| Study characteristics |  |  |
| --- | --- | --- |
| Methods | Individual randomisation by computer; randomisation ratio 1:2.<br>Information on cause of death was obtained from the central register of the National Health Service. |  |
| Participants | Women aged 39-41 years.<br>53,914 randomised into screened group; 107,007 into control group.<br>30 and 51 excluded after randomisation. |  |
| Interventions | Two-view mammography at first screen, and by single mediolateral oblique view thereafter, with recall for full assessment if an abnormality was suspected.<br>7 annual screens planned.<br>Control group: usual care. |  |
| Outcomes | Total mortality.<br>Breast cancer mortality. |  |
| Notes | Number of cancers in latest report given per 1000 women-years.<br>Participation rate: ca 66% at prevalence screen, below 50% at 8th screen.<br>Funding through the National Institute for Health Research Health Technology Assessment programme. |  |
| Risk of bias |  |  |
| Bias | Authors' judgement | Support for judgement |
| Random sequence generation (selection bias) | Low risk | Computer. |
| Allocation concealment (selection bias) | Low risk | Individual randomisation by computer. |
| Blinding of participants and personnel (performance bias)<br>All outcomes | Low risk | Not possible for a screening trial and not relevant. |
| Blinding of outcome assessment (detection bias)<br>All outcomes | Low risk | Information on cause of death was obtained from the central register of the National Health Service. |
| Incomplete outcome data (attrition bias)<br>All outcomes | Low risk | Very few women excluded after randomisation. |
| Selective reporting (reporting bias) | Low risk | We found no evidence for this |
| Other bias | Low risk | We found no evidence for this |

#### Östergötland, 1978

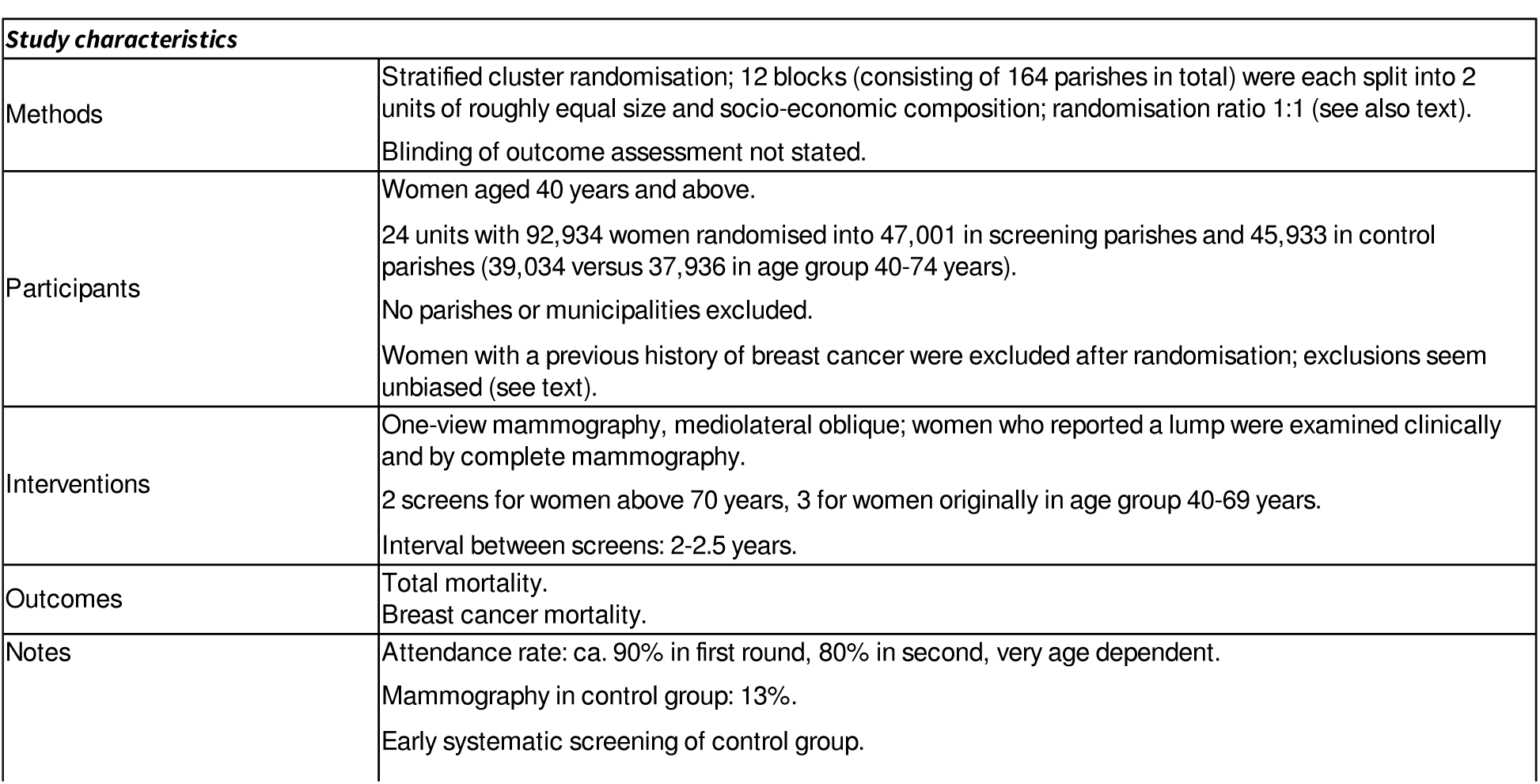
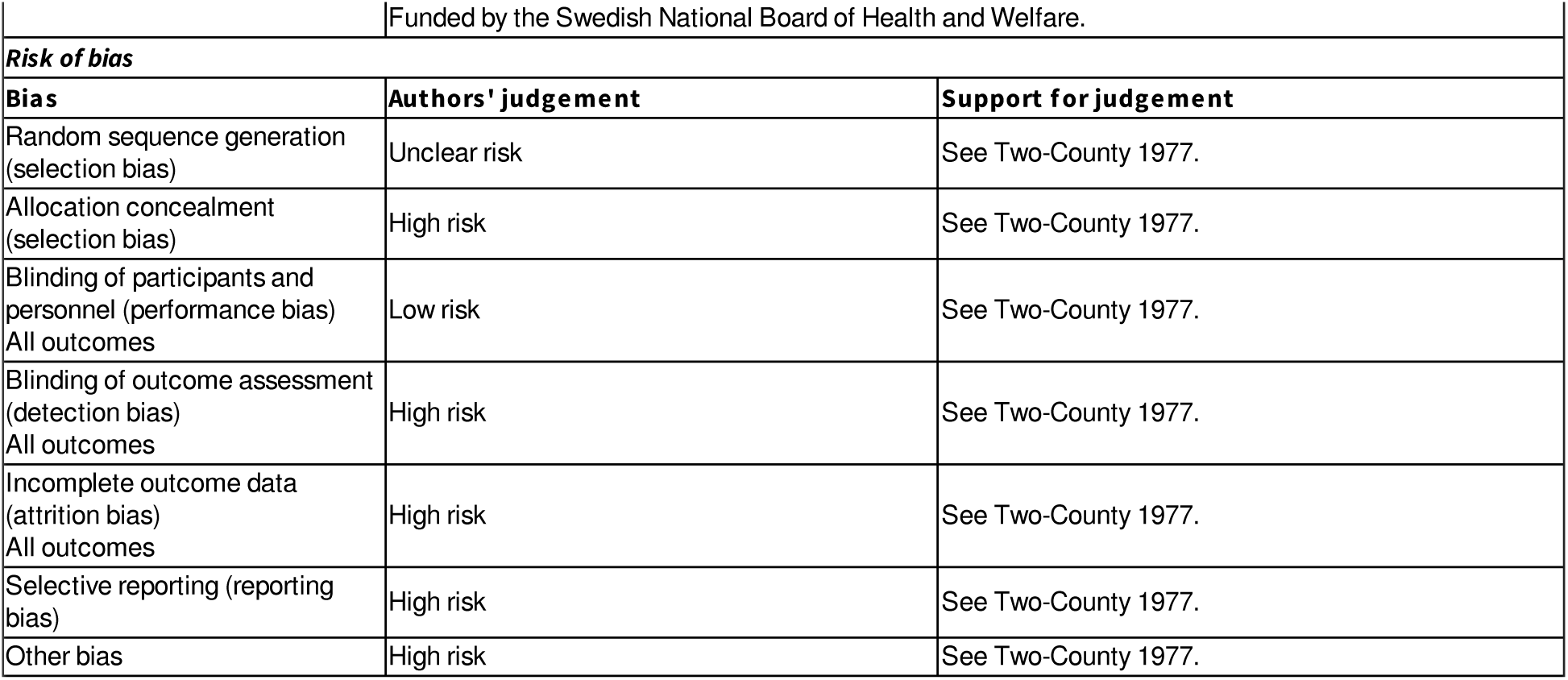

### Characteristics of excluded studies [ordered by study ID]

| Study | Reason for exclusion |
| --- | --- |
| Berglund 2000 | Multiple risk factor intervention study, with several interventions, including mammography, not a randomised trial but alternating allocation of birth year cohorts with resulting age differences at baseline between the two groups; 50 women died from cancer of 8,712 participants, no data on breast cancer. |
| Dales 1979 | Multiple risk factor intervention trial, with several interventions, regular mammography was only one of the interventions and only about 1000 women were invited for mammography. |
| Singapore 1994 | Singapore Breast Screening Project. Randomised 166,600 women aged 50-64 years, but the only intervention was the prevalence screen, and exclusions after randomisation occurred only in the screened group. Previous cancer at any site was an exclusion criterion; more than 1500 women were excluded from the screened group, 468 because they were already dead. |

### Characteristics of studies awaiting classification [ordered by study ID]

#### AgeX Trial

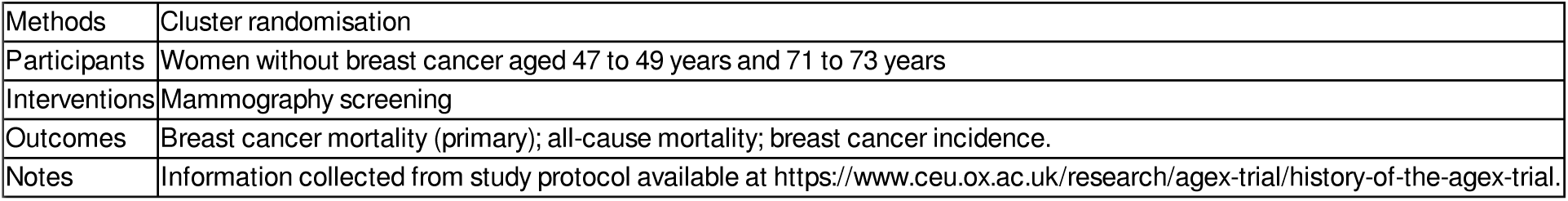

#### Murillo, 2016

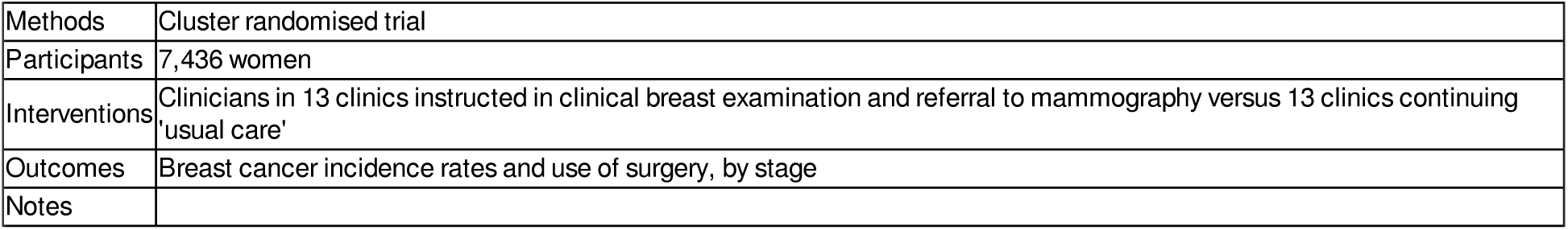

## Appendices

## Appendix 1. PubMed search strategy

#1 (breast neoplasms[MeSH Terms] OR “breast cancer” OR mammography[MeSH Terms] OR mammograph*)

#2 (mass screening[MeSH Terms] OR screen*)

#3 #1 AND #2

#4 (((randomized controlled trial [pt] OR “controlled clinical trial“[Publication Type] OR “randomized“[Title/Abstract] OR “placebo“[Title/Abstract]) OR (“clinical trials as topic” [mesh terms]) OR (randomly [tiab] OR trial [ti])) NOT (animals [mh] NOT humans [mh]))

#5 #3 AND #4

#6 (“2012/11/01“[Date - Publication] : “2023/02/28“[Date - Publication])

#7 #5 AND #6

## Appendix 2. CENTRAL Search Strategy

#1 MeSH descriptor: [Breast Neoplasms] explode all trees

#2 (early breast cancer* or early breast neoplas* or early breast carcinoma* or early breast tumour* or early breast tumor*)

#3 MeSH descriptor: [Mammography] explode all trees

#4 Mammograph*

#5 #1 OR #2 OR #3 OR #4

#6 MeSH descriptor: [Mass Screening] explode all trees

#7 screen*

#8 #6 OR #7

#9 #5 AND #8 with Publication Year from, 2012 to present, in Trials

## Appendix 3. World Health Organization (WHO) International Clinical Trials Registry Platform (ICTRP) Search Strategy

Basic search:

Breast cancer AND mammograph* Advanced search:

1. Condition: breast AND (cancer* OR carcinoma* OR neoplas* OR tumour* OR tumor*) Intervention: mammograph*

Recruitment Status: All

2. Condition: breast AND (cancer* OR carcinoma* OR neoplas* OR tumour* OR tumor*) Intervention: screen*

Recruitment Status: All

## Appendix 4. Clinicaltrials.gov Search Strategy

Basic search:

Condition or disease: Breast cancer

Other terms: mammography

Status: All studies Advanced search:

1. Condition: breast cancer OR breast neoplasm Intervention/treatment: mammography

2. Condition: breast cancer OR breast neoplasm Intervention/treatment: screening

---

**Analysis 1.1.**
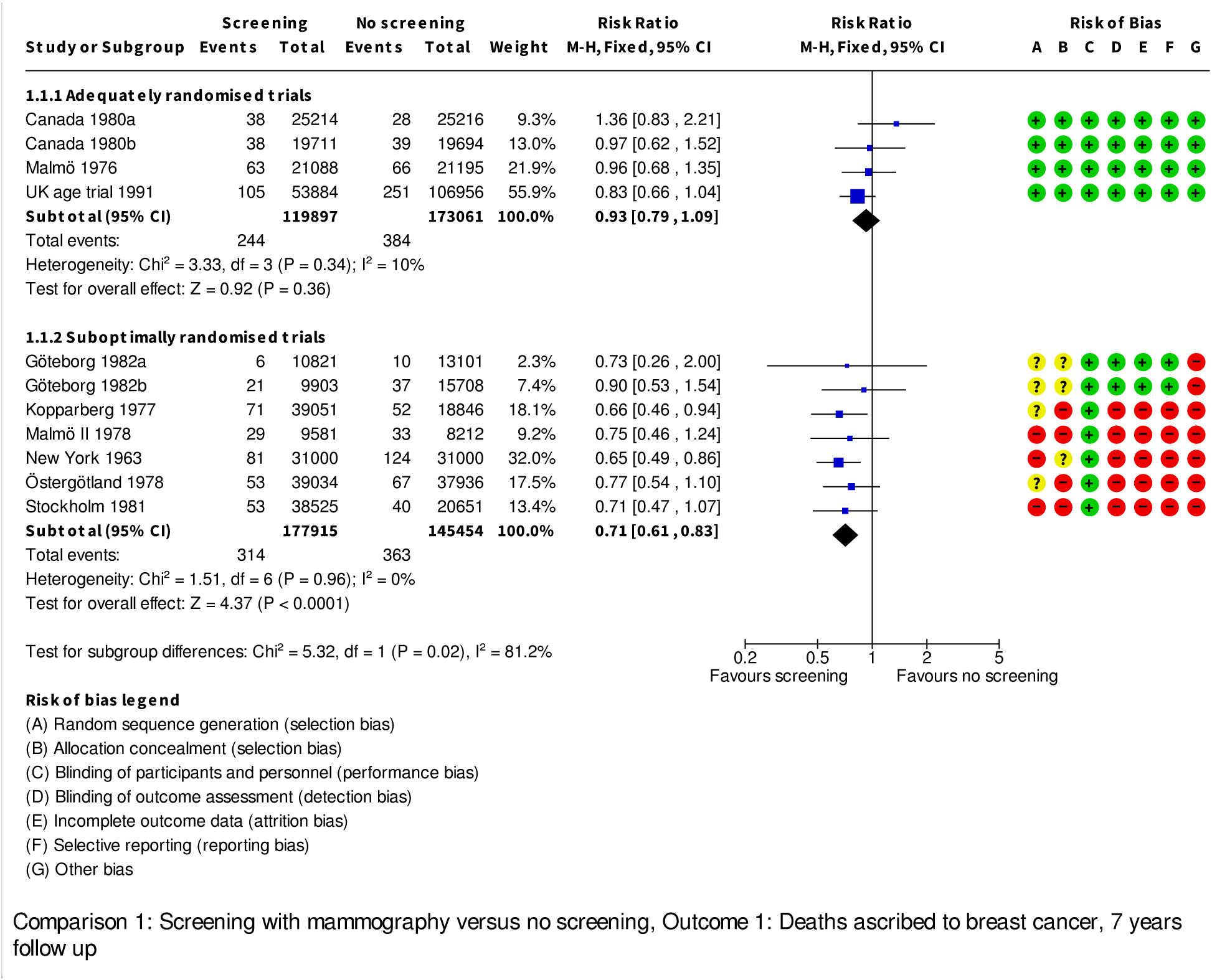

**Analysis 1.2.**
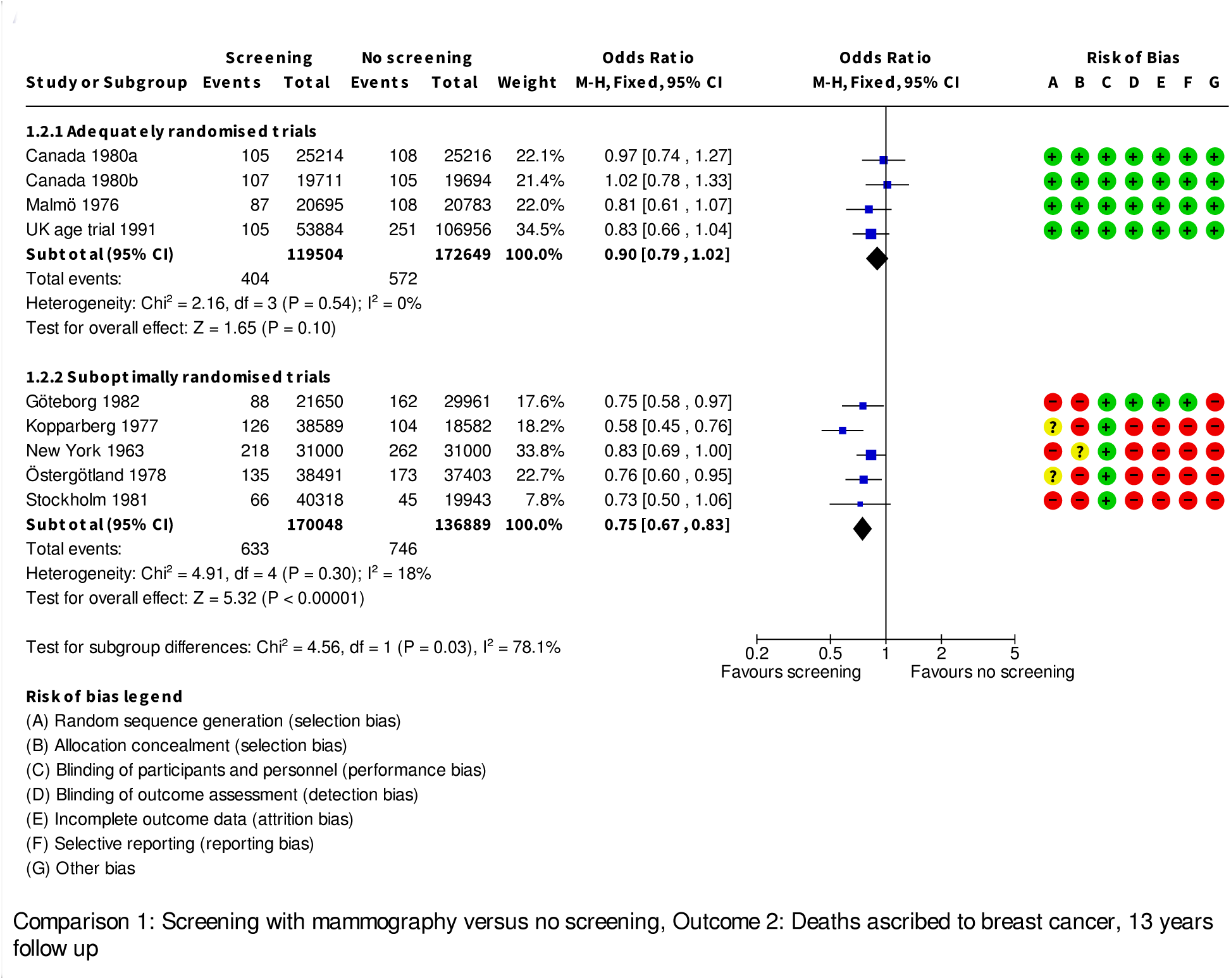

**Analysis 1.3.**
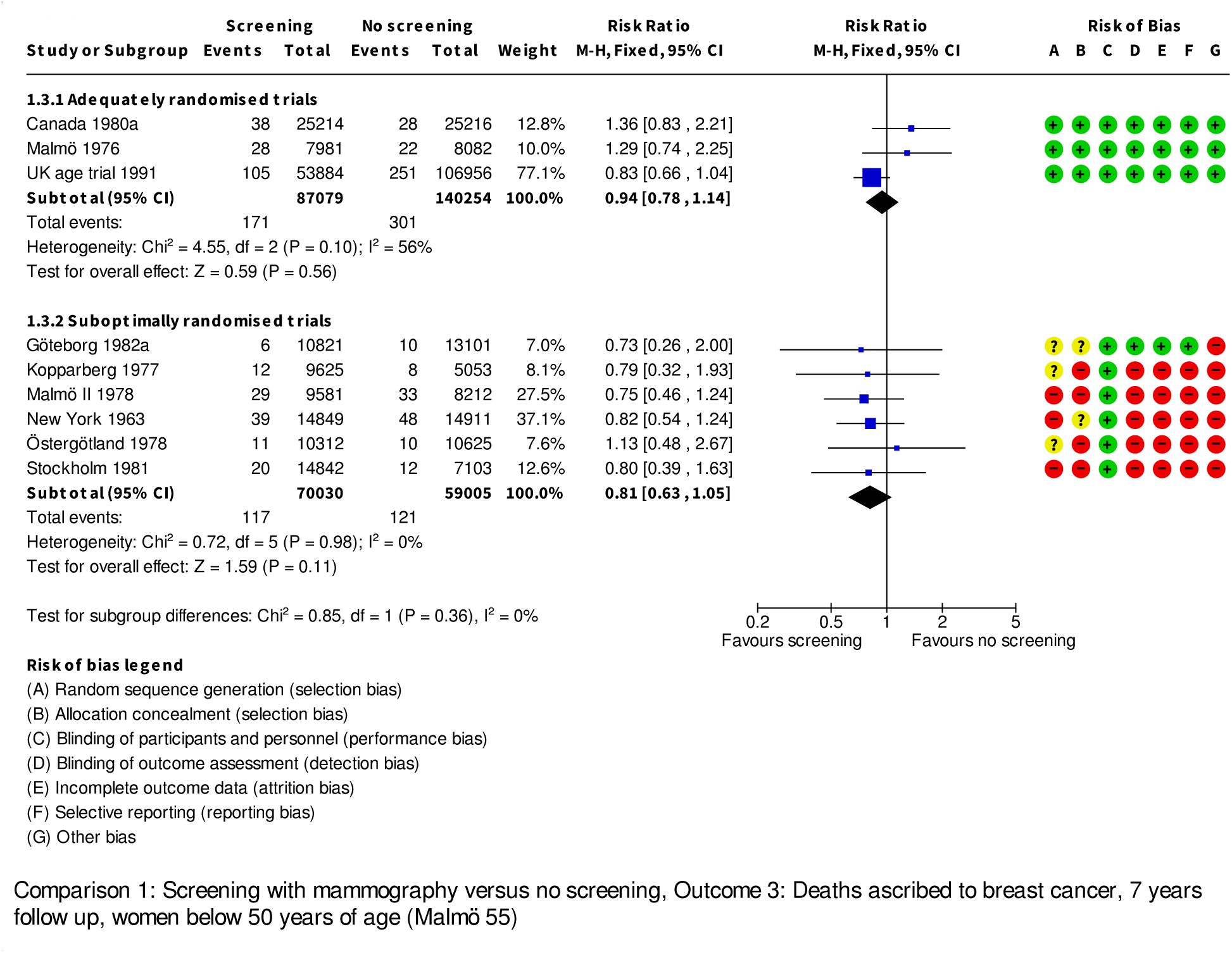

**Analysis 1.4.**
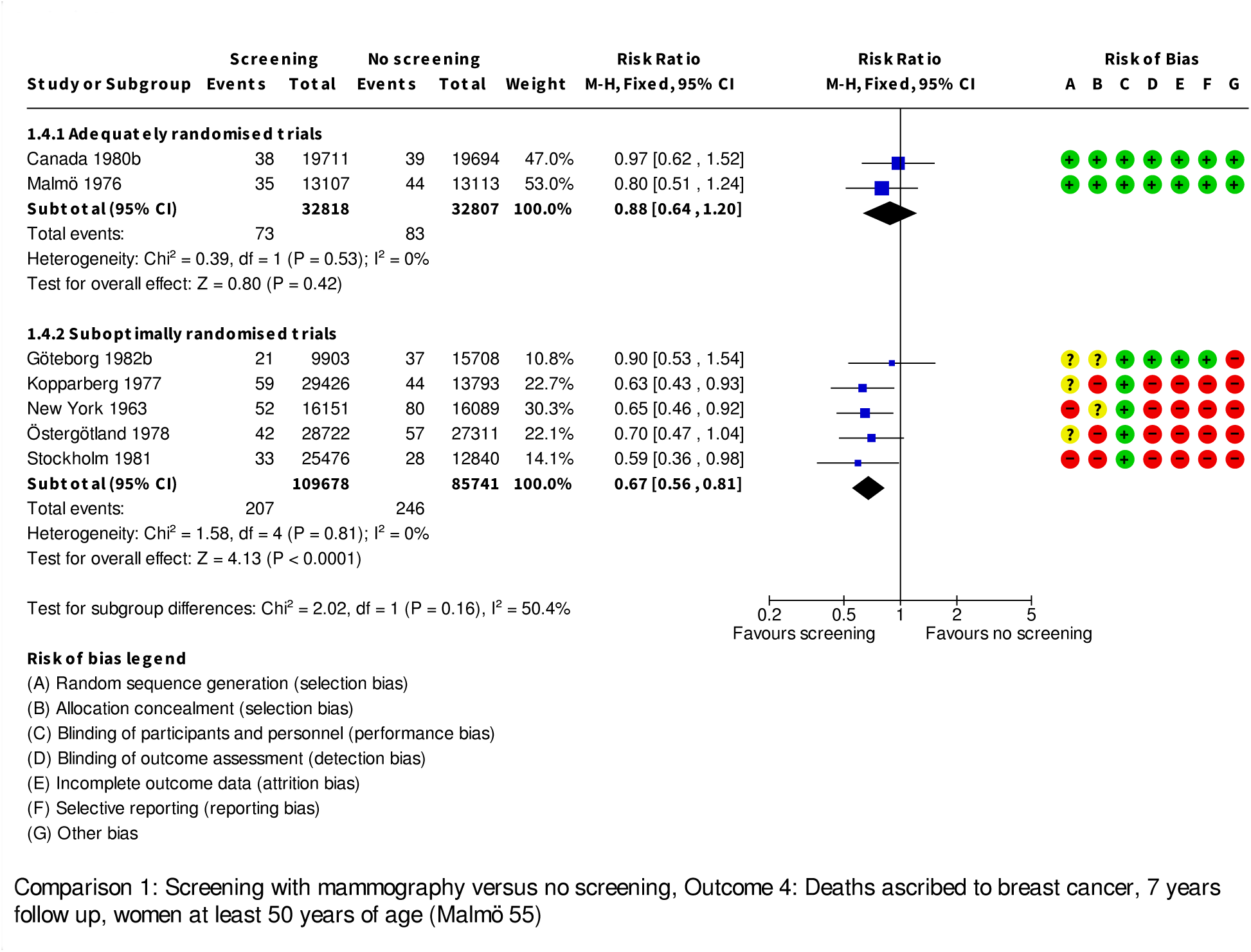

**Analysis 1.5.**
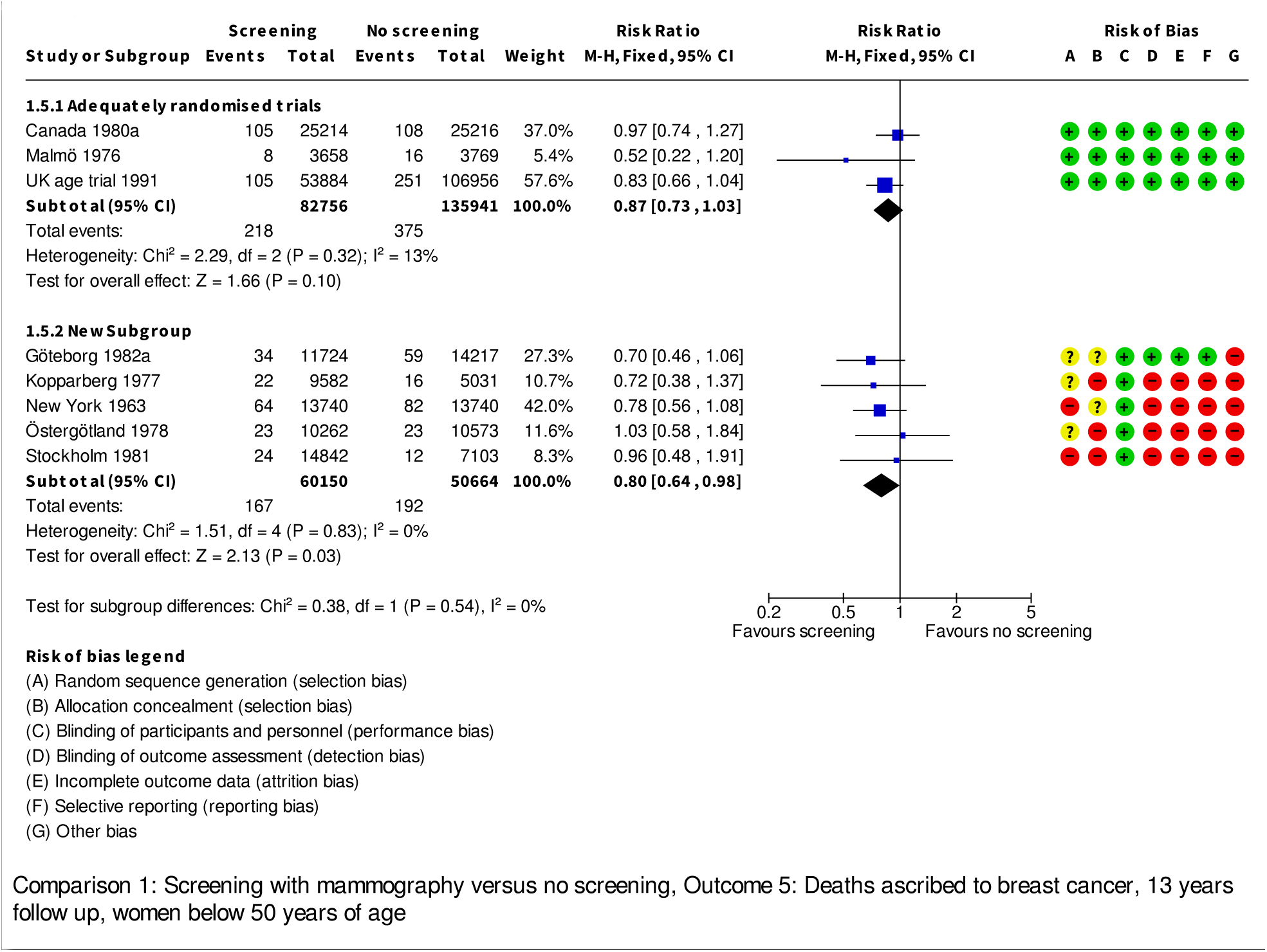

**Analysis 1.6.**
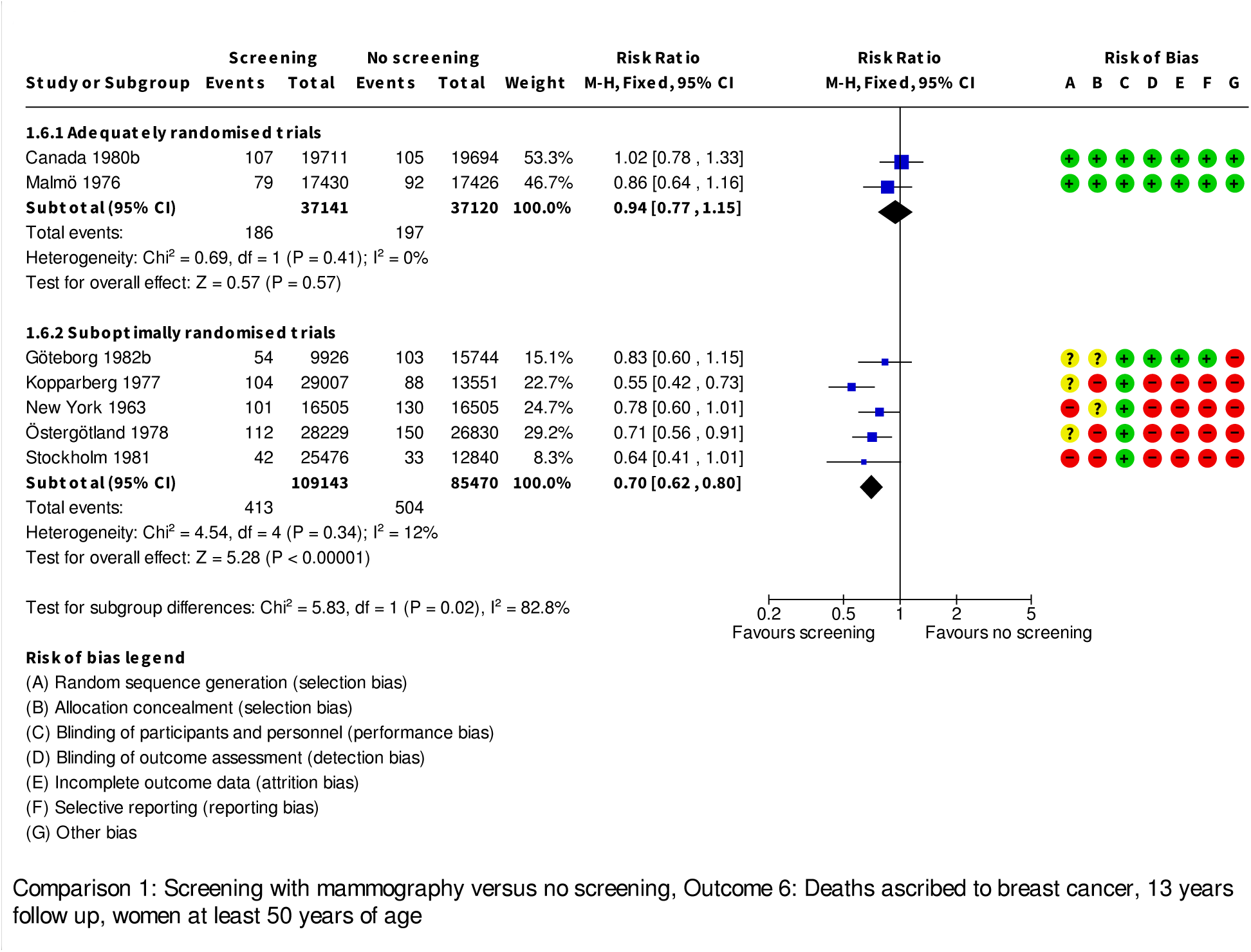

**Analysis 1.7.**
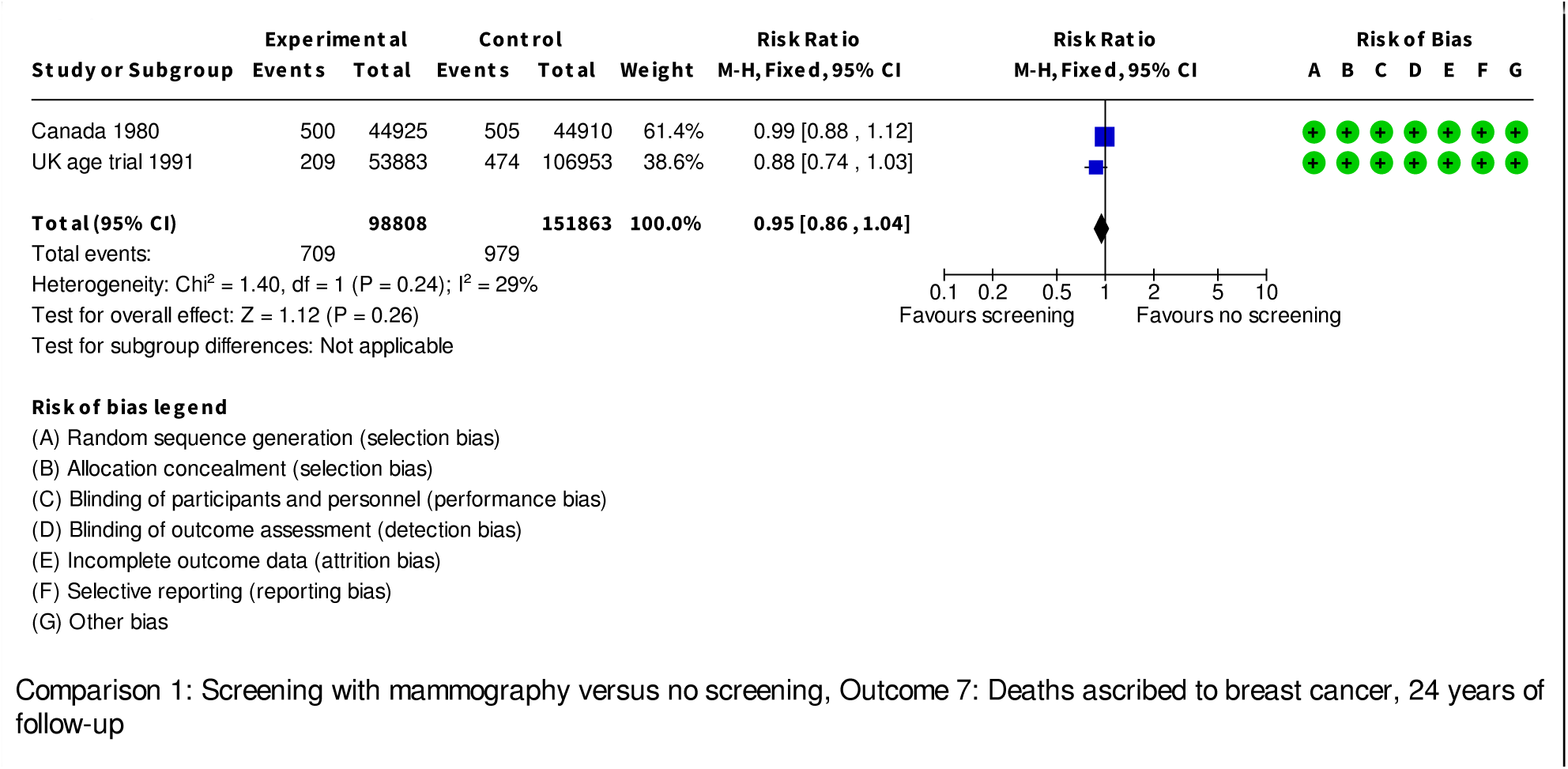

**Analysis 1.8.**
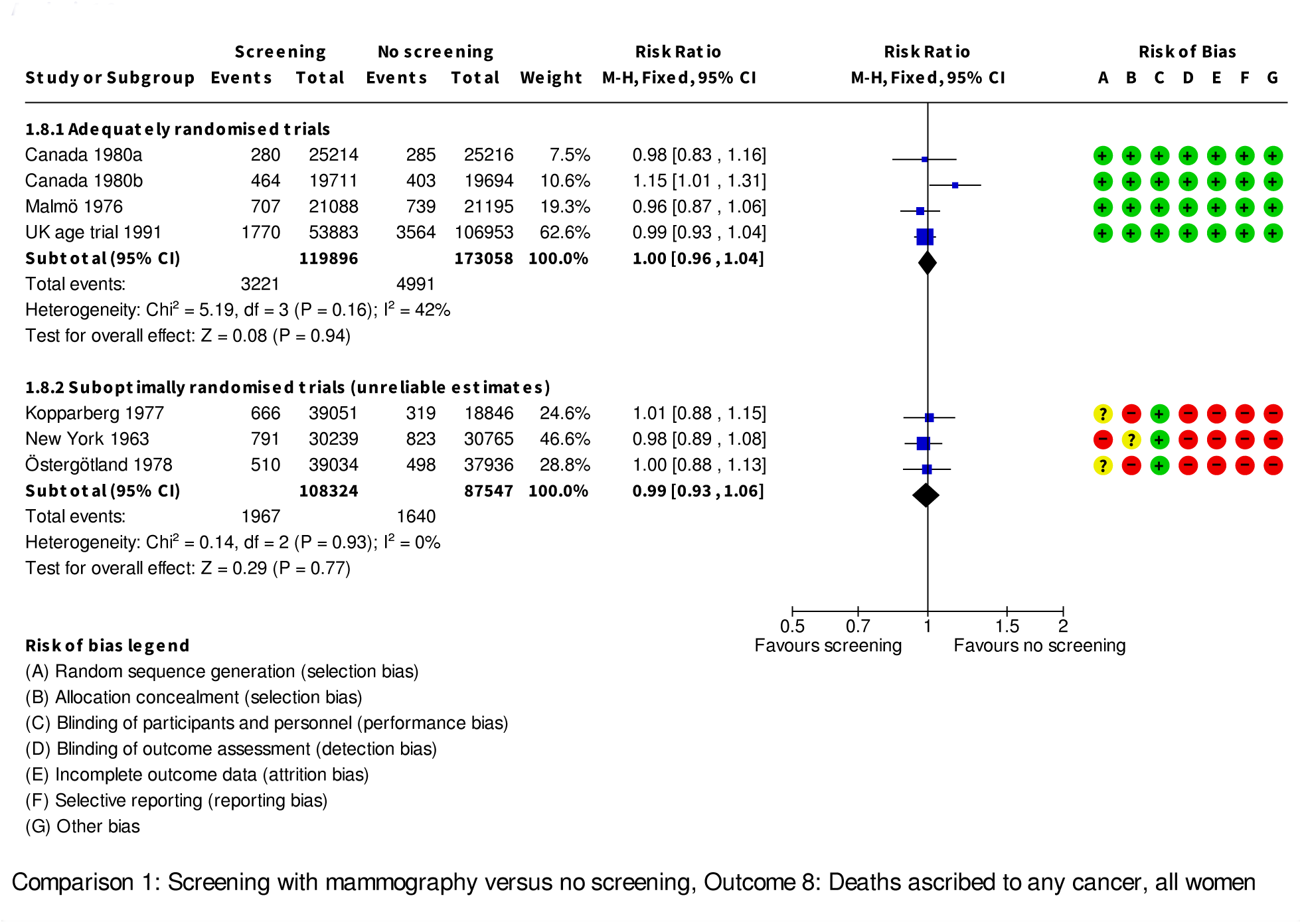

**Analysis 1.9.**
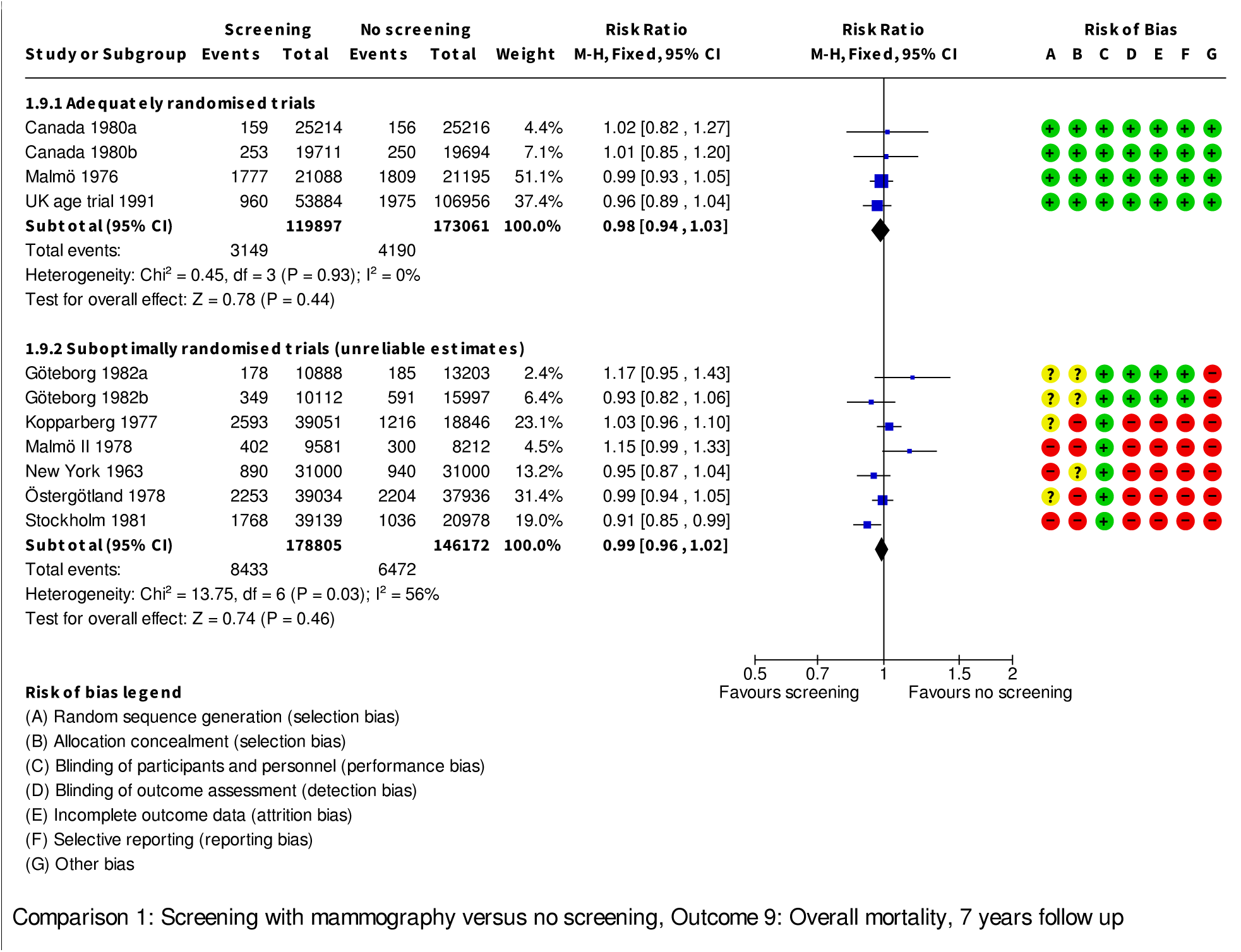

**Analysis 1.10.**
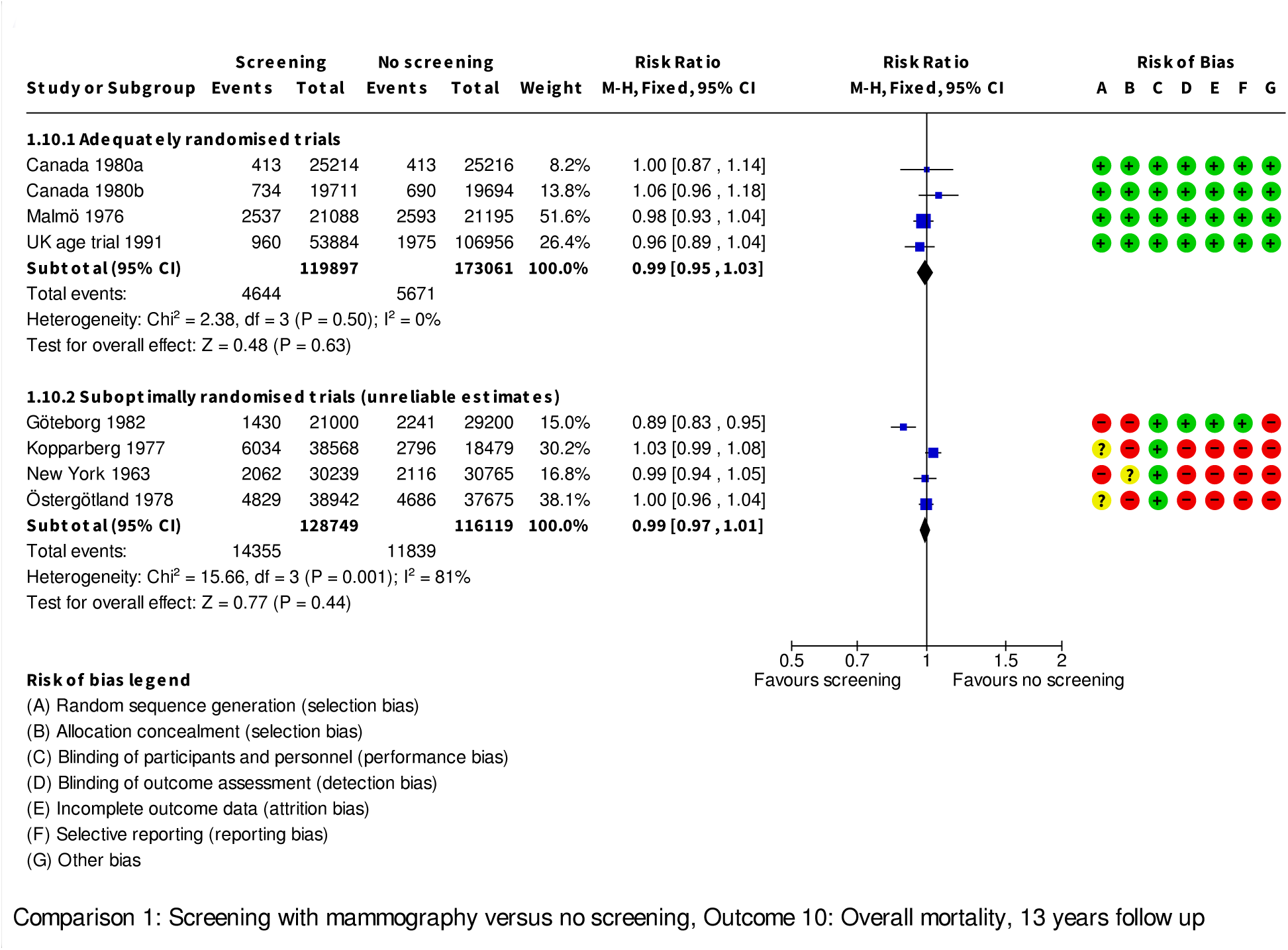

**Analysis 1.11.**
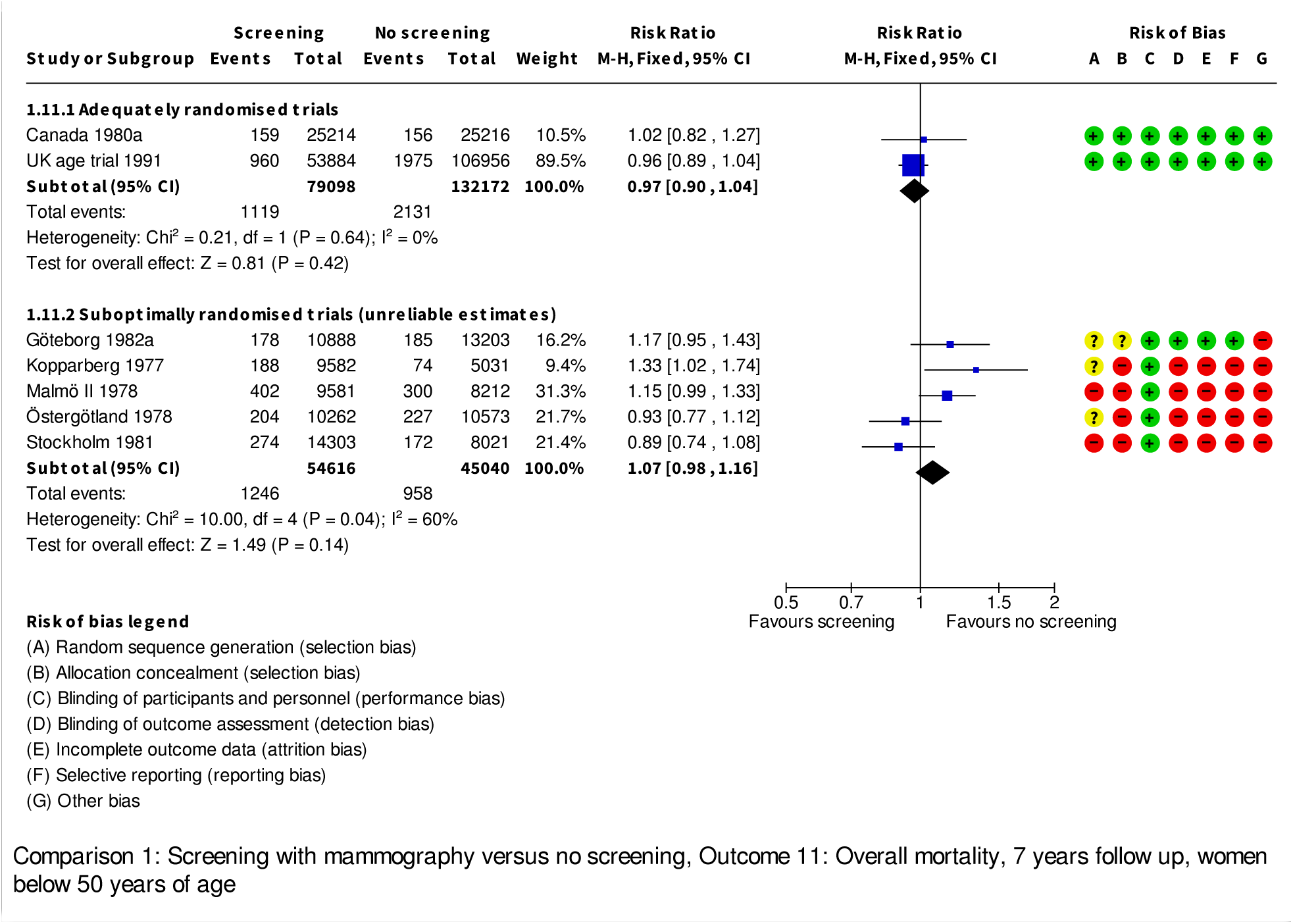

**Analysis 1.12.**
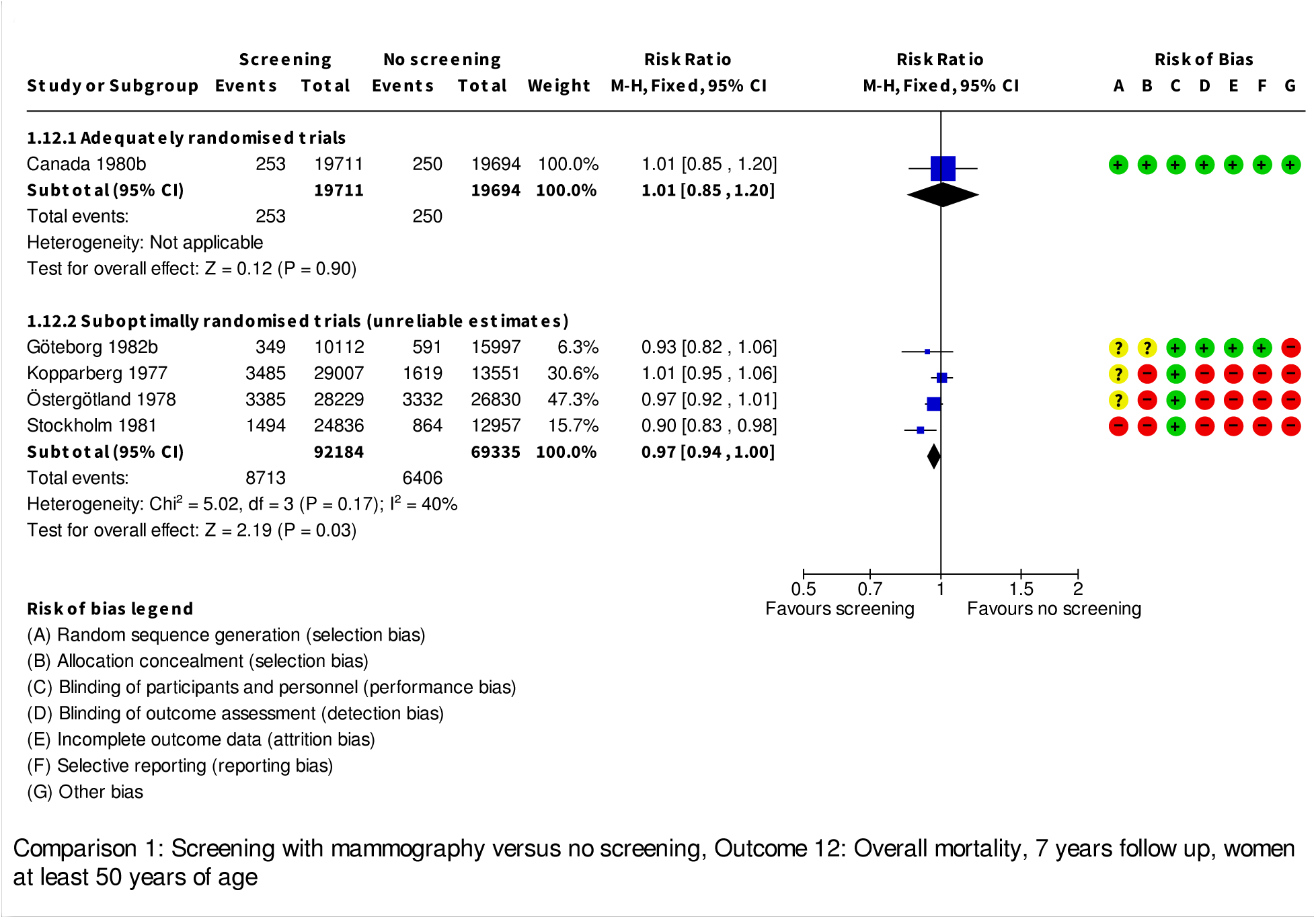

**Analysis 1.13.**
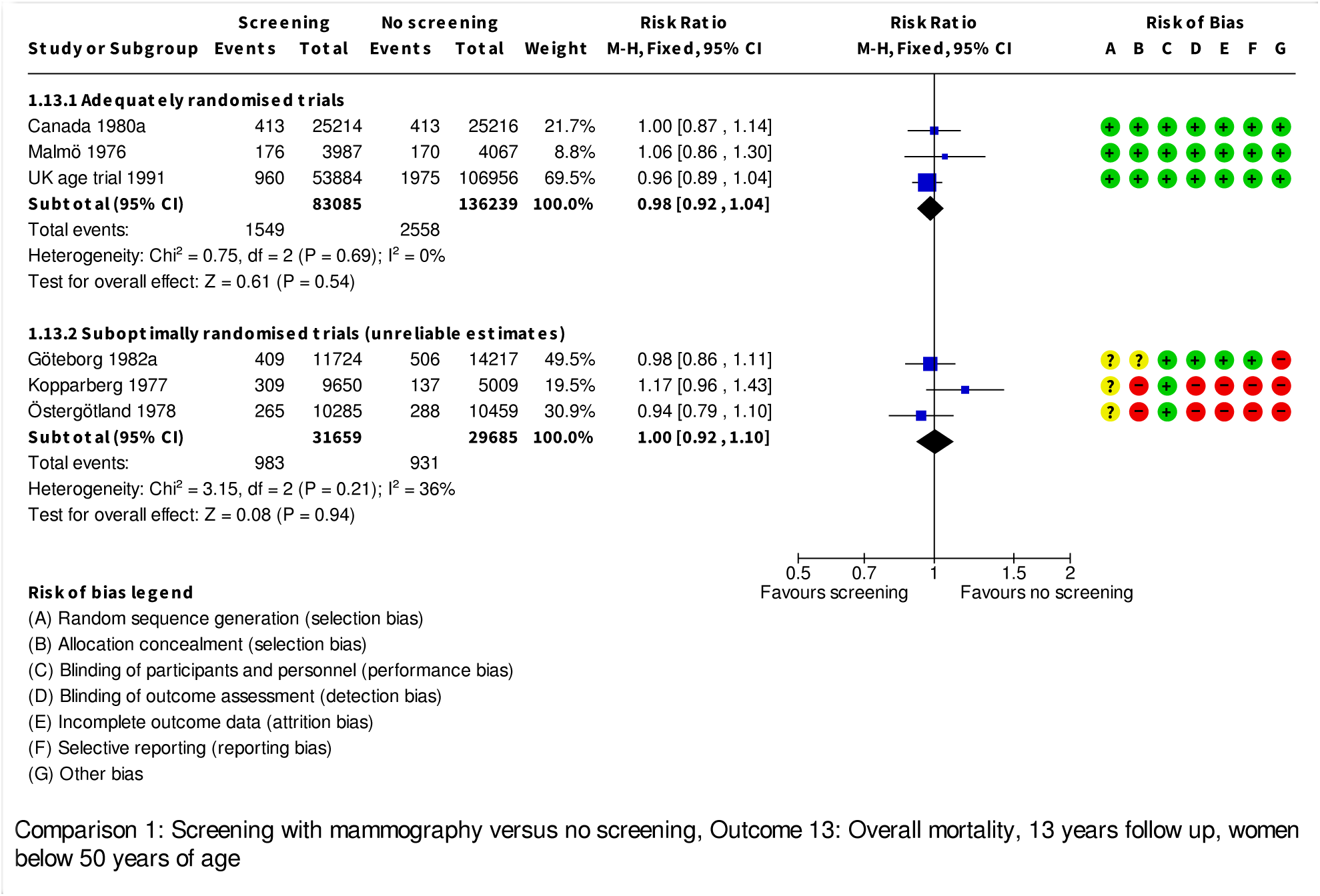

**Analysis 1.14.**
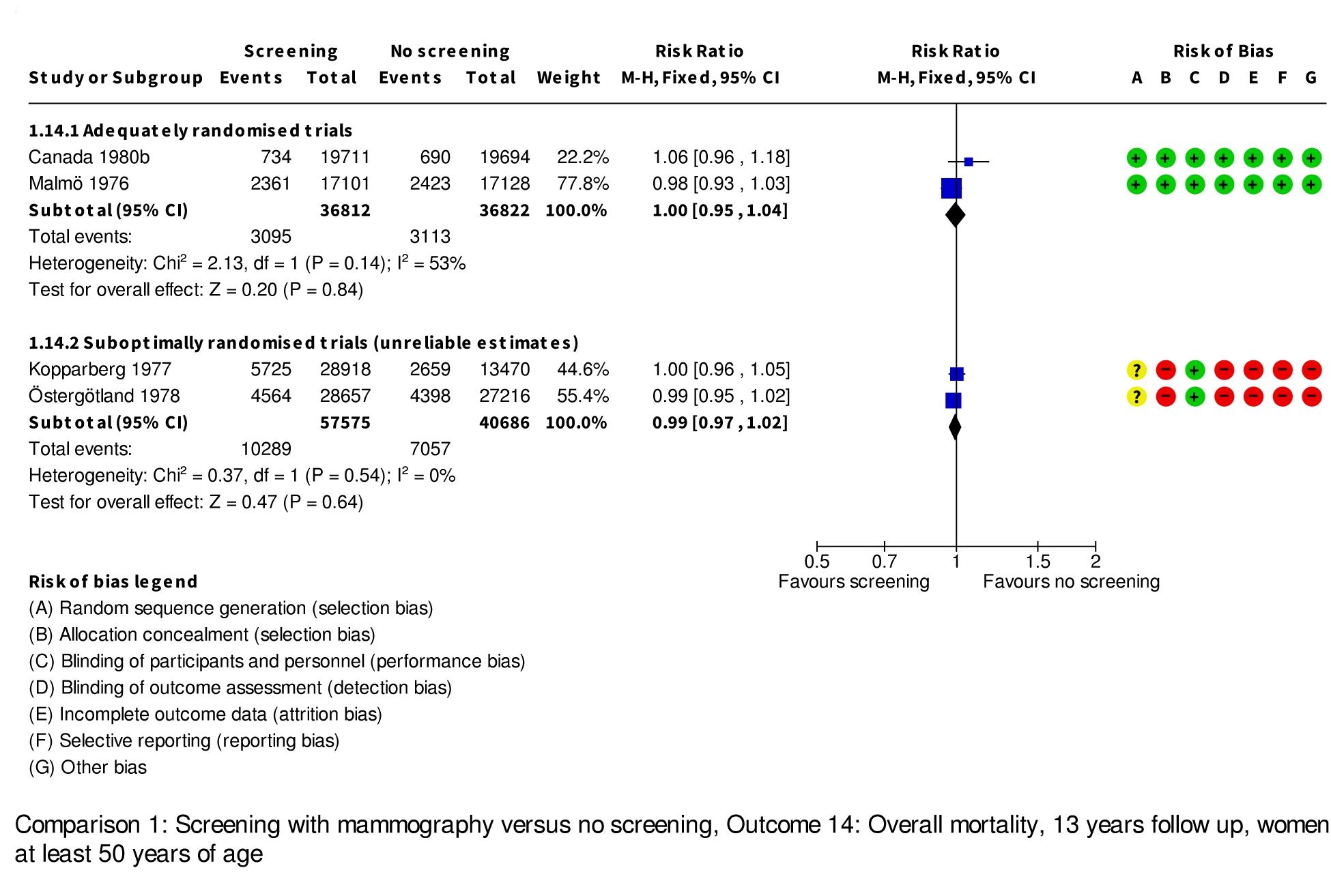

**Analysis 1.15.**
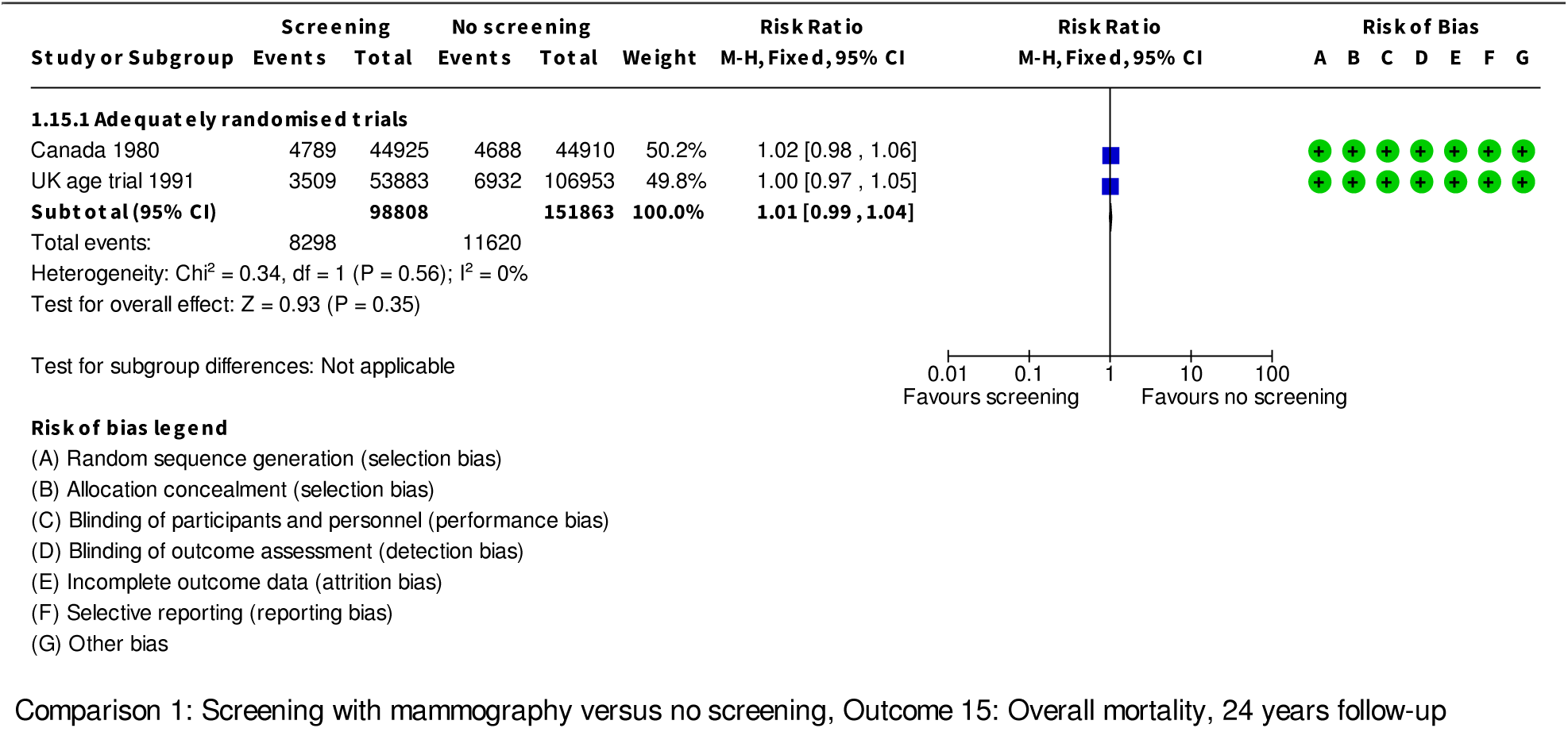

**Analysis 1.16.**
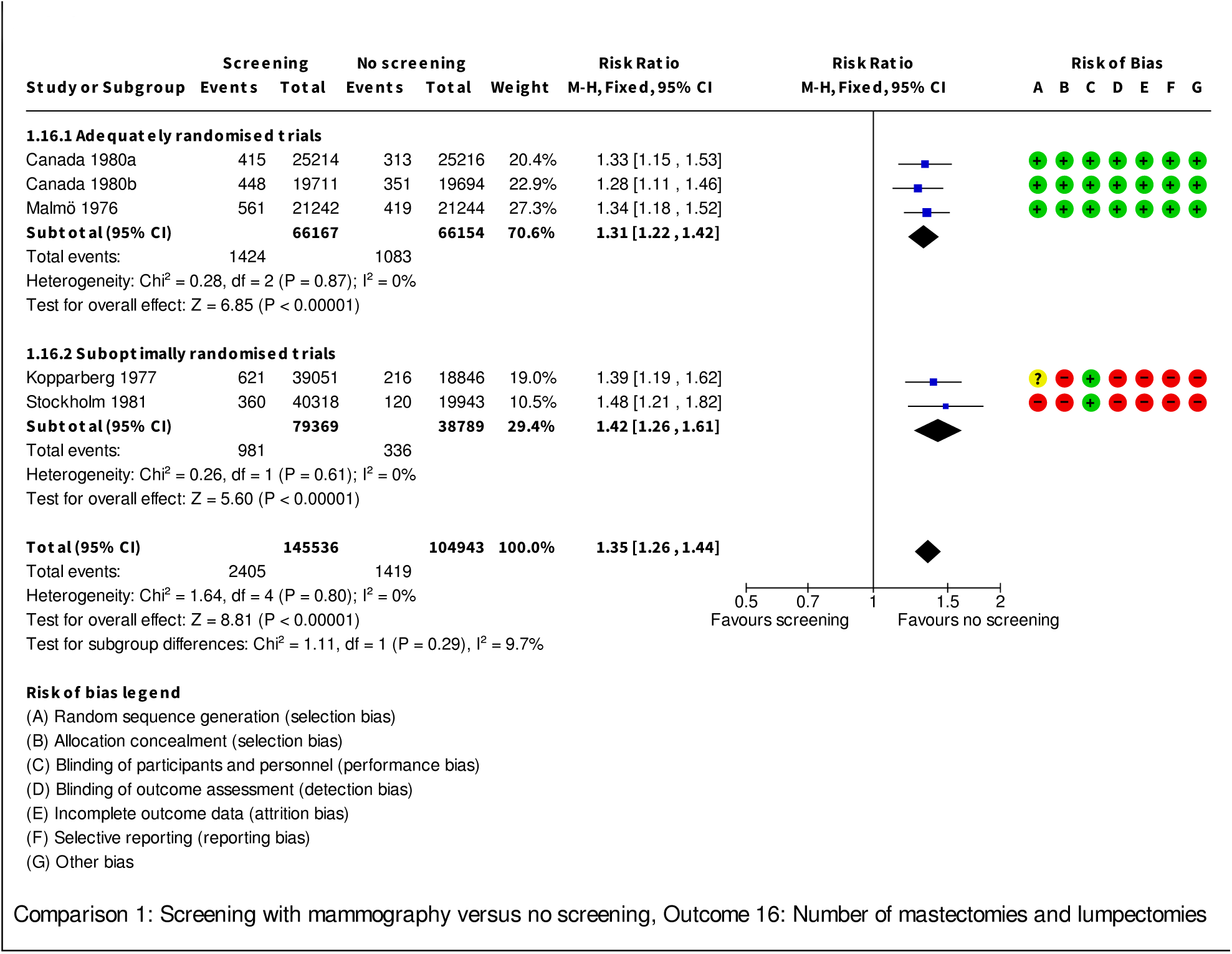

**Analysis 1.17.**
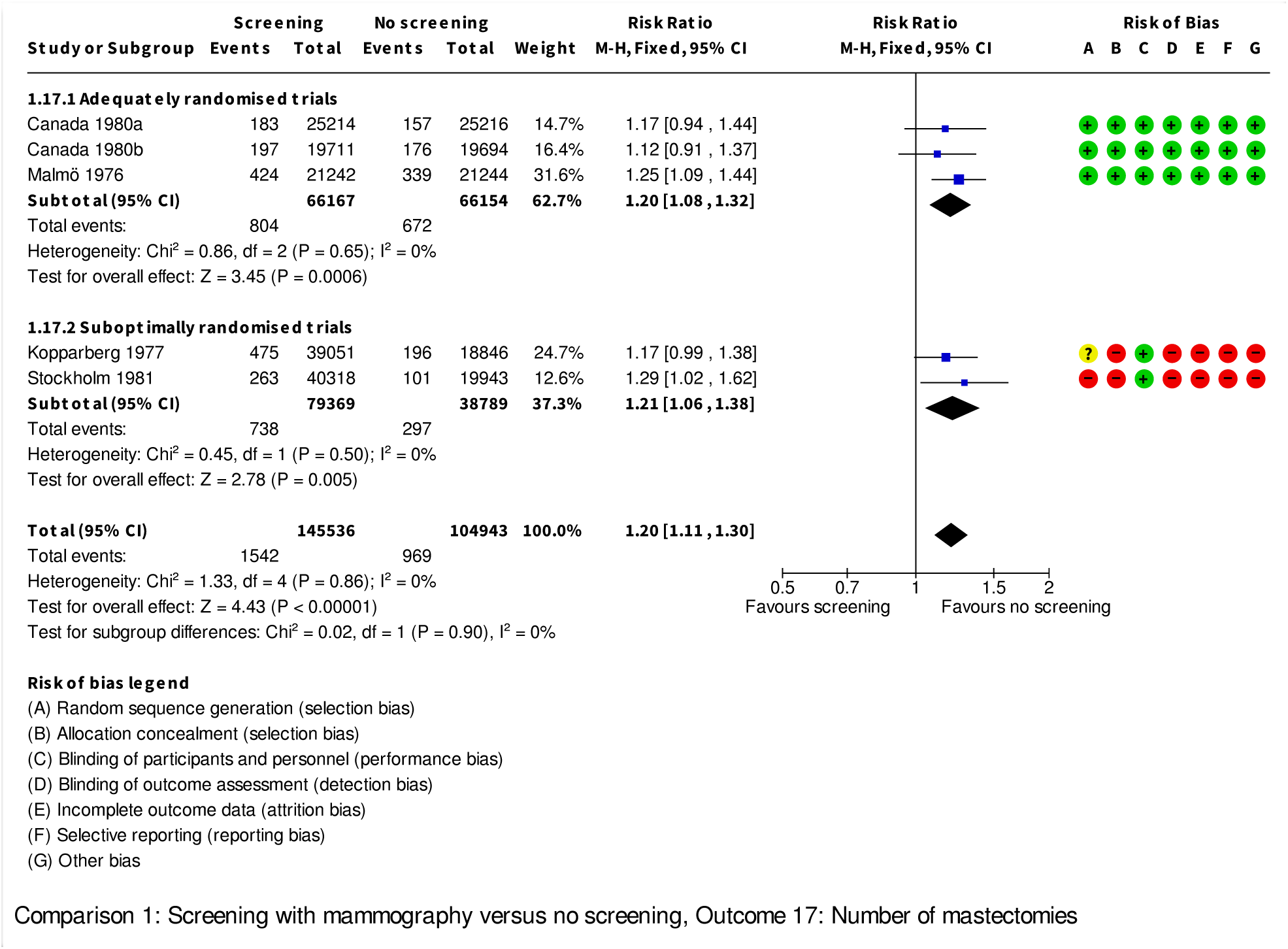

**Analysis 1.18.**
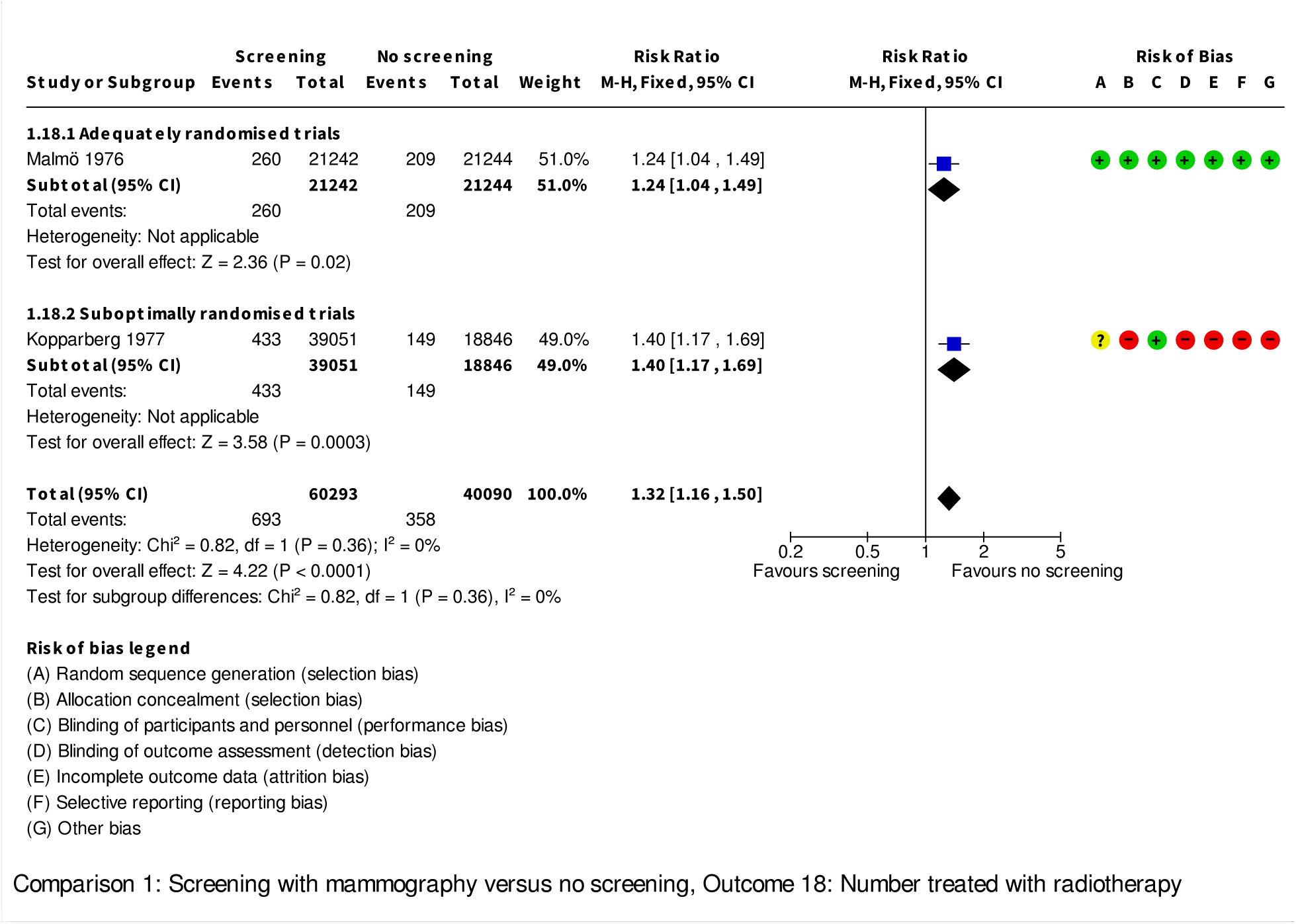

**Analysis 1.19.**
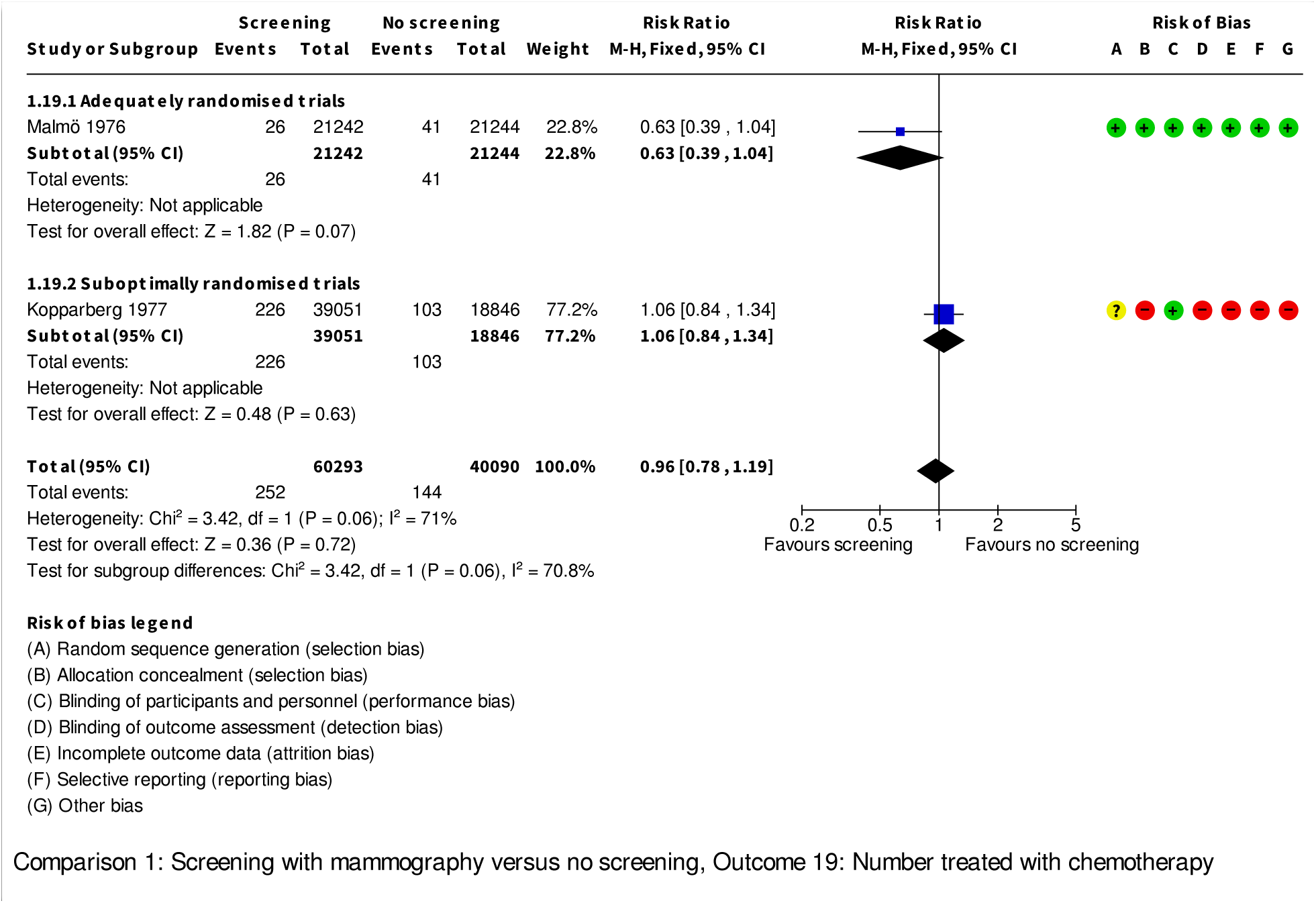

**Analysis 1.20.**
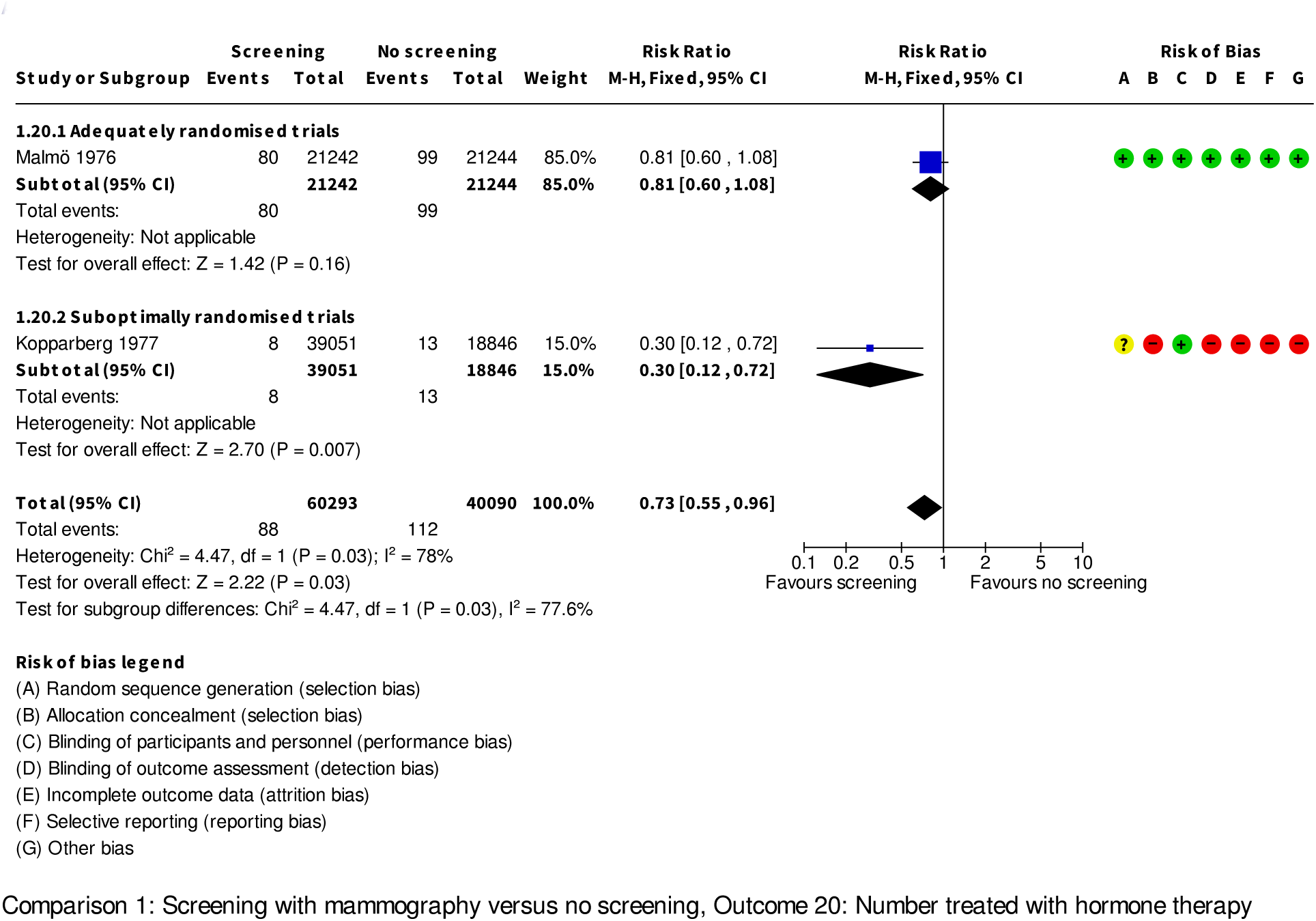

**Analysis 1.21.**
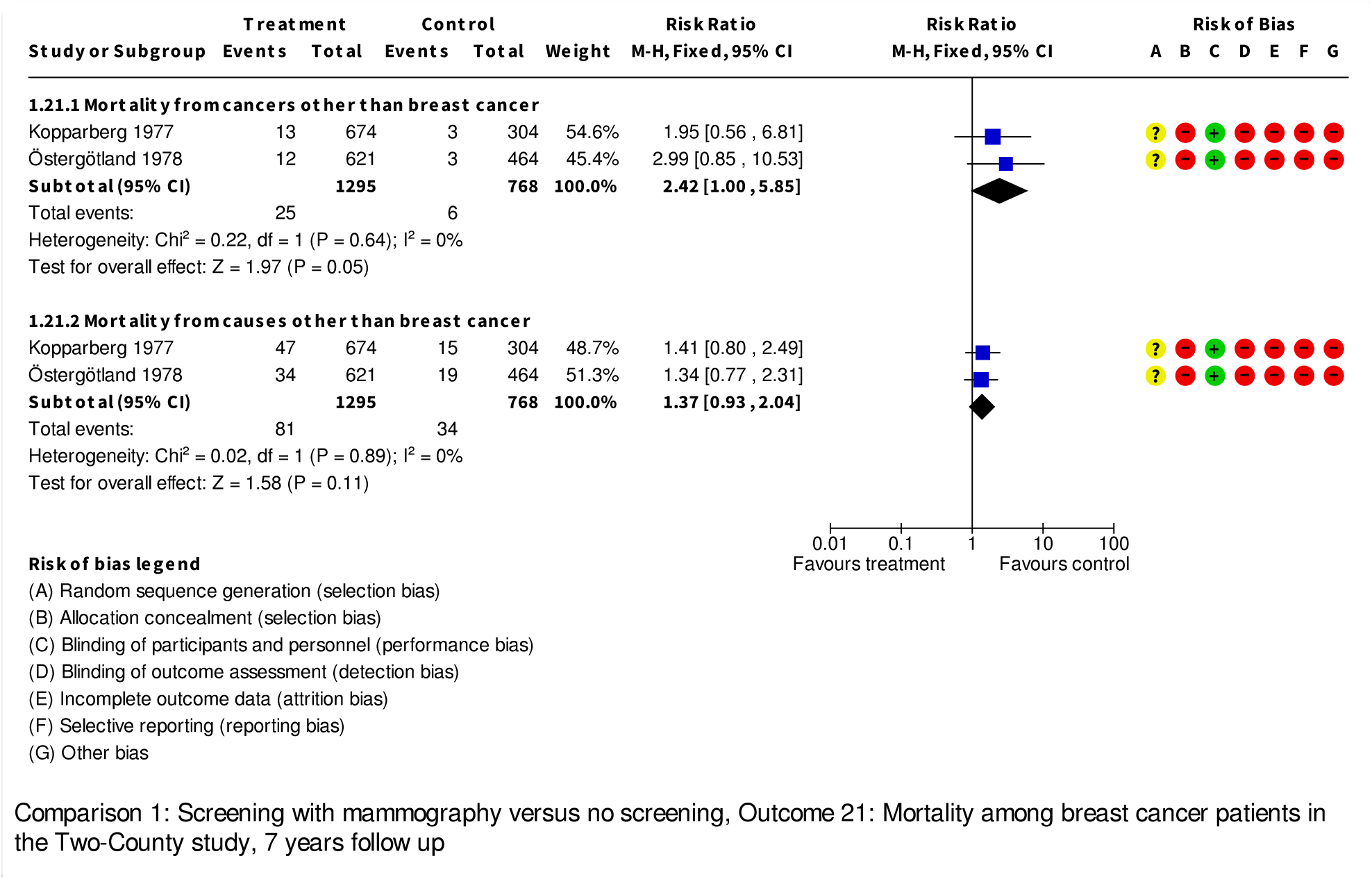

**Analysis 1.22.**
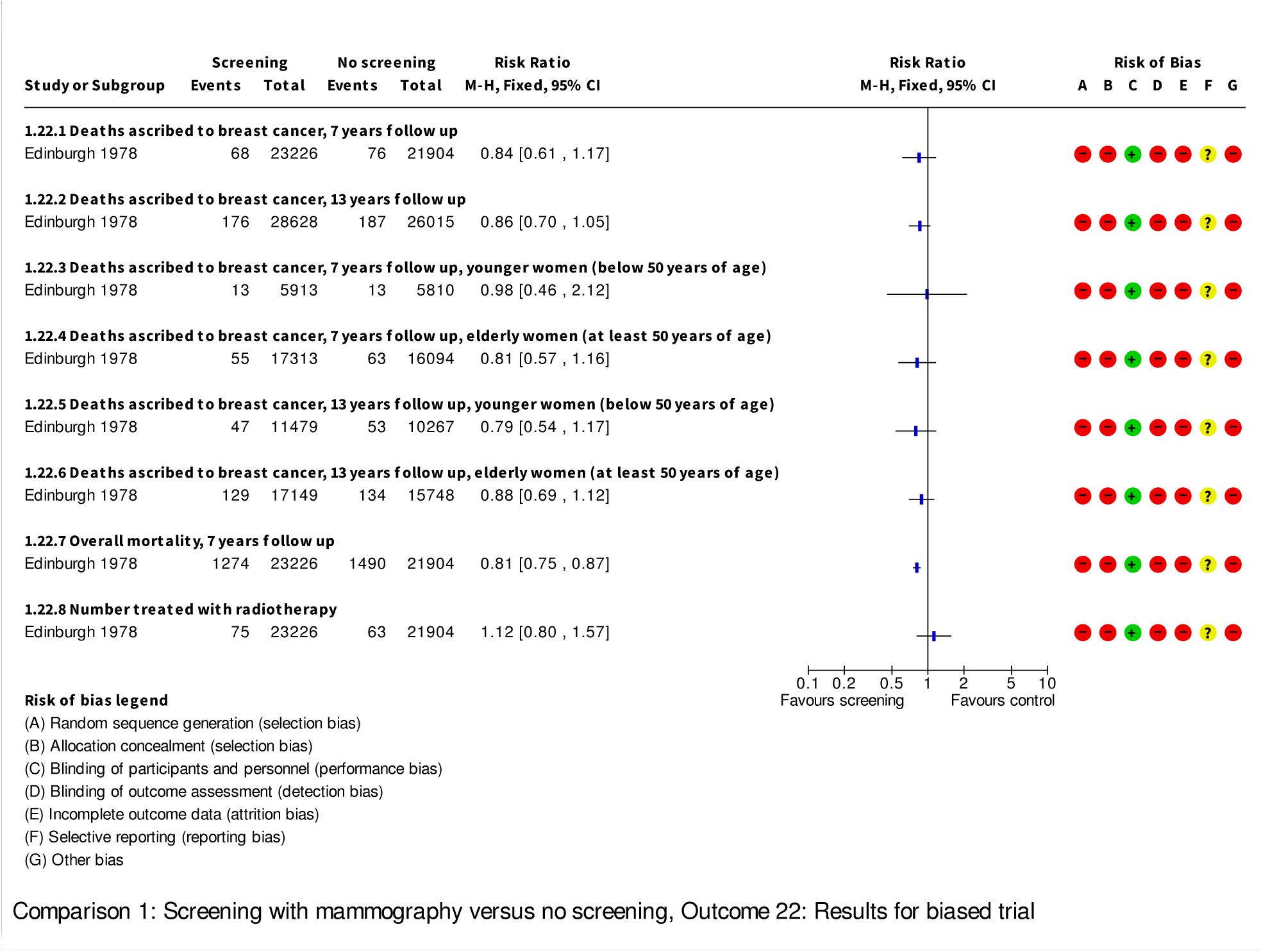

**Analysis 1.23.**
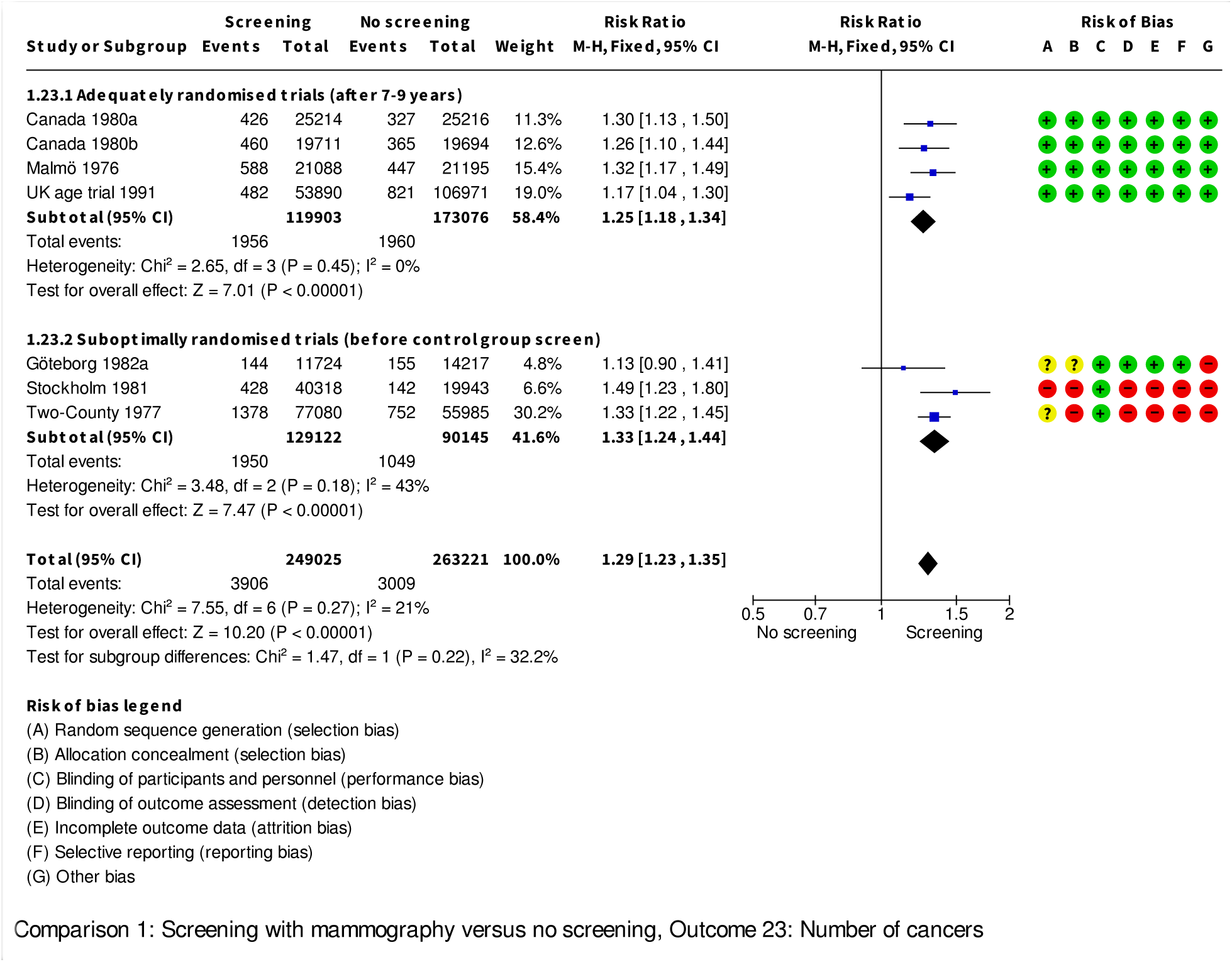

